# What is the effectiveness of interventions to support the mental and emotional health and wellbeing of young people who are not in education, employment or training (NEET)? A Rapid Evidence Review

**DOI:** 10.1101/2025.10.29.25339049

**Authors:** Alesha Wale, Salina Khatoon, Claire Morgan, Amy Fox-McNally, Helen Morgan, Kirsty Little, Hannah Shaw, Kelly Davies, Ben Williams, Olivia Gallen, Jacob Davies, Rhiannon Tudor Edwards, Adrian Edwards, Deborah Edwards, Ruth Lewis

## Abstract

Mental health problems among young people have continued to rise over the last decade. Support services are available in higher education and in many workplace environments. However, while approaches to maintain or enhance mental health and wellbeing within these settings are valuable, it is important to understand what alternative community-based supportive measures or interventions are effective for young people who may not be able to access these services. It is estimated that 13.4% of all young people (aged 16-24) are considered not in education, employment or training (NEET) across the UK. Young people who are NEET have also been found to have a higher incidence of mental health problems than young people who are not NEET.

The aim of this rapid review is to identify the effectiveness of interventions to support mental and health and wellbeing in young people who are NEET. Included studies were published between 2013 and 2025. Nine primary studies, conducted in a range of countries, were included. Studies investigated psychological interventions (n=3), nature-based interventions (n=2), animal assisted interventions (n=2), social prescribing interventions (n=1), and holistic coaching interventions (n=1).

Overall, there is evidence to suggest that interventions delivered in a nonclinical, community-based, or home setting could potentially improve a range of mental health and wellbeing outcomes in young people who are NEET. Participants were generally accepting of the interventions. The psychological interventions were found to improve psychological, social and occupational functioning, reduce difficulties in emotion regulation, reduce psychological distress and led to positive behaviour changes. The nature-based interventions were found to improve social, emotional and behavioural functioning, as well as social connection and mental wellbeing and may be more effective for those who meet the criteria for anxiety or depression. The animal assisted interventions were found to improve social behaviour, and participant abilities however some mixed findings were reported for the impact on self-esteem. The social prescribing intervention improved mental wellbeing and psychological distress. Lastly, the holistic coaching intervention improved participants sense of wellbeing; anxiety; access to peer support resources; knowledge of and access to services; and connection to learning and earning opportunities.

Our confidence in the evidence is limited as the majority of outcomes were only evaluated by a single study. However, while the evidence base identified was limited in number and quality, the findings may help to inform the development and delivery of interventions in young people who are NEET. Given the limited evidence base, robust evaluations should be considered when developing and implementing an intervention for young people who are NEET. Further robust studies assessing long term effects are needed to determine how best to support this population.

## 1. BACKGROUND

### 1.1 Who is this review for?

This rapid review was conducted as part of the Health and Care Research Wales Evidence Centre Work Programme. The research question was suggested by Hywel Dda University Health Board. This review is intended to help support strategic planning and service design, particularly in relation to mental and emotional wellbeing for this vulnerable population. The review is relevant to a wide range of stakeholders, including commissioners of mental health services, local authority teams, third sector organisations, and education and training providers. It may also inform multi-modal interventions (e.g., employability programmes, youth justice services), to improve a range of outcomes for young people who are NEET. Researchers and policy makers may find the evidence useful in informing and shaping regional and national strategies.

### 1.2 Background and purpose of this review

Mental health problems among young people have continued to rise over the last decade (The Health Foundation, 2025). It is estimated that there are almost three times more children and young people accessing mental health services compared to seven years ago. In response to this, the Welsh Government have made several commitments to support children and young people’s mental and emotional health and wellbeing through initiatives such as the <u>‘whole</u> <u>school approach to emotional and mental wellbeing’</u> (Welsh Government, 2021).

While approaches to maintain or enhance mental health and wellbeing within schools, in further education, or the workplace are extremely valuable, it is important to understand what supportive measures or interventions are available for young people in the community who may not be able to access services provided in these environments. The number of young people (aged 16 to 24) who are not in education, employment or training (NEET) across the UK has increased between October to December 2024 compared to the previous year (up 1.3%), with an estimated 13.4% of all young people being NEET (ONS, 2025). In Wales, over the three-year period ending December 2024, the proportion of young people aged 16 to 24 who were NEET varied from 11.4% in the South East, to 14.6% in the Mid and South West (Welsh Government, 2025). Young people who are NEET have also been found to have a higher incidence of mental health problems than young people who are not NEET. In 2021 29.6% of young people who were NEET reported mental health problems compared to 10% of young people who were not NEET (Learning and Work Institute, 2022). This highlights a particularly vulnerable group that may benefit from community-based prevention interventions to enhance mental and emotional wellbeing, build resilience, and reduce reliance on mental health services.

A preliminary literature review identified a lack of existing reviews looking at interventions to support the mental and emotional health and wellbeing of young NEET populations. Therefore, this rapid review aimed to identify and synthesise the evidence for the effectiveness of interventions to support mental and emotional health and wellbeing in young people who are NEET.

## 2. RESULTS

This section details the characteristics of the available evidence base and summary of the findings. The methods used to conduct the review and eligibility criteria for selecting studies for inclusion are detailed in Section 6 of this report. The outcome of the searches and study selection process are presented in Section 7. Section 7 also includes detailed summary of the included studies and an assessment of their methodological quality.

### 2.1 Overview of the Evidence Base

A total of nine comparative primary studies were included in the review. These comprised of different study designs, including one randomised controlled trial (RCT) (Anestis et al., 2020), two non-randomised controlled studies (Kendall & Maujean., 2015; Roemer et al., 2021), six uncontrolled before and after (pre-post) studies (Bertotti et al., 2025; Conway & Clatworthy., 2015; Davies et al., 2020; De Lannoy, Graham & Grotte. 2024; Lee et al., 2013; Wong, et al., 2019). Three of these were mixed methods studies, which also included some qualitative data collection (non-numerical data such as people’s views) (Bertotti et al., 2025; De Lannoy, Graham & Grotte. 2024; Wong, et al., 2019). Included studies were published from 2013 to 2025. Included studies were conducted in a range of countries including the UK (n=2), Australia (n=1), Hong Kong (n=1), Italy and Portugal (n=1), New Zealand (n=1), South Africa (n=1), South Korea (n=1), and the USA (n=1). A total of 1,691 participants were included across all the studies. Sample sizes were generally small, ranging between 21 and 516, with five studies including more than 100 participants and four studies with less than 100 participants. The age ranges varied from 14 to 34 years old across all included studies. Data collection methods utilised self-report methods such as surveys and questionnaires and also included focus groups and interviews. The majority of data collection methods included validated scales and measures.

A range of terms were used within the literature to define those who are considered NEET and some studies included specific populations that fall within the broader category of NEET. Such populations included: NEET (n=3); socially withdrawn youths (n=2); disadvantaged and hard to reach NEET (n=1); disengaged youths (n=1); unemployed young adults (n=1) and those with borderline personality features, self-harm, or suicidal ideation who had dropped out of high school (n=1).

The included studies evaluated a range of intervention types, including psychological interventions (n=3); nature-based interventions (n=2); animal assisted interventions (n=2); social prescribing interventions (n=1); and holistic coaching interventions (n=1). Some interventions included multiple components. A summary of the interventions is presented in Table 1, and a more detailed summary of the included studies can be seen in section 7.2. The interventions were delivered at residential boot camps (n=1), training camps (n=1), organic farms (n=1), sustainable construction sites (n=1), youth centres (n=1), participants’ homes (n=1) and multiple community sites (n=1). Two studies did not state the intervention setting, however one of these involved working with horses. Interventions were delivered through group and/or individual sessions; and facilitated by a range of professionals (including psychologists, psychotherapists, occupational therapists, horticultural therapists, social workers, social prescribers, case workers, link workers, child and youth care workers mindfulness instructors and mentors). Two studies did not provide details of who delivered the intervention.

**Table 1.**
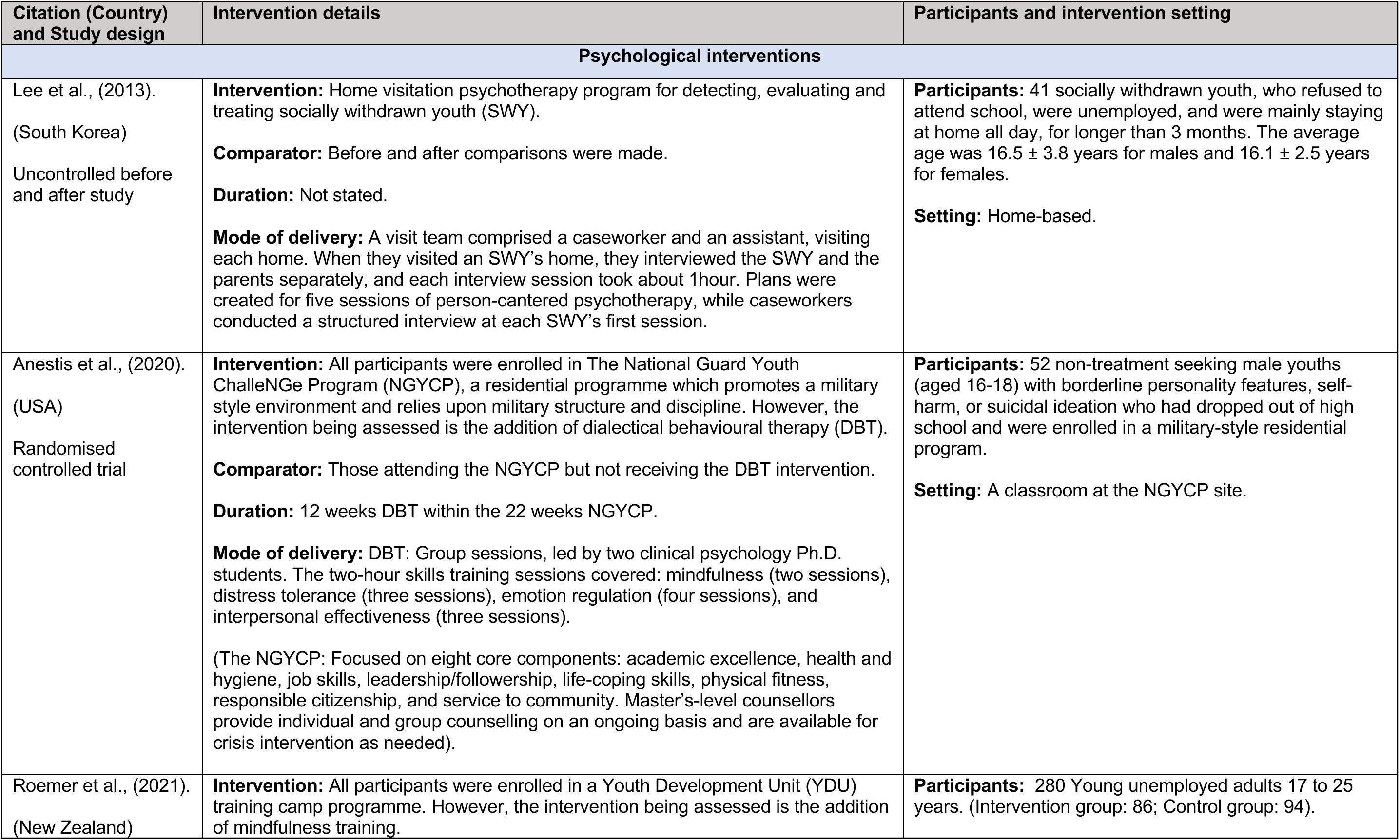
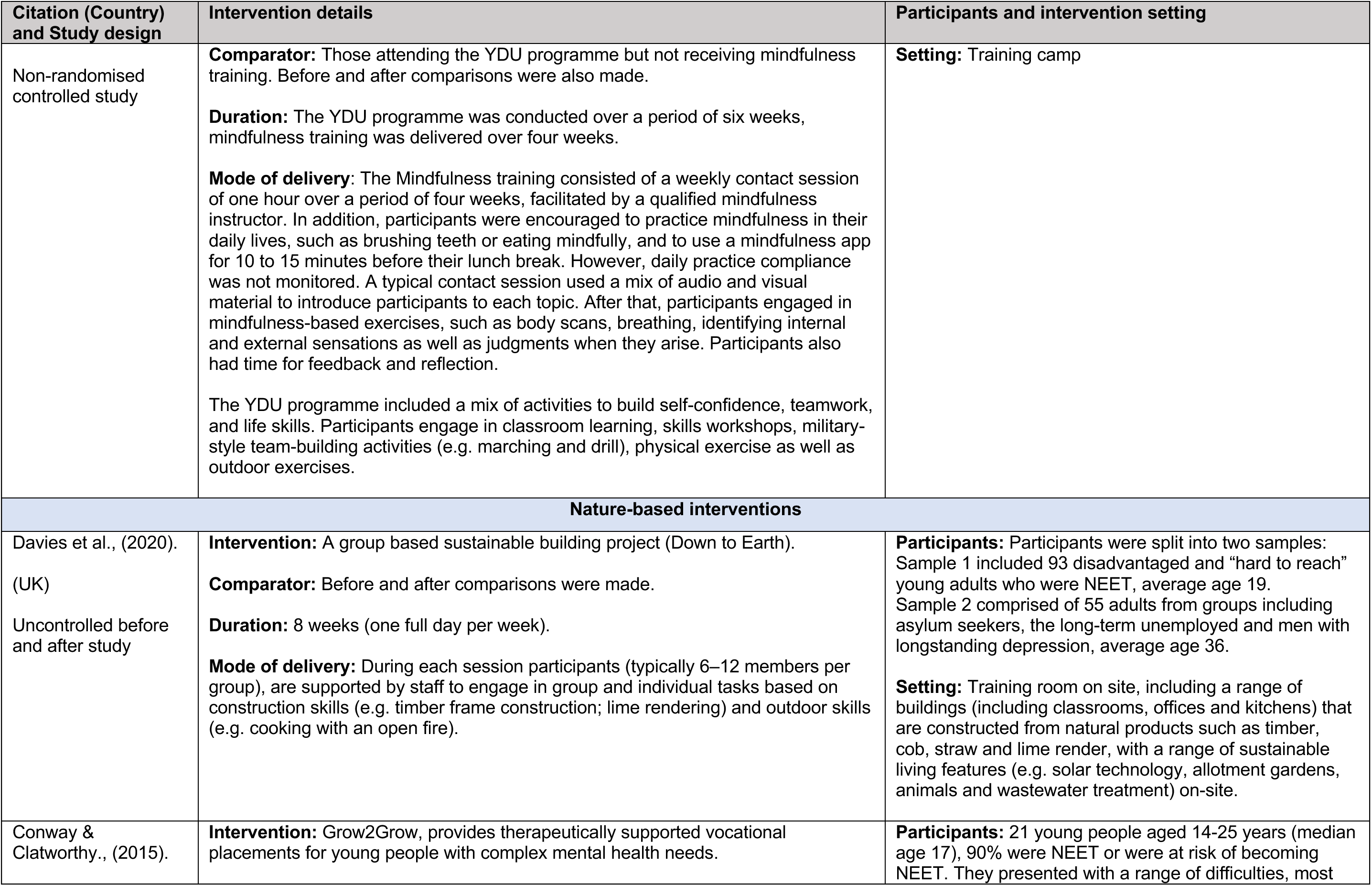
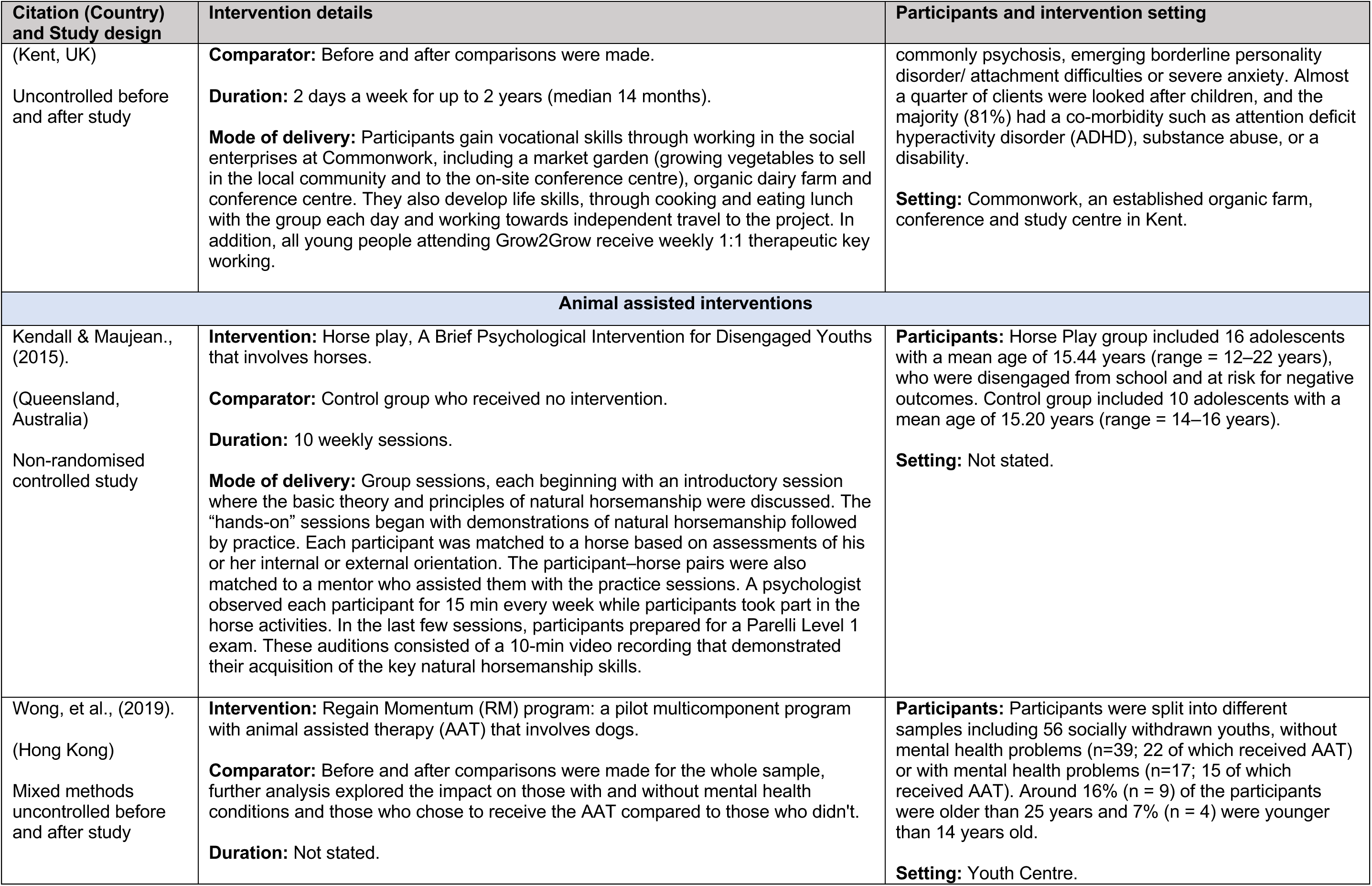
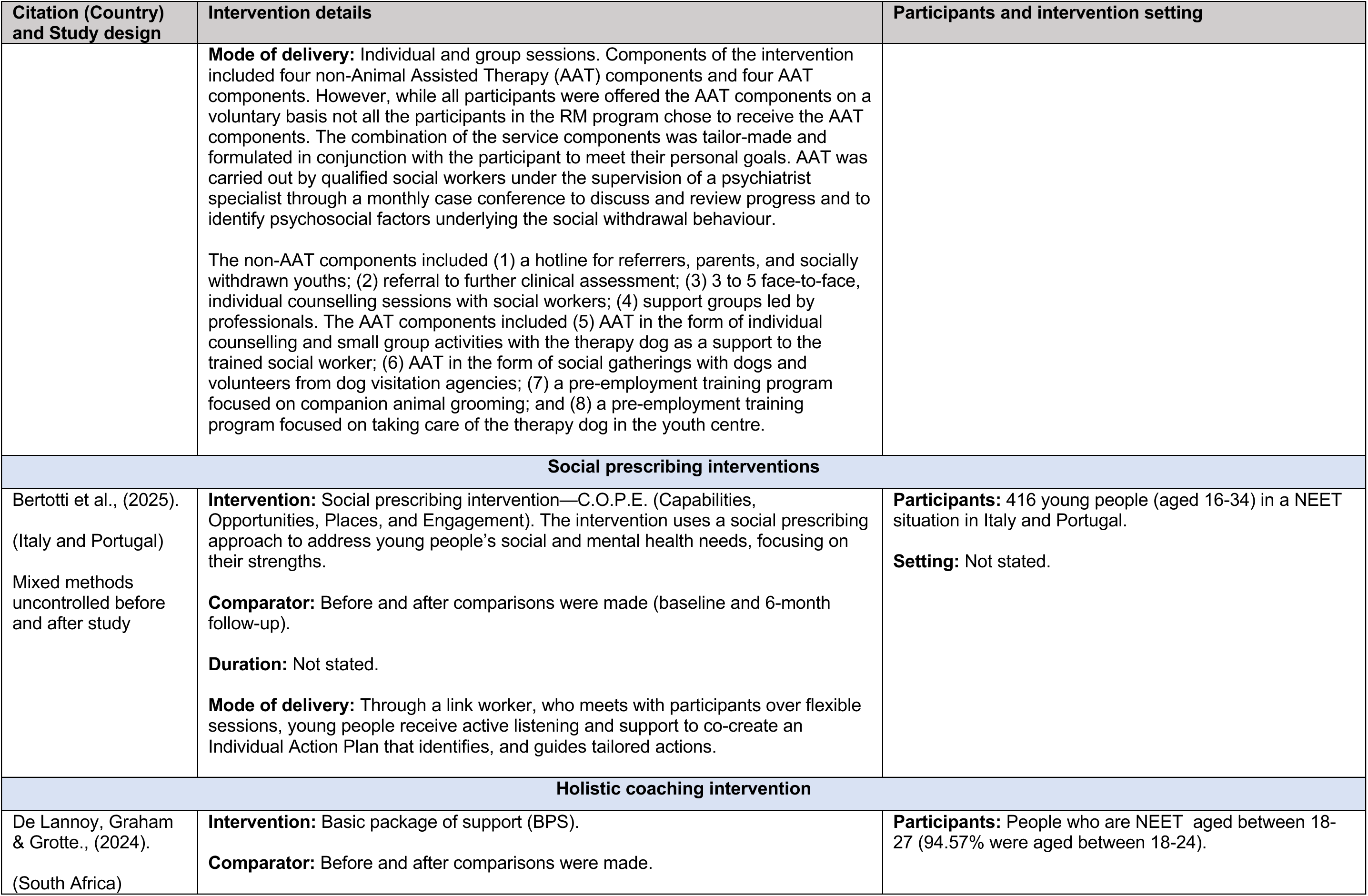
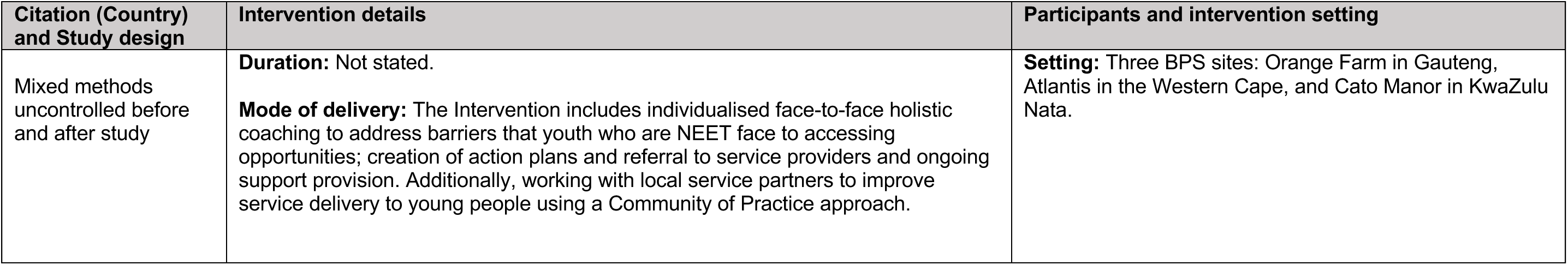
Summary of interventions.

In two studies, participants in the intervention group received an intervention in addition to usual care, and outcomes were compared to a control group who received usual care only. In these studies, the usual care component was a boot camp program (n=1), and a Youth Development Unit (YDU) course (n=1). One study compared the animal assisted intervention with a control group who received no treatment. In six studies, there was no control group, but measured outcomes before and after the intervention within the same population.

A wide range of mental and emotional health and wellbeing outcomes were reported across the studies, a summary of the findings can be found in Table 2. The most common outcomes were mental wellbeing and psychological distress but also included: self-esteem; self-efficacy; depression, anxiety, resilience; social anxiety; distress tolerance; difficulties in emotion regulation; psychological, social and occupational functioning; social, emotional and behavioural functioning; mindfulness; social connection; social behaviour; ability to handle stress, and multiple behaviour changes. Six studies reported participants acceptability of the interventions, three of which were mixed method studies and offered some qualitative insight into participants’ experiences engaging with the interventions. The qualitative findings are reported in section 2.2 alongside the quantitative findings to reflect the participants’ perceptions and views about the individual intervention they attended. While four studies did not state the duration of the interventions, where stated, the interventions ranged from four weeks up to two years. None of the included studies assessed the long-term effectiveness of the interventions. While one study reported findings at 6 months follow-up, the duration of the intervention was not clearly reported, making it unclear if the 6 months follow-up data was collected directly after the intervention or 6 months after the intervention had finished.

**Table 2.**
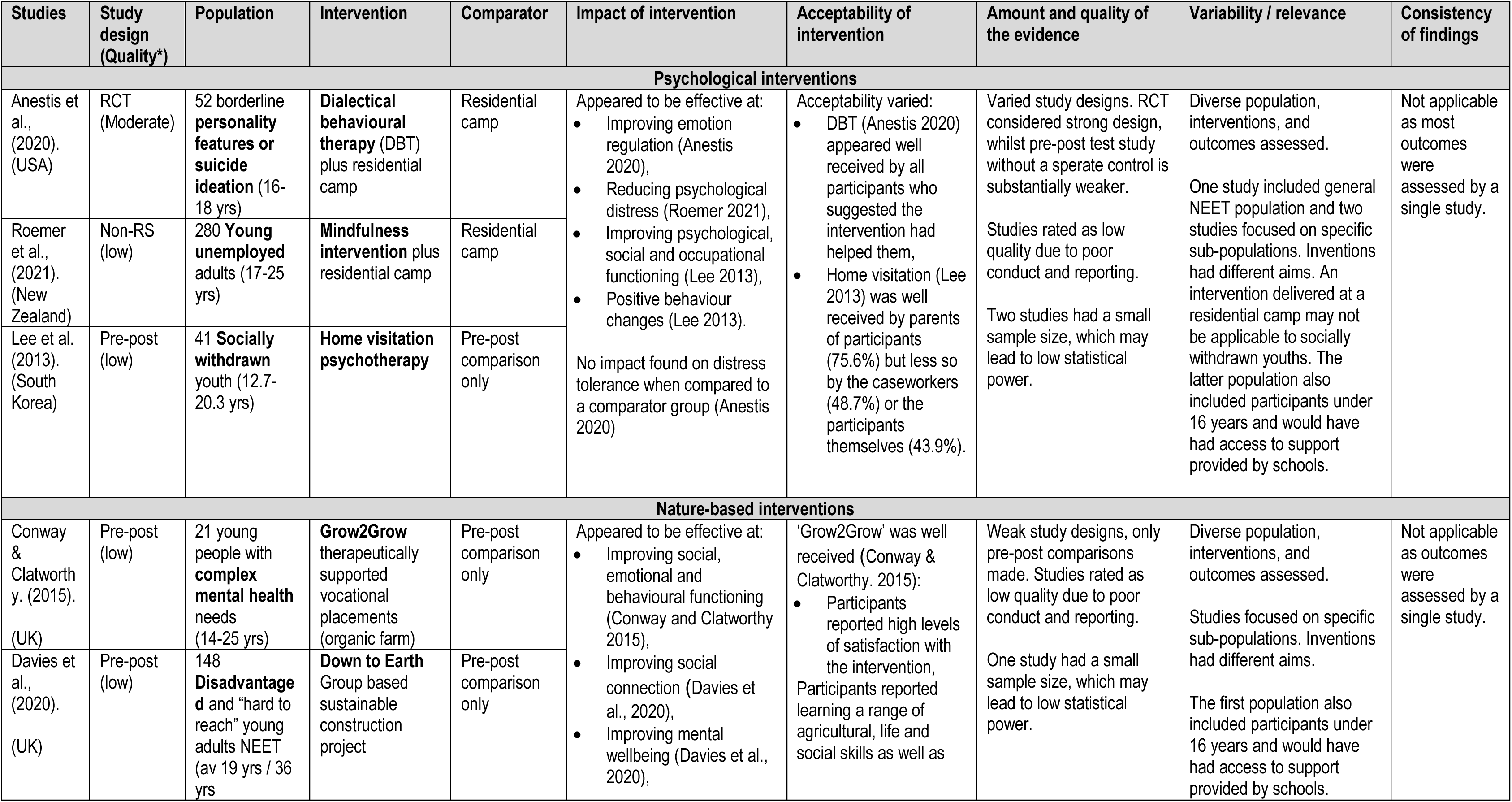
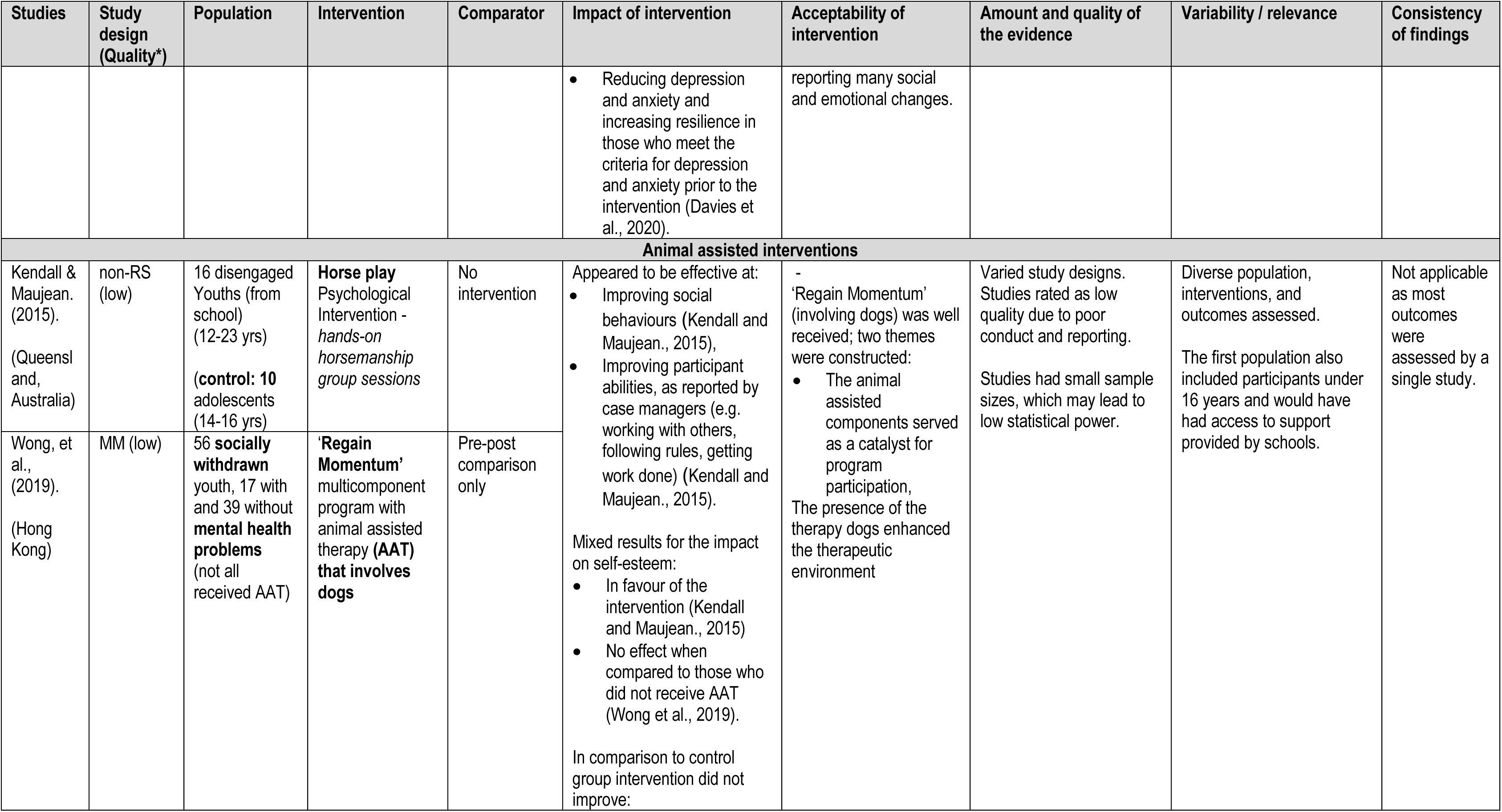
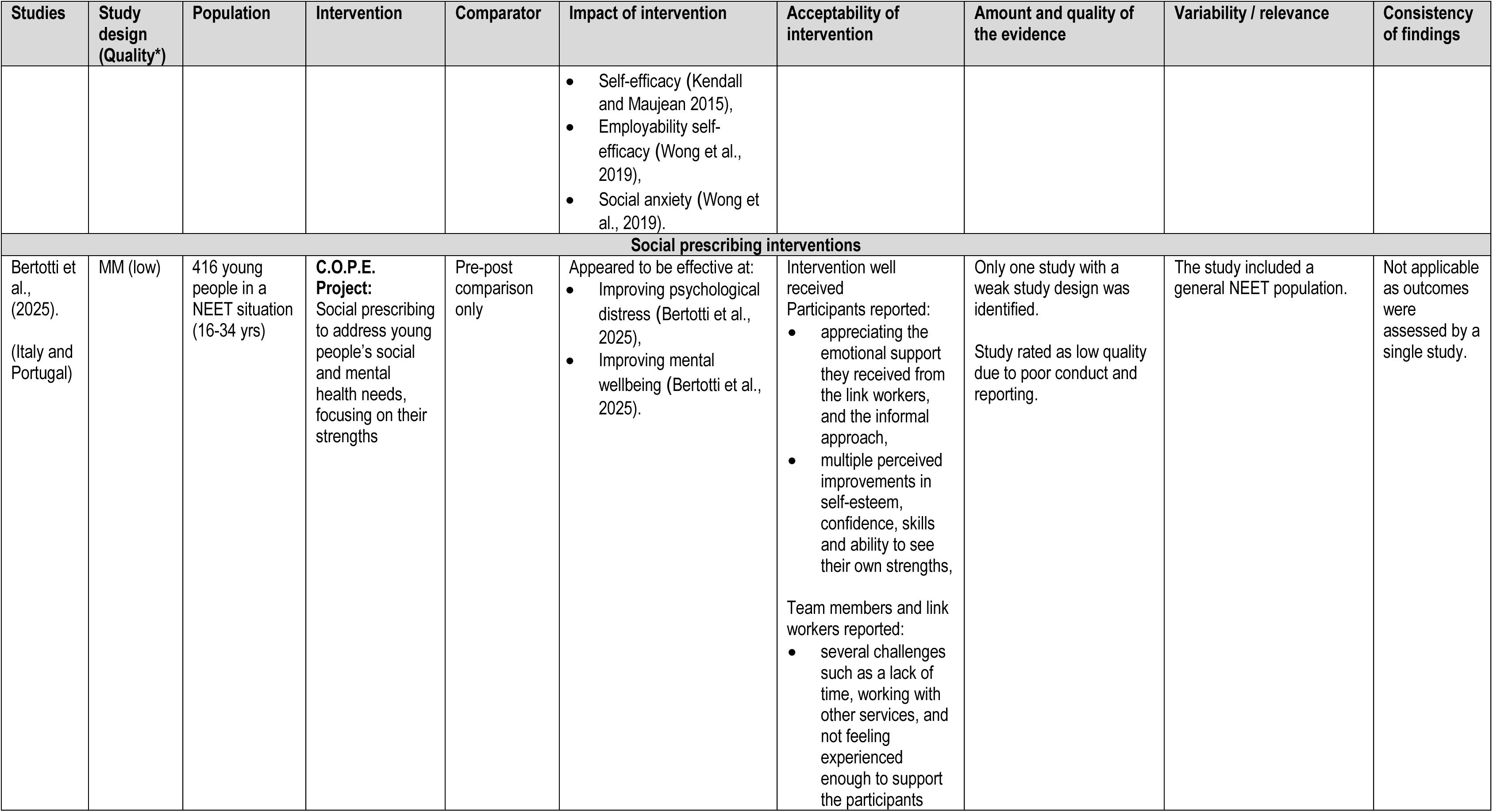
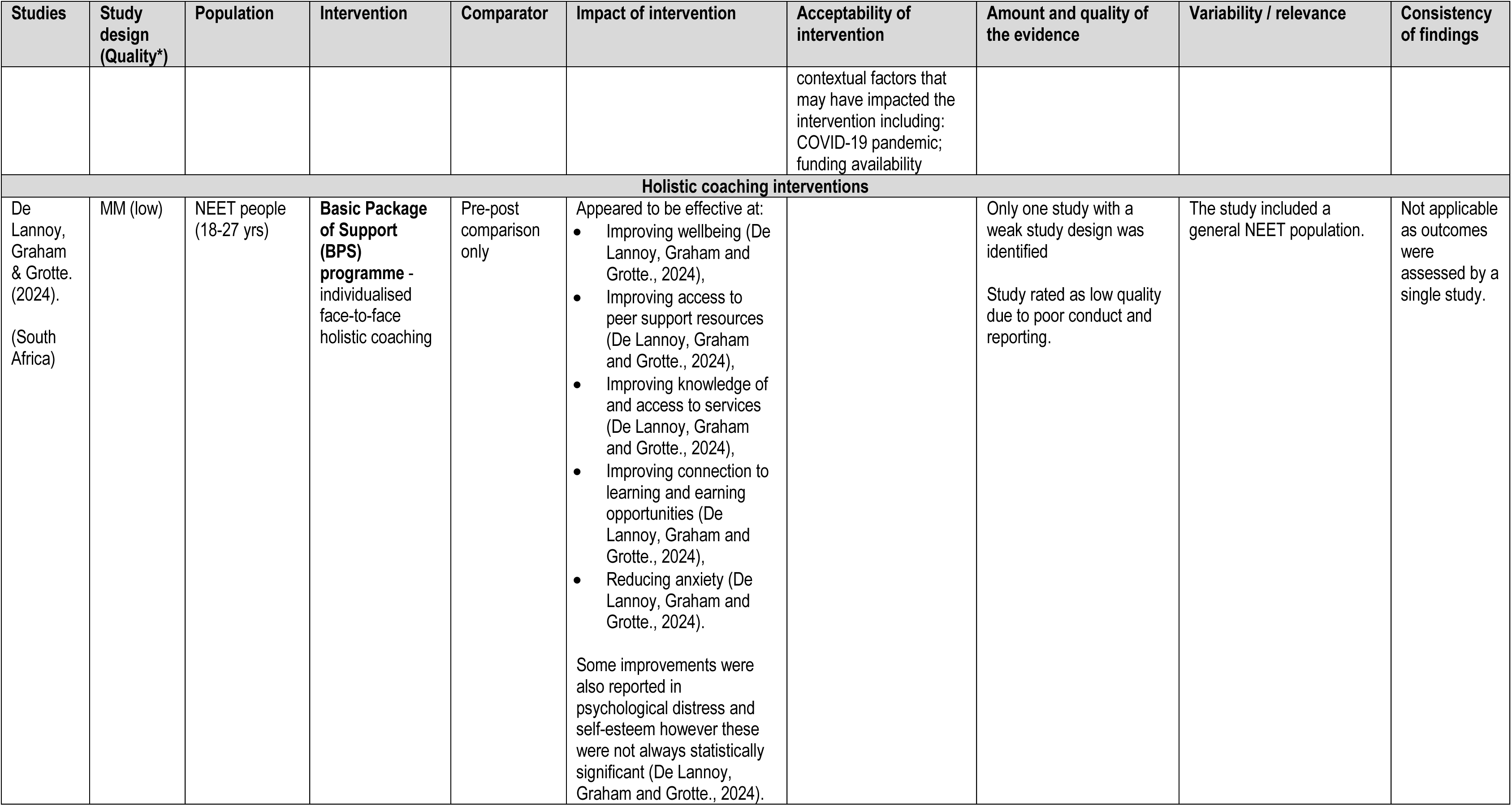
Summary of findings.

The methodological quality of included studies was assessed using appropriate critical appraisal tools for the study designs. All included studies were judged to have some risk of bias, often because of poor reporting of methods. Further details of the quality assessment undertaken for each individual study can be seen in section 7.3. The randomised controlled trial was judged to be of moderate quality due to poor reporting of the randomisation and blinding process, a small sample size, and a lack of a power calculation. The two non-randomised controlled studies were judged to be of low quality due to poor reporting, and a potential lack of appropriate statistical analysis (Kendall & Maujean., 2015; Roemer et al., 2021). One of the studies also reported differences in the characteristics of their control and intervention group which may lead to confounding (Kendall & Maujean., 2015). The three uncontrolled before and after studies were judged to be of low quality and were limited by a lack of a control group and did not provide multiple measurements of the outcomes (both pre and post the intervention); loss to follow-up was also not always adequately described or analysed. For the three mixed method uncontrolled before and after studies, all were considered to be of low quality. All were limited by a lack of a control group. Other limitations included a lack of clarity around whether the participants were representative of the target population; a lack of information on complete outcome data and whether the intervention was administered as intended; divergences and inconsistencies between quantitative and qualitative results were not adequately addressed; and uncertainty around if confounders were adjusted for in the quantitative portion of the study. A lack of integration of the findings from the qualitative and quantitative components of the study was also identified in one study.

### 2.2 Summary of the findings

In this section, the findings have been grouped according to the intervention type and synthesised by the different outcomes measured. The available evidence for each intervention type was assessed in terms of relevance, the amount and quality of the evidence, direction of effect and consistency in the findings. A summary of this can be seen in Table 2.

#### 2.2.1 Effectiveness of psychological interventions

Three studies assessed the effectiveness of psychological interventions on young people who could be considered NEET (Anestis et al., 2020; Lee et al., 2013; Roemer at al., 2021). The studies included one RCT conducted in the USA (Anestis et al., 2020), one non-randomised controlled study conducted in New Zealand (Roemer et al., 2021) and one uncontrolled before and after study conducted in South Korea (Lee et al., 2013).

One RCT, deemed as moderate quality, assessed the effectiveness of a dialectical behavioural therapy intervention (Anestis et al., 2020). Participants in the intervention group received 12 weekly sessions of dialectical behavioural therapy whilst attending a military-style residential programme. Participants in the control group took part in the military-style residential programme only. Participants included non-treatment seeking male youths with borderline personality features, self-harm, or suicidal ideation who had dropped out of high school (Anestis et al., 2020). One non-randomised controlled study deemed as low quality, assessed the effectiveness of a mindfulness intervention in young people who were unemployed members of the Youth Development Unit (a development programme offered by the New Zealand Defence Force). Participants in the intervention group received four weekly mindfulness sessions (Roemer et al., 2021). One uncontrolled before and after study deemed as low quality, assessed the effectiveness of a psychotherapy home visitation intervention in socially withdrawn youth, who refused to attend school, were unemployed, and were mainly staying at home all day and had been for longer than 3 months. The mean number of psychotherapeutic sessions was 2.8 (Lee et al., 2013).

#### Distress Tolerance

Anestis et al., (2020) used the Distress Tolerance Scale to assess distress tolerance before and after the dialectical behavioural therapy intervention. The intervention group showed a statistically significant improvement from baseline in distress tolerance after receiving the intervention (M=39.16; SD:10.65 vs M=46.58; SD:12.67; 95% CI:-12.91 to -1.93; t=-2.76; p<0.01; effect size=-0.50). When compared to the control group however, there was no significant difference in distress tolerance (F(1, 50)=1.30; p=0.26).

#### Difficulties in Emotion Regulation

Anestis et al., (2020) used the Difficulties in Emotion Regulation Scale (DERS) to assess emotional regulation tolerance before and after the dialectical behavioural therapy intervention. The intervention group showed statistically significant improvements for overall emotion regulation after receiving the intervention, compared to before the intervention (M=94.29(SD:23.15) vs M=81.68(SD:23.64); 95%CI:1.73 to 23.50; t=2.37; p<0.05; effect size=0.43). The DERS scale included multiple dimensions and while improvements were seen across them all for the intervention group after the intervention, these improvements were not all statistically significant, (nonacceptance: p<0.01; goals p>0.05; impulse p>0.05; awareness p>0.05; strategies p>0.05; clarity p>0.05). When compared to the control group, a statistically significant difference was reported for overall emotion regulation difficulties (F(1, 50)=6.60; p=0.013), the intervention group reported decreases in difficulties while the control group reported increases in emotion regulation difficulties after the intervention.

#### Psychological distress

Roemer et al., (2021) used the Kessler 10 Psychological Distress Scale (K10) to assess psychological distress before and after the mindfulness intervention. Psychological distress was inversely and significantly predicted by both mindfulness (F (1,172) =28.75; p<0.001) and wellbeing (F (1,172) =7.04; p=0.009). After accounting for this, a reduction in psychological distress was reported for both the control and intervention group. However, those in the intervention group showed a statistically significant greater reduction in distress when compared to the control group (F (1,172) =4.06; p=0.04).

#### Psychological, social and occupational functioning

Lee et al., (2013) used the Global Assessment Functioning scale (GAF) to assess participants psychological, social and occupational functioning after completion of a home visitation psychotherapy programme. A statistically significant improvement in GAF scores was reported for socially withdrawn youth after the intervention when compared to before the intervention (M= 44.6; SD11.1 vs 53.4; SD13.2; p<0.001). However, 48.8% of the participants who received the intervention showed no change in GAF scores (whether this was statistically significant was not reported).

#### Behaviour change

Lee et al., (2013) found that caseworkers reported the following behaviour changes in a number of participants after completion of the home visitation intervention: 14 participants who increased outdoor activities; 13 participants had more family conversations; 11 participants increased interpersonal contacts; 11 participants had improved family relationships; nine participants spent more time outside; six participants increased participation in group activities; four participants returned to school; two participants acquired part-time jobs; and one participant went to a private academy.

#### Acceptability

Two studies reported the acceptability of psychological interventions (Anestis et al., 2020; Lee et al., 2013).

Anestis et al., (2020) used the working alliance Inventory (WAI) and an open-ended treatment evaluation questionnaire to assess acceptability of the dialectical behavioural therapy intervention. The findings from the WAI indicated a strong working relationship between participants and the leaders of the dialectical behavioural therapy sessions, with average scores between 4 and 5 on a 5-point scale. All participants who completed the questionnaire found the group had helped them, 64.3% of participants reported having no complaints or suggestions for improvement. For those that suggested improvements this included more contact with clinicians, and feedback around the snacks provided. Participants reported mindfulness and having a place to discuss emotions as having worked best and some highlighted improvements in their ability to handle their emotions.

Clinicians also provided feedback on their experiences of the dialectical behavioural therapy intervention. They suggested that participants responded well to the behavioural components of the training and were especially engaged when the group used role play to practice interpersonal effectiveness. Participants reported to clinicians that in the boot camp programme, difficult emotions were not openly discussed, however they appreciated the opportunity to discuss emotions and learn skills to cope with emotions in this intervention, instead of ignoring them.

Lee et al., (2013) found that positive responses about the home visitation intervention were reported by 43.9% of participants, 48.7% of the caseworkers and 75.6% of the parents. However, details about what was found to be positive or negative about the intervention were not reported.

#### 2.2.2 Bottom line results for psychological interventions

There is evidence to suggest that psychological interventions may be effective at: reducing difficulties in emotion regulation; reducing psychological distress; improving psychological, social and occupational functioning and may lead to positive behaviour changes (such as improved family relationships, increased engagement in group activities and outdoor activities). However, no significant improvements were found for distress tolerance when compared to a control group.

Acceptability of the psychological interventions varied. The dialectical behavioural therapy intervention appeared to be well received by all participants with suggestions for more sessions to be included. The home visitation intervention was also well received by parents of participants, but less so by the caseworkers or the participants themselves.

The evidence available for the effectiveness of psychological interventions is very limited consisting of one moderate and two low quality studies. There was considerable heterogeneity in the outcomes assessed meaning each outcome was only reported by one or two studies, as such the overall confidence in the findings is low, firm conclusions cannot be made, and further research is needed.

### 2.3 Effectiveness of nature-based interventions

Two studies assessed the effectiveness of nature-based interventions on young people who were NEET (Davies et al., 2020; Conway and Clatworthy., 2015). The studies were both uncontrolled and compared findings before and after the interventions and both were conducted in the UK. One study deemed as low quality involved a group based sustainable building project ‘Down to Earth’, which aimed to improve mental health and social connection through construction and outdoor skills (Davies et al., 2020). The other, also deemed as low quality, aimed to provide combined mental health and vocational support at an organic farm ‘Grow2Grow’, (Conway and Clatworthy., 2015).

#### Social, emotional and behavioural functioning

Conway and Clatworthy (2015), used the Children’s Global Assessment Scale (CGAS) to assess participants’ social, emotional and behavioural functioning on entry to the ‘Grow2Grow’ intervention and at completion. Individual variation in CGAS score was seen over the course of the intervention, with most of the 21 participants showing both increases and decreases. The overall results showed a statistically significant improvement in CGAS after receiving the ’Grow2Grow’ intervention compared to baseline (CGAS Score:35.9 (SD=7.4); vs 47.5 (SD:10.9); t(20)-5.6; p<0.001). However, it was reported that three participants did not improve at the end of the intervention.

#### Mental health: depression, anxiety and resilience

Davies et al., (2020) assessed the impact of a sustainable building intervention ‘Down to Earth’ on multiple mental health outcomes such as depression, anxiety and resilience before and after the intervention. For these outcomes results were combined for both groups within the study: study 1 (NEET population) and study 2 (asylum seekers, the long-term unemployed and men with longstanding depression). The findings show that depression, anxiety and resilience scores were not found to change significantly after the intervention when looking at all participants. However, when the analysis only included those at or above the threshold for depression and anxiety at baseline, statistically significant improvements were seen for depression (n=27; t=-4.21; p<0.001); anxiety (n=43; t=-6.39; p<0.001) and resilience (n=24; t=3.73; p<0.001). A total of 55% of participants who met the criteria for depression at the first data collection point of the study were considered to have met the criteria for recovery after receiving the intervention as well as 49% of those who met the criteria for anxiety.

#### Mental Wellbeing

Davies et al., (2020) assessed mental wellbeing using the Short Warwick-Edinburgh Mental Wellbeing Scale. While a slight improvement was reported, no significant improvements in wellbeing were seen after the ‘Down to Earth’ intervention for the NEET population (Study 1; n=12; M=17.11(SD:1.56) vs M=18.21(SD:2.17); t=-1.31; p=not significant).

#### Social connection

Davies et al., (2020) assessed social connection using an adaptation of the ‘inclusion of community in the self’ scale. After receiving the ‘Down to Earth’ intervention, statistically significant improvements were reported in perceived social connection to the ‘community at large’ (n=23; M=1.78(SD:0.85) vs 2.78(SD:1.24); p<0.005) and to the ‘Down to Earth’ programme itself (n=12; M=2.33(SD:0.78) vs 4.0(SD:0.74); P<0.001) in the NEET population (Study1).

#### Acceptability

Conway and Clatworthy (2015), explored participants satisfaction with the ‘Grow2Grow’ intervention using a project evaluation sheet and a seven-item satisfaction measure. The findings show participants reported a high level of satisfaction with the intervention (mean score of 31.7 (SD:2.3) out of a possible score of 35). On the project evaluation sheets participants reported learning a range of agricultural, life and social skills and many social and emotional changes as a result of the intervention.

#### 2.3.1 Bottom line results for nature-based interventions

There is evidence to suggest that nature-based interventions may be effective at improving social, emotional and behavioural functioning, as well as social connection and mental wellbeing (although the improvement in mental wellbeing was not statistically significant). There is also evidence to suggest that nature-based interventions may be more effective at reducing depression and anxiety and increasing resilience in those who meet the criteria for depression and anxiety prior to the intervention compared to those who do not.

Acceptability was reported for only the ‘Grow2Grow’ intervention, but participants reported high levels of satisfaction and multiple positive impacts.

The evidence available for the effectiveness of nature-based interventions is very limited, consisting of two low quality studies. While a range of outcomes were reported, each outcome is only reported by a single low quality study. As such, the overall confidence in the findings is low, firm conclusions cannot be made, and further research is needed.

### 2.4 Effectiveness of animal assisted interventions

Two studies, both of which were deemed as low quality (Kendall and Maujean., 2015; Wong et al., 2019), assessed the effectiveness of animal assisted interventions on young people who could be considered NEET. The studies included one non-randomised controlled study conducted in Australia (Kendall and Maujean., 2015) and one uncontrolled before and after mixed method study conducted in Hong Kong (Wong et al., 2019).

One study involved a brief psychological intervention ‘Horse Play’, involving horses to improve self-esteem and self-efficacy in disengaged youths (Kendall and Maujean., 2015). The other was a pilot multicomponent programme ‘Regain Momentum’, involving dogs, which aimed to enhance self-esteem and perceived employability, as well as reducing social interaction anxiety in socially withdrawn youth with and without mental health problems (Wong et al., 2019).

#### Self-esteem

Kendall and Maujean (2015), used the Rosenberg Self-Esteem Scale (RSES) before, during and after the ‘Horse Play’ intervention to assess participants’ self-esteem. Compared to before, the results showed a statistically significant improvement in self-esteem after the intervention (Wilks’s Lambda=0.09; F(2,8)=42.72; p=0.0001), as well as statistically significant improvements from pre to mid intervention (p=0.007), mid to post intervention (p=0.002) for the intervention group. This contrasts with the control group who did not improve significantly over time. When the differences in self-esteem score were compared between the intervention and control group, the intervention group was reported to have increased their self-esteem statistically significantly more than the control group (M=7.16; 95%CI: 3.87 to 10.44; eta squared=0.57; p=0.0001).

Wong et al., (2019) also used the RSES before and after the ‘Regain Momentum’ intervention. The results showed a statistically significant improvement in self-esteem for the socially withdrawn participants after the intervention (M=14.5 vs 16.4; d=0.43; p=0.005). However, a non-significant difference was found for those with mental health problems. No significant effects were reported between those who received the animal assisted component to those who did not.

#### Self-efficacy

Kendall and Maujean (2015) used the General Self-efficacy (GSE) scale throughout the course of the ‘Horse Play’ intervention. A statistically significant effect was reported for the intervention group after receiving the intervention (Wilks’s Lambda=0.15; F(2,8)=21.90; p=0.001), as well as statistically significant improvements in self-efficacy from pre to post intervention (p=0.009) and mid to post intervention (p=0.048). However no significant differences were reported and pre to mid intervention (p=0.243). The control group did not improve significantly overtime. However, when compared to the control group there was no significant difference in self-efficacy scores after the intervention between the two groups. (M=3.49; 95% CI: –1.39 to 8.38; eta squared=0.27; p=0.15).

#### Employability self-efficacy

Wong et al., (2019) used the Perceived Employability Self-Efficacy Scale (PESES) to assess employability self-efficacy before and after the ‘Regain Momentum’ intervention. A statistically significant difference was found when comparing PESES scores before and after the intervention for the sample overall (M=44.8 vs 48.6; d=0.96; p<0.001). However, no significantly different effect sizes were reported between those receiving the animal assisted component compared to those who did not.

#### Social anxiety

Wong et al., (2019) used the Interaction Anxiousness Scale (IAS) to assess social anxiety before and after the ‘Regain Momentum’ intervention. Social anxiety after the intervention was reported to be significantly lower than at baseline for the whole sample (M=44.3 vs 47.6; d=1.16; p=0.003). When comparing participants who received the animal assisted therapy (AAT) with those who did not, the mean score was significantly lower after the intervention for those who did not receive the AAT component (p=0.019), but the change in those who did receive the AAT component was not statistically significant.

When comparing socially withdrawn participants with or without mental health problems, the evidence showed a statistically significant reduction in anxiety in both samples (without mental health problems: M=47.4 vs 44.5; d=1.14; p=0.032; with mental health problems: M=48.4 vs 43.7; d=1.21; p=0.017). Upon further exploration it was found that the reduction from baseline to post intervention was only statistically significant for the withdrawn group with mental health problems who did not receive the AAT component (M=48.5 vs 33.5; d=1.48; p=0.014).

#### Social behaviour

Kendall and Maujean (2015), used the Social Behaviour Observation Form (SBOF) to rate participants’ positive and negative behaviours during every session. Social behaviour was reported to be stable across the first 6 sessions and improved during the last two sessions. Statistically significant improvements were found when comparing social behaviour before and after the intervention (M=18.33; SD=5.88 vs M=30.00; SD=6.06; t(3)=–13.22; p=0.001).

#### Case managers perceptions of participants abilities

Kendall and Maujean (2015), asked case managers to complete a survey about their perceptions of participants’ strengths and abilities before and after the intervention. Statistically significant increases in perceived abilities were found in five domains (where significance was considered p<0.1), which included: working with others (p=0.04); following rules (p=0.04); getting work done (p=0.04); energy level (p=0.09); and confidence to try new things (p=0.08). However, no significant changes were reported for participants abilities to work on their own (p=0.29).

#### Acceptability

Wong et al., (2019) conducted qualitative interviews with 10 participants who had received the AAT and found, in general, the intervention was reported to be positive, with no harm being recorded. Two main themes were constructed: The AAT component(s) served as a catalyst for program participation; and the presence of the therapy dog(s) enhanced the therapeutic environment. Three participants reported that they likely would not have joined the intervention if it wasn’t for the AAT components. Engagement through the therapy dog was also found to reduce the stigma of being ‘in counselling’ and helped to reduce anxiety around interacting with the social worker. Overall, taking care of the dog was reported to give the participants a sense of worth and satisfaction:

*“If you helped them [the nonhuman animals], they would show their happiness directly, as if they wanted to say ‘Thank you.”* (Participant from Wong et al., 2019).

#### 2.4.1 Bottom line results for animal assisted interventions

The evidence revealed a mixed picture. There is evidence to suggest that animal assisted interventions may be effective at improving a range of social behaviours and abilities (e.g. working with others, following rules, getting work done). Mixed results were found for the impact of animal assisted interventions on self-esteem. Some improvements were seen in self-efficacy, employability self-efficacy and social anxiety among those who received the intervention. However, when comparing with those who did not receive the intervention or did not receive the animal assisted component of the intervention, the findings were not significant.

Participants who received the animal assisted intervention reported high levels of acceptability, with the presence of the animals appearing to facilitate participation in the intervention and reducing the stigma or anxiety around engaging in the counselling component.

The evidence available for the effectiveness of animal assisted interventions is very limited consisting of two low quality studies. As each outcome is only reported by one or two low quality studies with some mixed findings, the overall confidence in the findings is low. As such, further research would be needed to draw firm conclusions.

### 2.5 Effectiveness of social prescribing interventions

One study assessed the effectiveness of a social prescribing intervention in young people who were NEET (Bertotti et al., 2025). Social prescribing describes a non-clinically focussed intervention that offers a flexible, supportive community-based approach to helping people (Bertotti et al., 2025). The study was an uncontrolled before and after study which utilised mixed methods and was conducted in Italy and Portugal.

The study, deemed as low quality, assessed the effectiveness of the ‘C.O.P.E’ (Capabilities, Opportunities, Places, and Engagement) intervention which aims to address young people’s social and mental health needs, focusing on their strengths (Bertotti et al., 2025).

#### Mental wellbeing

Bertotti et al., (2025) assessed mental wellbeing using the Short Warwick Edinburgh Mental Wellbeing Scale before and after the C.O.P.E intervention. The results showed a statistically significant increase in mental wellbeing at 6-month follow-up when compared to baseline (p<0.05). When looking at the two countries being studied, while the samples were both seen to improve overtime, only the Portuguese sample were seen to improve significantly (p values not reported). However, the mental wellbeing at baseline was lower in the Italian group than in the Portuguese group. Greater positive changes were seen for females and for the younger populations (aged 16-24). The prevalence of probable clinical depression was estimated, and it was suggested that the percentage of participants with probable clinical depression almost halved at 6-month follow-up (23% to 12%).

#### Psychological distress

Bertotti et al., (2025) assessed psychological distress using the Clinical Outcomes in Routine Evaluation - Outcome Measurement (CORE-OM) before and after the C.O.P.E intervention. The results showed a statistically significant decrease in psychological distress at 6-month follow-up (Z=-4.064; p<0.001) across both samples.

#### Contextual factors that may impact the intervention

Bertotti et al., (2025) conducted qualitative interviews with 30 participants, 13 link workers and nine team members to identify outer (high level) and inner (specific characteristics of each intervention) contextual factors that may have impacted the intervention.

Outer contextual factors that may have impacted the intervention were reported. The cost-of-living crisis was reported to have affected the conditions of young people and their parents. The COVID-19 pandemic was also reported as potentially having affected young people by exacerbating their feelings of isolation and mental vulnerability:

> “The only thing that C.O.P.E. might have suffered from COVID especially, is that the isolation that young NEETs are feeling has been made a lot harder.” (C.O.P.E Team member from Bertotti et al., 2025).

Inner contextual factors that may have impacted the intervention were also reported. Interviews revealed that funding availability was seen as a challenge in the project implementation:

> *“The part of co-financing that the partners put in, and in my opinion it was not only numerically in economic terms but also in terms of capital hours more than what was estimated.”* C.O.P.E Team member from Bertotti et al., 2025).

#### Acceptability

Bertotti et al., (2025) also conducted qualitative interviews with 30 participants, 13 link workers and nine team members to identify their acceptability of the C.O.P.E intervention. Respondents reported mechanisms that they felt may have led to the intervention being or not being effective, and perceptions around the intermediate and final outcomes.

Several mechanisms that may have supported the impact of the intervention were highlighted. First, the flexibility in terms of location, timing and availability helped participants to feel comfortable and to build a trusting relationship with those delivering the intervention.

Participants reported appreciating the emotional support they received from the link workers, and the informal approach the link workers adopted:

> *"we talked about my life, about my head, how it was, if everything was okay, psychologically, everything, if I was holding up. To summarise her work more, I think she was almost my ‘real’ psychologist."* (Participant from Bertotti et al., 2025).

Link workers also mentioned the importance of creating trust between the participant and those delivering the intervention to support improved motivation and responsibility but also highlighted the relationship between the parent/carer of the participant and those delivering the intervention. Some link workers suggested the intervention could benefit from an integrated approach with both parents and the participant. The qualitative findings highlighted the importance of trust between participants and those delivering the intervention and suggested the emotional support, and flexibility were facilitators for engagement.

The team members and link workers did highlight some challenges faced that could be improved. This included the underestimation of the time and effort required to support young people who are NEET, especially for link workers who had to balance their role as a link worker with their usual role within their organisation. Link workers often spent additional time outside of contracted hours with participants as they recognised the need for flexibility and the importance of building a trusting relationship which may not be sustainable on a long-term basis:

> *“I was talking to a Link Worker who works at the employment agency, who is very motivated, very convinced, of the method. Unfortunately, the rest of her work requires a very fixed schedule of appointments, so she has a very tight timescale which is not necessarily compatible with the flexibility and speed needed to support the young person.”* (Team member from Bertotti et al., 2025).

Other challenges included working with other services to provide support or opportunities to participants, and link workers not feeling experienced enough to support participants who were NEET. Particularly for those who had experienced extreme social isolation, introversion and those with higher mental vulnerability.

When looking at the intermediate and final outcomes, the interviews highlighted a range of perceived improvements as results of the intervention. This included increased self-esteem, confidence, skills and ability to see their own strengths. The C.O.P.E intervention was well received among participants with some stating it was the first proper support they had received and that they had recommended the project to others. Participants highlighted the benefits of being active again on their mental health and wellbeing:

> *“Simply the fact that I work every day, leave the house even physically is good for me and then from a mental point of view.”* (Participant from Bertotti et al., 2025).

#### 2.5.1 Bottom line results for social prescribing interventions

There is evidence to suggest that social prescribing interventions can improve mental wellbeing and psychological distress in people who are NEET. However, the change in mental wellbeing was not always statistically significant.

The C.O.P.E intervention was well received by participants who reported multiple benefits and those involved in delivering the intervention identified some challenges that could be addressed to further improve the intervention.

The evidence available for the effectiveness of social prescribing interventions is very limited and consists of a single low quality study. As such, the overall confidence in the findings is low, firm conclusions cannot be made, and further research is needed.

### 2.6 Effectiveness of holistic coaching interventions

One study assessed the effectiveness of a holistic coaching intervention in people who were NEET (De Lannoy, Graham and Grotte., 2024). The study was an uncontrolled before and after study which utilised mixed methods and was conducted in South Africa.

The study, deemed low quality, assessed the effectiveness of an individualised face-to-face holistic coaching intervention, the ‘basic package of support’ (BPS) aimed to support young people who are NEET, and to reconnect them to learning and earning opportunities (De Lannoy, Graham and Grotte., 2024). A concurrent triangulation design was used where the quantitative and qualitative data were collected during the same phase of the study, data was then analysed separately, and the findings were then compared. The quantitative evidence was used to establish trends, and the qualitative evidence were used to confirm the trends or provide a more detailed understanding. As such the qualitative findings will be reported alongside the outcomes below.

#### Sense of wellbeing

De Lannoy, Graham and Grotte (2024), used the Cantril’s Ladder and the Life Evaluation Index to assess participants’ sense of wellbeing. A statistically significant improvement in sense of wellbeing after receiving three BPS coaching sessions was found on both Cantril’s ladder (p=0.001) and the Life Evaluation Index (p=0.01), with an increase of over 10 percentage points in the number of participants classed as ‘thriving’ after three coaching sessions compared to baseline.

Qualitative findings support this increase in sense of wellbeing with participants describing how the intervention had helped them to feel more in control and optimistic:

> *“And then, like when I leave, I feel like a little calmer, you know, like I’d come to her before going to work just to talk to her. And then I’d leave for work leaving like feeling, you know, optimistic. And I’d say, hey, I can take this. Yeah. And stuff like that. Coaching helped me cope and feel more optimistic.”* (participant from De Lannoy, Grahem and Grotte., 2024).

#### Anxiety

De Lannoy, Graham and Grotte (2024) used the Generalised Anxiety Disorder (GAD-2) Scale to assess participants anxiety before and after the intervention. A statistically significant reduction in anxiety was reported after receiving three BPS coaching sessions (p=0.0129) with 40% of participants who presented with possible generalised anxiety disorder at baseline and a reduction of 14.85% reported after the coaching sessions.

#### Psychological distress

De Lannoy, Graham and Grotte (2024) used the Kessler Psychological Distress Scale (K6) to assess participants psychological distress before and after the intervention. After three BPS coaching sessions, the percentage of those with moderate to high levels of distress remained the same, however more had moved from high to moderate distress showing a reduction in high distress. The difference was reported to be statistically significant at the 10% level (p=0.0804).

The qualitative findings support the positive shifts reported above with participants describing how the intervention had helped their mental health:

> *“Through the program, I can say that Sis’ M helped me a lot because most of the time I was feeling like small. When you talk to me, I was feeling angry too quick. I was like a heavy man with depression.”* (participant from De Lannoy, Grahem and Grotte., 2024).

#### Self-esteem

De Lannoy, Graham and Grotte (2024) used a five-point scale to assess how participants felt about themselves. No statistically significant improvements in self-esteem were reported after the intervention (p=0.2622). However small improvements were noted, including decreases in the number of participants who: felt they could not do anything right (3.55% point reduction); or felt there were things they didn’t like about themselves (2.30% point reduction). There was also a small increase reported in the number of participants who felt good about themselves (3.00% point improvement).

#### Sense of support

De Lannoy, Graham and Grotte (2024) assessed participants perceived sense of support before and after receiving the BPS coaching sessions. No significant differences were reported overall (p=0.791), however there was a small decrease in the number of participants feeling connected most of the time and a reduction in the number of participants reporting feeling isolated, alone or disconnected from others was seen. The biggest increase was seen in the number of participants ‘kind of feeling connected’ (almost 9% increase). However, it was unclear if these changes were statistically significant.

The qualitative results show the importance of a good relationship between participants and those delivering the intervention in terms of fostering a sense of support:

> *“Just being able to speak to my coach about anything. She was not exposing me, and she was not talking about me. I felt like I needed to talk to someone because at that time, I felt like I had a lot of anger […] I didn’t know how to express my feelings towards anyone, and I hated people.”* (participant from De Lannoy, Grahem and Grotte., 2024).

#### Access to peer support resources

De Lannoy, Graham and Grotte (2024) assessed participants perceived access to peer support resources after receiving the BPS coaching sessions. Statistically significant improvements were seen after three BPS sessions (p=0.0337). The biggest increase was seen in the number of participants reporting to ‘have friends that they could trust and rely on’ (almost 8% increase).

#### Ability to handle stress

De Lannoy, Graham and Grotte (2024) assessed participants perceived ability to handle stress before and after receiving the BPS coaching sessions. The findings were mixed and the overall difference in mean scores was not statistically significant (p=0.6547). The biggest increase reported was in those who felt they were learning to deal with stress and emotions (15% increase) after the intervention. However, it was unclear if this difference was statistically significant.

Qualitative findings showed that participants reflected on the positive impact on the intervention on their stress levels:

> *“I felt comfortable [a lot] because they welcomed me really well and at the present moment, I don’t have any stress. I feel like I’m at home.”* (participant from De Lannoy, Grahem and Grotte., 2024).

#### Asking for help when upset and feeling overwhelmed

De Lannoy, Graham and Grotte (2024) assessed how often participants ask for help when feeling upset or overwhelmed before and after receiving the BPS coaching sessions. No significant differences were reported (p=0.1517). However, there was a decrease in the number of participants who felt they could not be helped or did not seek help (8.28% reduction).

Qualitative findings showed the positive impact the intervention had on how participants dealt with stress:

> *“Also, rejection is part of life, and I have accepted that, so it doesn’t hurt as much because the way to success passes through that bridge. I will continue to try until I get something. Even when I’m not okay I will come to BPS and maybe talk to someone.”* (participant from De Lannoy, Grahem and Grotte., 2024).

#### Knowledge of, and access to services

De Lannoy, Graham and Grotte (2024) assessed whether participants accessed services and resources in the community before and after receiving the BPS coaching sessions. Statistically significant improvements were found in the number of participants reporting that they were aware of services and resources in the community that were available to them (p=0.0000).

Qualitative findings showed how the intervention had helped participants to be aware of, and how to engage with, services available to them:

> *“You come and it’s almost like they take your hand and that’s how my experience was to get from this point to the next. That’s when I was like okay if I for instance if I want to apply now for my license, I know this is where I need to go and get the training I need.”* (participant from De Lannoy, Grahem and Grotte., 2024).

#### Youth connect to learning and earning opportunities

De Lannoy, Graham and Grotte (2024) assessed the number of youths transiting from NEET to EET throughout the intervention. The sample for each data collection point was dependent on the number of participants who had completed that stage of the intervention, therefore the sample used for this outcome was different from the outcomes reported above. Statistically significant results were reported showing a decrease in the proportion of NEET youths as the coaching sessions continued, with 88.39% of participants being NEET after session one and 64.37% after session four (p=0.0000).

Qualitative findings showed how the intervention had helped to increase participants’ motivation to learn and earn:

> *“But now I am convinced [that I must also continue studying] because […when we drew up my action plan], I looked and yoh: if I stop, I cannot go further, so I need to go. So, the next thing was ‘OK, we’re gonna get that weekend job’. [The team helped me apply] and just a few weeks into college, I got a phone call [and then I got the job].”* (participant from De Lannoy, Grahem and Grotte., 2024).

#### 2.6.1 Bottom line results for holistic coaching interventions

There is evidence to suggest that holistic coaching interventions can improve participants sense of wellbeing; anxiety; access to peer support resources; knowledge of and access to services; and connection to learning and earning opportunities. Some improvements were also reported in psychological distress and self-esteem; however these were not always statistically significant. Mixed findings were reported for sense of support; ability to handle stress; and asking for help when upset and feeling overwhelmed.

Qualitative findings supported and provided context for the positive changes seen in outcomes post intervention and overall reflected a positive experience with many perceived benefits reported by the participants.

The evidence available for the effectiveness of holistic coaching interventions is very limited and consists of a single low quality study. As such, the overall confidence in the findings was low, and further research is needed.

## 3. DISCUSSION

### 3.1 Summary of the findings

This rapid review aimed to identify and synthesise the evidence for the effectiveness of interventions to support mental and emotional health, and wellbeing in young people who are NEET. A range of interventions were identified across nine studies, including psychological interventions (n=3), nature-based interventions (n=2), animal assisted interventions (n=2), a social prescribing intervention (n=1), and a holistic coaching intervention (n=1).

The evidence suggests that interventions delivered in a nonclinical, community-based, or home setting could potentially improve a range of mental health and wellbeing outcomes in young people who are NEET. The psychological interventions were found to improve psychological, social and occupational functioning, reduce difficulties in emotion regulation, reduce psychological distress and led to positive behaviour changes. The nature-based interventions were found to improve social, emotional and behavioural functioning, as well as social connection and mental wellbeing and may be more effective for those who meet the criteria for anxiety or depression. The animal assisted interventions were found to improve social behaviour, and participant abilities however some mixed findings were reported for the impact on self-esteem. The social prescribing intervention improved mental wellbeing and psychological distress. Lastly, the holistic coaching intervention improved participants sense of wellbeing; anxiety; access to peer support resources; knowledge of and access to services; and connection to learning and earning opportunities.

The evidence highlights that young people who are NEET are a complex population group with varying needs. Many of the studies included specific sub-populations within the broader category of NEET such as socially withdrawn youth, disengaged youth, and those with complex mental health needs etc. As such, some interventions may be more suitable or applicable than others depending on an individuals’ circumstances. This should be considered when developing or planning any new interventions, for example those who suffer from social anxiety may be less likely to attend the residential camps or group-based interventions.

The most applicable to the Welsh context may be the nature-based interventions, as both studies were conducted in the UK and showed promising results. The findings from one study suggests that the intervention may be more effective for those who meet the criteria for anxiety or depression (Davies et al., 2020). Participants were highly satisfied with the ‘Grow to Grow’ nature-based intervention, reporting learning a range of agricultural, life and social skills and experiencing many social and emotional changes as a result of the intervention (Conway and Clatworthy 2015) (acceptability of the other nature-based intervention was not assessed).

Participants were generally accepting of all of the interventions that assessed this outcome and often felt empowered by them. Participants highlighted factors that they found particularly helpful such as: the relationship with those delivering the intervention; the emotional and physical support they received, having a place to share and discuss emotions; and being able to engage with animals or nature.

However, the overall evidence base was limited with only nine studies being included in this review. Due to the limited number of studies available for each intervention type, the low quality of reporting within these studies, and the heterogeneity in outcomes resulting in most outcomes only being reported by a single study, the overall confidence in the findings is low and further evidence would be needed to draw any firm conclusions.

### 3.2 Strengths and limitations of the available evidence

The evidence base suggests that a range of different intervention strategies to improve the mental and emotional health and wellbeing of young people who are NEET may be effective. While multiple benefits were reported across the interventions, the addition of the mixed methods studies provided additional evidence for the acceptability of these interventions and provide some detail around factors that may improve engagement with future interventions.

However, the term NEET is not consistently used or defined within the literature across different countries. While methods were used to ensure that the majority of participants in any included study could be considered NEET and were aged between 16-24, the population groups were not always clearly defined and are likely to vary across studies.

In addition, the available evidence was primarily of low quality, with only one RCT being identified as moderate quality. The predominantly low quality of the included studies and the limited evidence base available for each intervention type reduces the overall confidence in the findings, which should be interpreted with caution.

In some studies, the impact of an additional intervention component was assessed against a comparator group receiving the original intervention. For example, those receiving dialectical behavioural therapy and a training camp intervention were compared against those who received the training camp intervention only. As such it is not always clear if the effects reported may have been influenced by the wider components of the intervention, in this case the training camp.

None of the included studies assessed the long-term effectiveness of the interventions. While one study reported findings at 6 months follow-up, the duration of the intervention was not clearly reported, making it unclear if the 6 months follow-up data was collected directly after the intervention or 6 months after the intervention had finished. As no long-term results were reported, it is unclear if any of the effects reported by the included studies would remain consistent after a period of time.

The evidence base was conducted across a range of countries which may limit the generalisability of the findings to the Welsh context.

### 3.3 Strengths and limitations of this Rapid Review

This rapid review utilised abbreviated systematic review methods to generate the evidence in a timely manner whilst maintaining attention to bias. However, it is possible that additional eligible publications may have been missed, which may bias the rapid review findings. The included studies were identified through an extensive search of electronic databases and citation tracking. Due to the limited timeframe to complete this rapid review, as well as the large number of references initially identified from literature searches, most records were screened by a single reviewer at title and abstract level. However, just over 10% of the references (n=1,058) were double screened for consistency. All full text records were screened by two reviewers for consistency, with any conflicts being discussed and resolved within the team.

As this review aimed to explore interventions to support the mental and emotional health and wellbeing of young people who were NEET, re-engagement interventions aimed at facilitating young people to engage in education, employment or training were excluded. However, some re-engagement interventions may also incorporate components aimed to improve the mental and emotional health and wellbeing of young people who were NEET and therefore may provide useful evidence for developing further interventions.

### 3.4 Implications for policy and practice

This rapid review identified a paucity of high quality research assessing the effectiveness of interventions to support the mental and emotional health and wellbeing of young people who are NEET. The limited number of studies identified for each intervention type as well as considerable methodological limitations reduces the overall confidence in the findings. However, the findings could help to inform the development and delivery of future interventions in young people who are NEET.

The acceptability findings and the qualitative evidence highlighted many aspects of the interventions that young NEET people have reported to be helpful. This included: the emotional and physical support they received from those delivering the intervention, having a place to share and discuss emotions; and being able to engage with animals or nature.

While the review sought to identify interventions that could be delivered in a nonclinical, community-based, or home setting, the majority of interventions required a number of resources. This included the availability of specific locations such as the nature or animal-assisted interventions, and requiring qualified staff such as therapists, social workers, social prescribers, child and youth care workers or mindfulness instructors, which may impact the applicability and feasibility of the interventions. A multi-agency approach to addressing the complex needs of NEET youth and promoting their long-term wellbeing may be required.

Some challenges were also reported from those delivering the social prescribing intervention (Bertotti et al., 2025) such as not having the time to support participants and continue their professional working responsibilities or not feeling experienced enough to support participants who may have experienced extreme isolation or are highly mentally vulnerable. Further interventions to support the mental and emotional health and wellbeing of young people who are NEET should ensure dedicated adequate time is provided to those delivering the intervention and that all those providing support are adequately trained to support the participants.

### 3.5 Implications for future research

A limited number of studies specifically looking to support the mental and emotional health and wellbeing of young people who are NEET were identified. None of the evidence identified reported any long-term findings. Further research is needed with robust evaluation to assess the long-term effects of community-based interventions to support the mental and emotional health and wellbeing of young people who are NEET. High-quality research is also needed to identify which interventions would be most effective and for who, as those considered NEET may have varying mental health needs. Further research could be conducted based on some of the promising findings identified in this review. However, careful consideration should be given to the resources needed to deliver the interventions and the plausibility of delivering them on a larger scale.

Through screening it became clear that the majority of evidence available in this population was focused on interventions that aim to facilitate re-engagement to education, training or work. However, young people who are NEET and suffering with poor mental health and wellbeing may be less inclined to re-engage or even to attend re-engagement interventions. Further research assessing interventions to support the mental and emotional health and wellbeing of young people who are NEET is needed to ensure that those whose mental health and wellbeing may be acting as a barrier to re-engagement are not left behind.

## 4. ECONOMIC CONSIDERATIONS

### This section has been completed by the Centre for Health Economics & Medicines Evaluation (CHEME), Bangor University

- Future research assessing interventions to support the mental and emotional health and wellbeing of young people who are NEET should consider the economic impacts of intervention delivery and wider implications concerning entry to employment, education or training facilitated by the scheme. Evaluations should consider both NHS and wider societal perspectives to capture all outcomes associated with re-entering employment, education or training.
- Young people who are NEET possess economic potential if they are to transition to employment, education or training. These economic gains may be manifest in economic growth and productivity through employment, as well as wider spillover effects related to improvements in individual mental and emotional health and wellbeing (Prince’s Trust, 2022).
- Integrating one young person who is NEET into the workforce could increase UK Gross Domestic Product (GDP) by over £74,000 per year*. This increase in GDP is defined by output per worker per year (PWC, 2024).
- If the proportion of young people who are NEET in Wales (currently 13%) were to reach 7.8%, annual economic gains of around £1.6bn* could be achieved (PWC, 2024). The figure of 7.8% in the analysis represents the UK region with the lowest rate of young people who are NEET, the Southwest of England (7.8%).

*figures inflated to 2025 prices using Bank of England Inflation Calculator: https://www.bankofengland.co.uk/monetary-policy/inflation/inflation-calculator

## Abbreviations

Acronym: Full Description
AAT: Animal assisted therapy
BPS: Basic package of support
CGAS: Children’s Global Assessment Scale
COP: Community of Practice
COPE: Capabilities, Opportunities, Places, and Engagement
CORE-OM: Clinical Outcomes in Routine Evaluation - Outcome Measurement
DERS: Difficulties in Emotion Regulation Scale
DERS: Difficulties in Emotion Regulation Scale
GAD: Generalised Anxiety Disorder
GAF: Global Assessment Functioning sale
IAP: Individual Action Plan
IAS: Interaction Anxiousness Scale
JBI: Joanna Briggs Institute
MMAT: Mixed methods appraisal tool
NEET: Not in education, employment or training
NGYCP: The National Guard Youth ChalleNGe Program
PESES: Perceived Employability Self-Efficacy Scale
PLR: Preliminary literature review
RCT: Randomised controlled trial
RM: Regain Momentum
RR: Rapid review
RSES: Rosenberg Self-Esteem Scale
SBOF: Social Behaviour Observation Form
SWEMWBS: Short Warwick-Edinburgh Mental Wellbeing Scale
SWY: Socially Withdrawn Youth
UK: United Kingdom
USA: United States of America
WAI: Working Alliance Inventory
WSAEMWB: Whole school approach to emotional and mental wellbeing
YDU: Youth Development Unit

## Data Availability

All data produced in the present study are available upon reasonable request to the authors

## 6. RAPID REVIEW METHODS

### 6.1 Eligibility criteria

#### REVIEW METHODS

Searches for primary sources were conducted to answer the review question: what is the effectiveness of interventions to support mental and emotional health and wellbeing in young people who are NEET. And sub-question: What is the acceptability of interventions to support the mental and emotional health and wellbeing in young people who are NEET?

**Table 3.** Eligibility criteria.

|  | Inclusion criteria | Exclusion criteria |
| --- | --- | --- |
| **Participants** | Children and young people, aged between 16 to 24 years and who are not in education, employment or training (NEET).<br>Studies in which the majority of the sample are our population of interest. E.g. studies that include NEET populations aged 16-35 where the majority are aged 16-24.<br>Mixed populations that include young NEET people as the majority, and/or where a sub analysis provides findings for the NEET group separately.<br>Any sub-populations of young people who are NEET e.g. those living in rural communities, those with disabilities etc. | Children and young people who are in education, employment or training.<br>Adults aged over 24 years.<br>Young people who are NEET and are already engaging in mental or emotional, health and wellbeing interventions/treatment. |
| **Intervention / exposure** | Interventions aimed at supporting, maintaining, preventing or enhancing the mental and emotional health and wellbeing of young people who are NEET.<br>Delivered in a nonclinical, community-based, or home setting. | Interventions aimed at improving employability or re-engagement in education, training, or employment.<br>Interventions delivered in School, training centres, clinical settings or workplaces. |
| **Comparison** | Standard care, no intervention, before and after |  |
| **Outcomes** | Primary outcomes:<br>Any mental and emotional health and wellbeing outcome e.g. resilience, coping skills, anxiety etc<br>Mental health service use.<br>Secondary outcome:<br>Acceptability of interventions among the NEET population | Return to employment, education, or training<br>Reduction in NEETS |
| **Study design** | Any comparative study design that reports on quantitative or qualitative data. | Evidence synthesis, economic evaluations, case studies. |
| **Countries** | All |  |
| **Language of publication** | English | Non-English |
| **Publication date** | Published from 2000 onwards |  |
| **Publication type** | Published and preprint | Grey literature, conference abstracts, editorials, letters, commentaries |

### 6.2 Literature search

A search was conducted of the electronic bibliographic databases: Medline, Embase, Scopus, sociology collection, Google Scholar, CINAHL and PsycINFO. The search strategy used for Medline is available in appendix 1. A search for ongoing and completed trials was conducted in Clinicaltrials.gov, and WHO International Clinical Trials Registry Platform (ICTRP). A search for grey literature was conducted in WHO, The Kings Trust, The national institute of mental health, The AnnaFreud.org, and Children and Young People’s Mental Health (ChYMe) research collaboration (Exeter Uni) websites. Citation tracking from secondary sources identified during the preliminary stages was also undertaken. All searches were conducted between 5th and 12th June 2025. Search concepts and keywords included ‘not in education, employment or training’ NEET, mental health, emotional health, resilience, and wellbeing. Searches were limited to records published after 2000 to ensure the recency and relevancy of the included studies and were published in the English language.

### 6.3 Study selection process

Following removal of duplicates in Endnote, all articles were imported to a systematic reviewing software Rayyan for screening. For title and abstract screening, titles were screened by a single reviewer against the eligibility criteria in section 6.1. To ensure consistency, a proportion (just over 10%) of studies were screened by two independent reviewers. Full text screening was carried out in duplicate by two independent reviewers, and any conflicts were discussed and resolved within the team.

### 6.4 Data extraction

Data was extracted directly from the primary studies by one reviewer and a second reviewer consistency checked the data extracted for accuracy and completeness. The information extracted included:

- Reference (author, year, country)
- Study design
- Study aim
- Sample Size
- Data collection methods and dates
- Study participants characteristics (e.g. number of participants, age, sex/gender, vulnerability category, ethnicity, disadvantage group, participants from deprived areas, protected characteristics)
- Setting
- Intervention details (including type of intervention/duration/delivery method/setting)
- Comparator intervention/ control
- Outcomes
- Findings
- Observations/Notes

### 6.5 Study design classification

The included studies were classified as RCT’s, non-randomised controlled studies, and uncontrolled before and after studies (three of which utilised mixed methods). A study classification algorithm was not necessary for this rapid review.

### 6.6 Quality appraisal

Quality assessment was undertaken in duplicate by two independent reviewers. Any discrepancies were discussed and resolved between reviewers. Comments on the quality were recorded and discussed in the analysis. The quality assessment of individual studies can be seen in section 7.3.

The JBI critical appraisal checklists for quasi-experimental studies (Barker et al., 2024) and RCTs were used (Barker et al., 2023, and the mixed methods appraisal tool (Hong et al., 2018) for mixed methods were used to assess the methodological quality of each included study. These checklists are not designed to assign an overall score to each study. For the purposes of this review, a pragmatic system devised and used in a previous rapid review (Weightman et al., 2022), was used to assess each study as being of high, moderate, or low quality for the quasi-experimental studies and the RCT.

*High quality:* RCT plus a score of 8 or more YES on the JBI RCT check list, plus, no concerns suggesting a medium or high risk of bias in any aspect of the study design.

*Moderate quality:* RCT plus a score of 6 or more YES on the JBI RCT check list OR a score of 8 or more YES on the JBI quasi-experimental study checklist, plus, no concerns suggesting a very high risk of bias in any aspect of the study design.

*Low quality*: Others

To determine the overall quality of the mixed method studies a similar approach was used. Each component (quantitative, qualitative and mixed methods) was scored out of 5, following the guidance from the MMAT the overall quality cannot exceed the quality of its weakest component (Hong et al., 2018). For the purposes of this review:

High quality: RCT with a score of 4 or more YES on the MMAT for each component, plus, no concerns suggesting a medium or high risk of bias in any aspect of the study design.

Moderate quality: RCT with a score of 3 YES on the MMAT for each component OR a score of 4 or more YES on the MMAT for each component, plus, no concerns suggesting a medium or high risk of bias in any aspect of the study design.

Low quality: Others

### 6.7 Synthesis

Data was synthesised narratively to provide a collective interpretation of the evidence. An assessment of the body of evidence and any limitations were discussed and conclusions provided.

### 6.8 Assessment of body of evidence

To assess the overall body of evidence considerations were made based on the relevance of the available evidence, the amount and quality of the evidence, the direction of effect, and consistency in the findings. This information is provided for the differing interventions in Table 2.

## 7. EVIDENCE

### 7.1 Search results and study selection

A visual representation of the flow of studies throughout the review can be found in Figure 1. A total of 16,140 records were retrieved via the searches and 9,880 records remained following deduplication. A total of 304 articles were screened at full text and nine studies were included in the rapid review.

**Figure 1.**
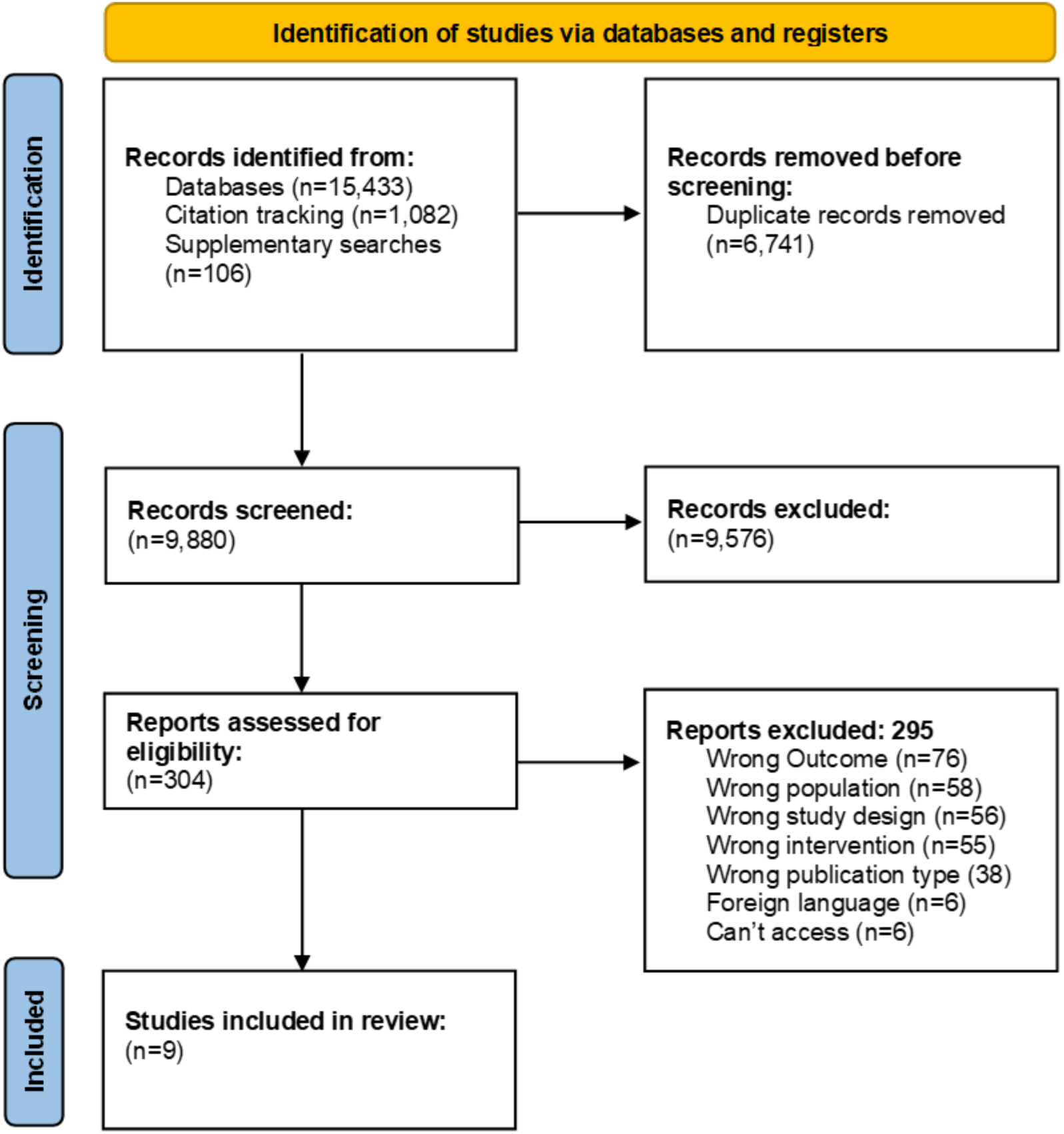
**PRISMA flow diagram**

### 7.2 Data extraction

**Table 4.**
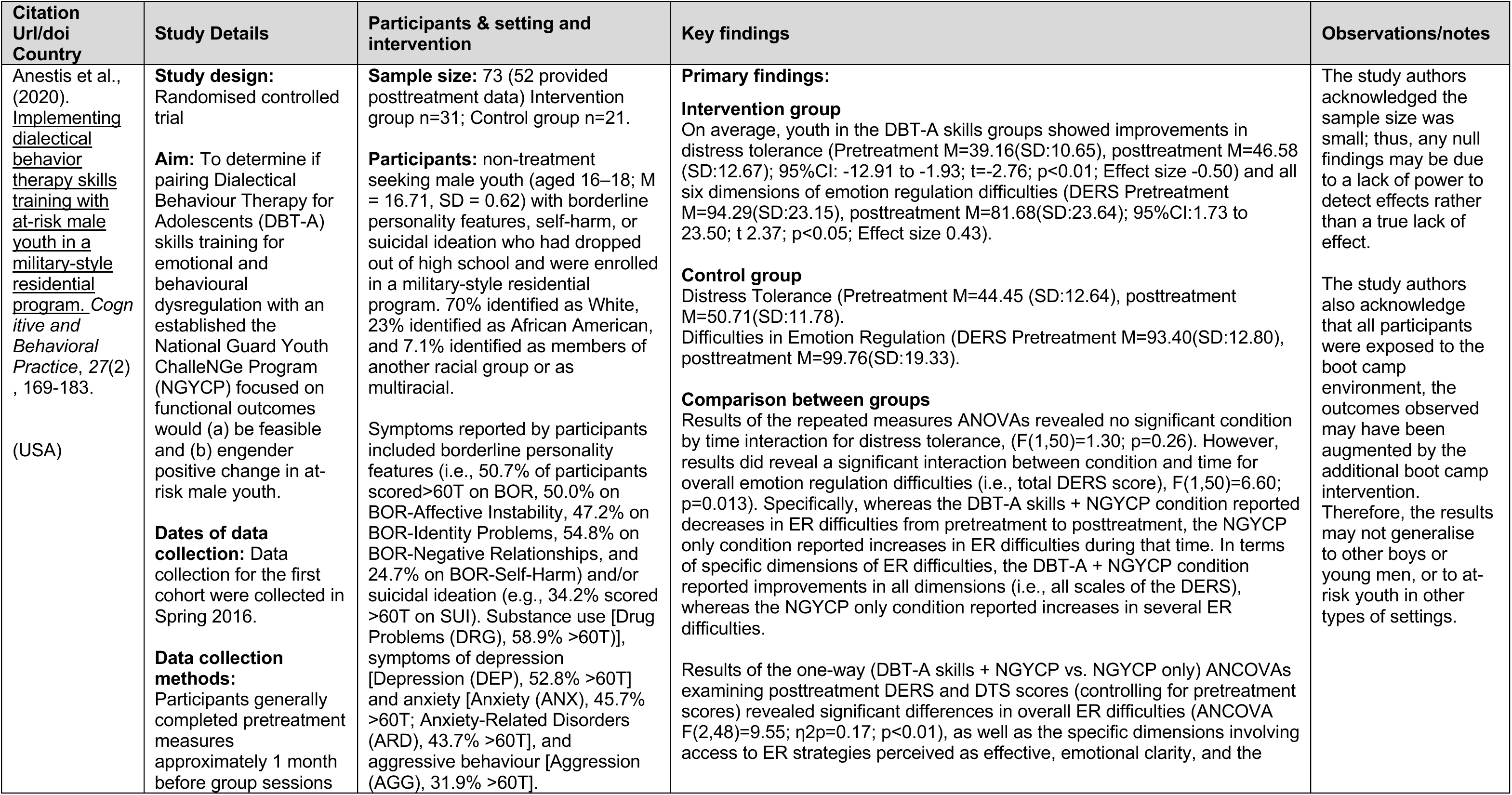
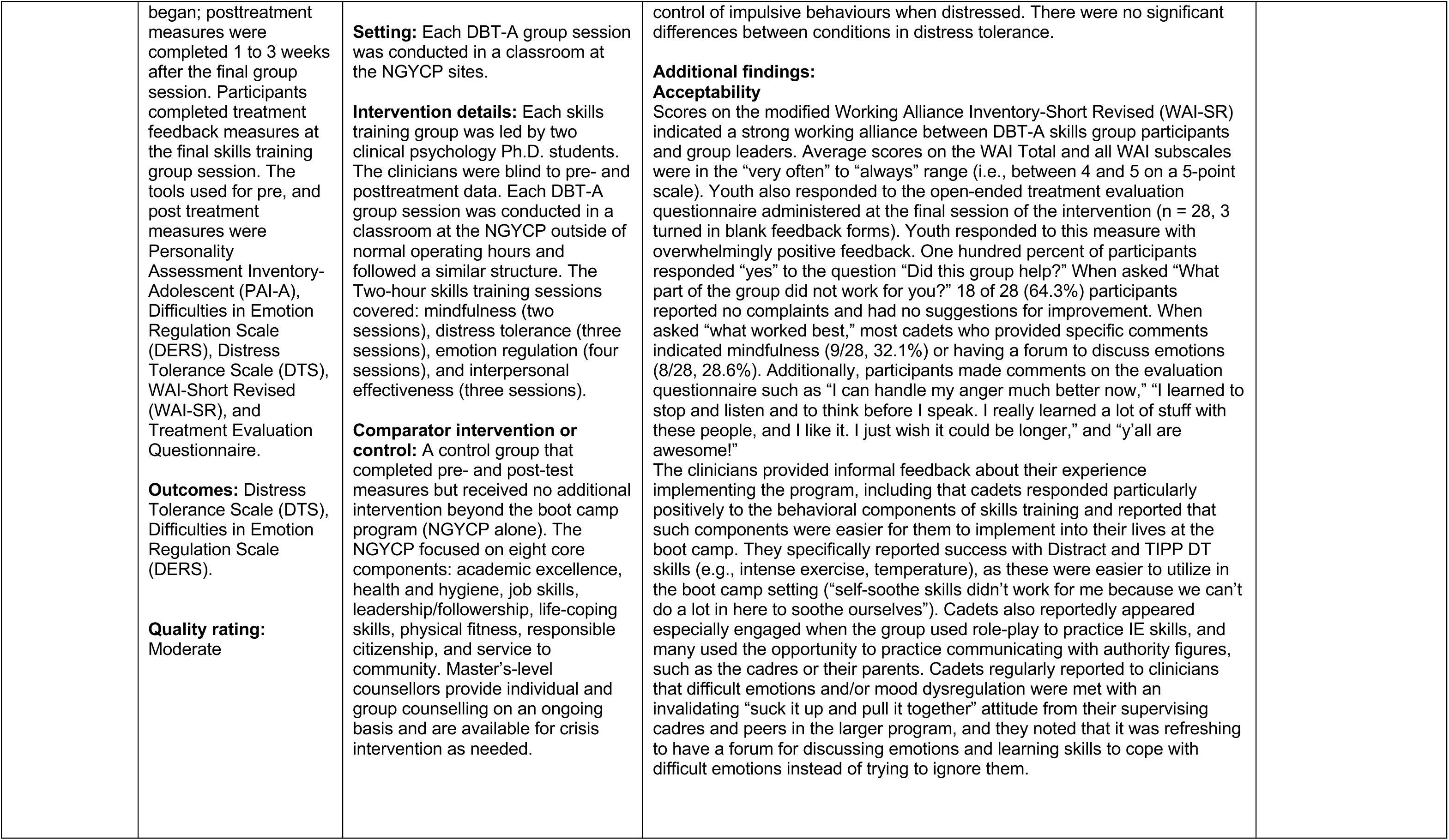
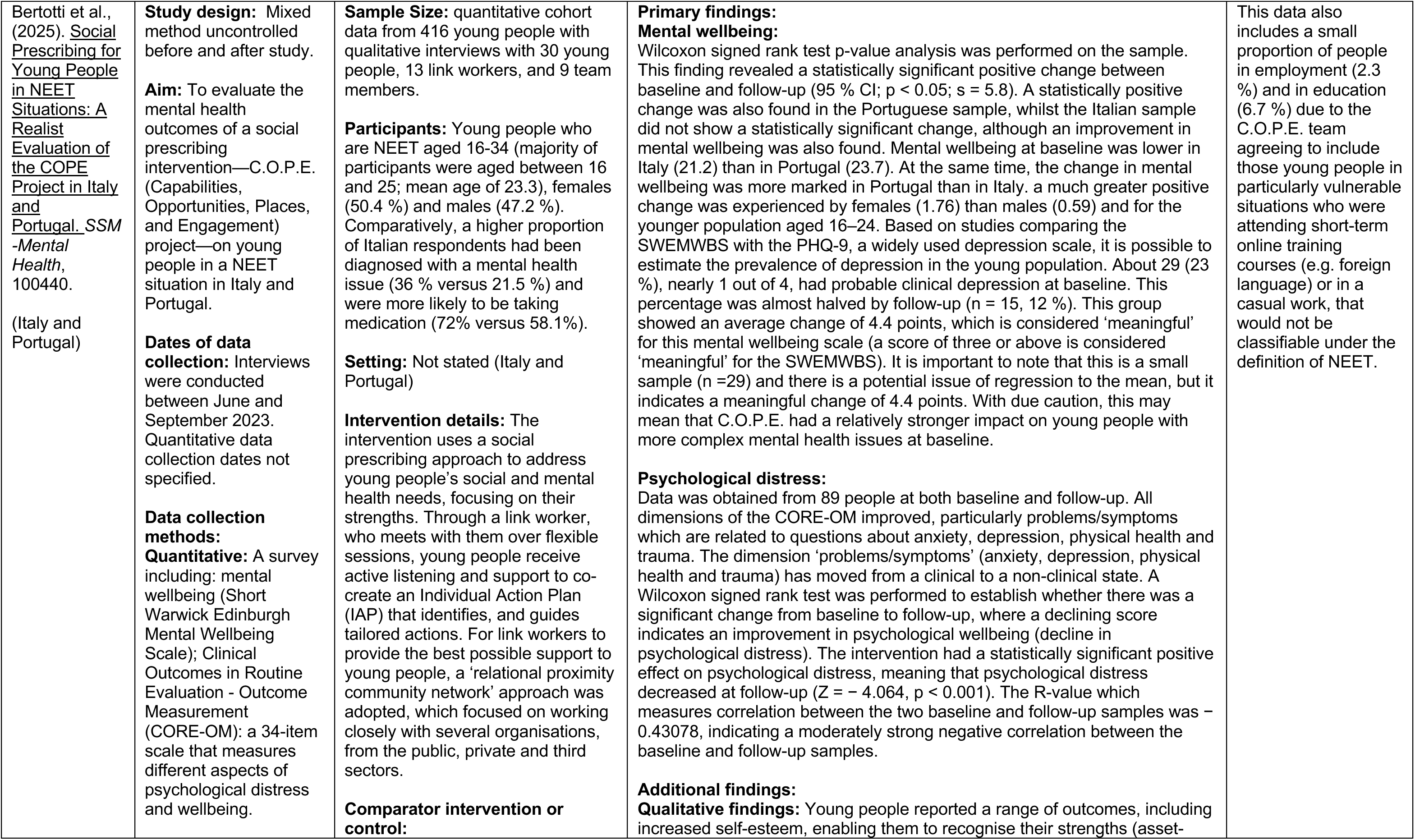
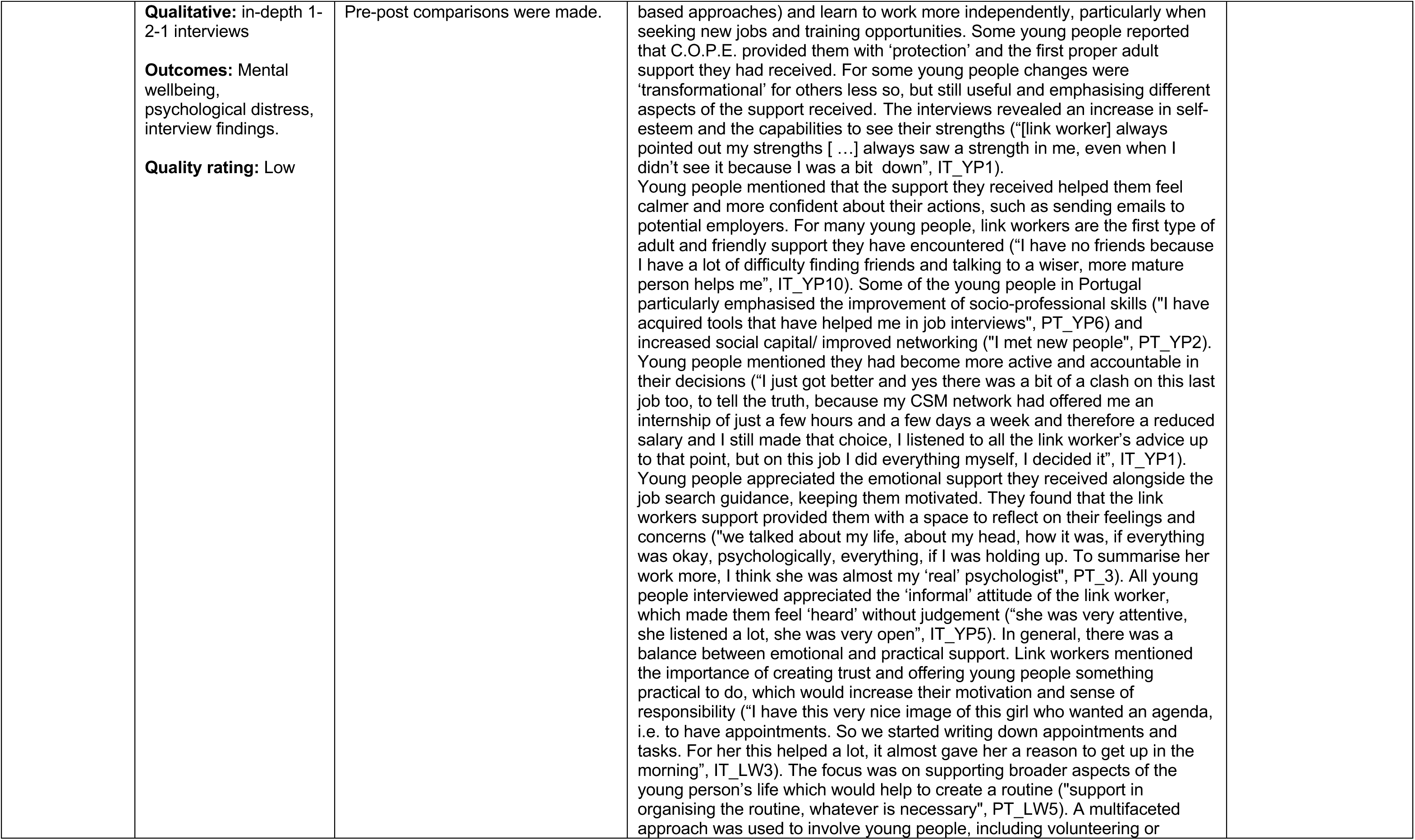
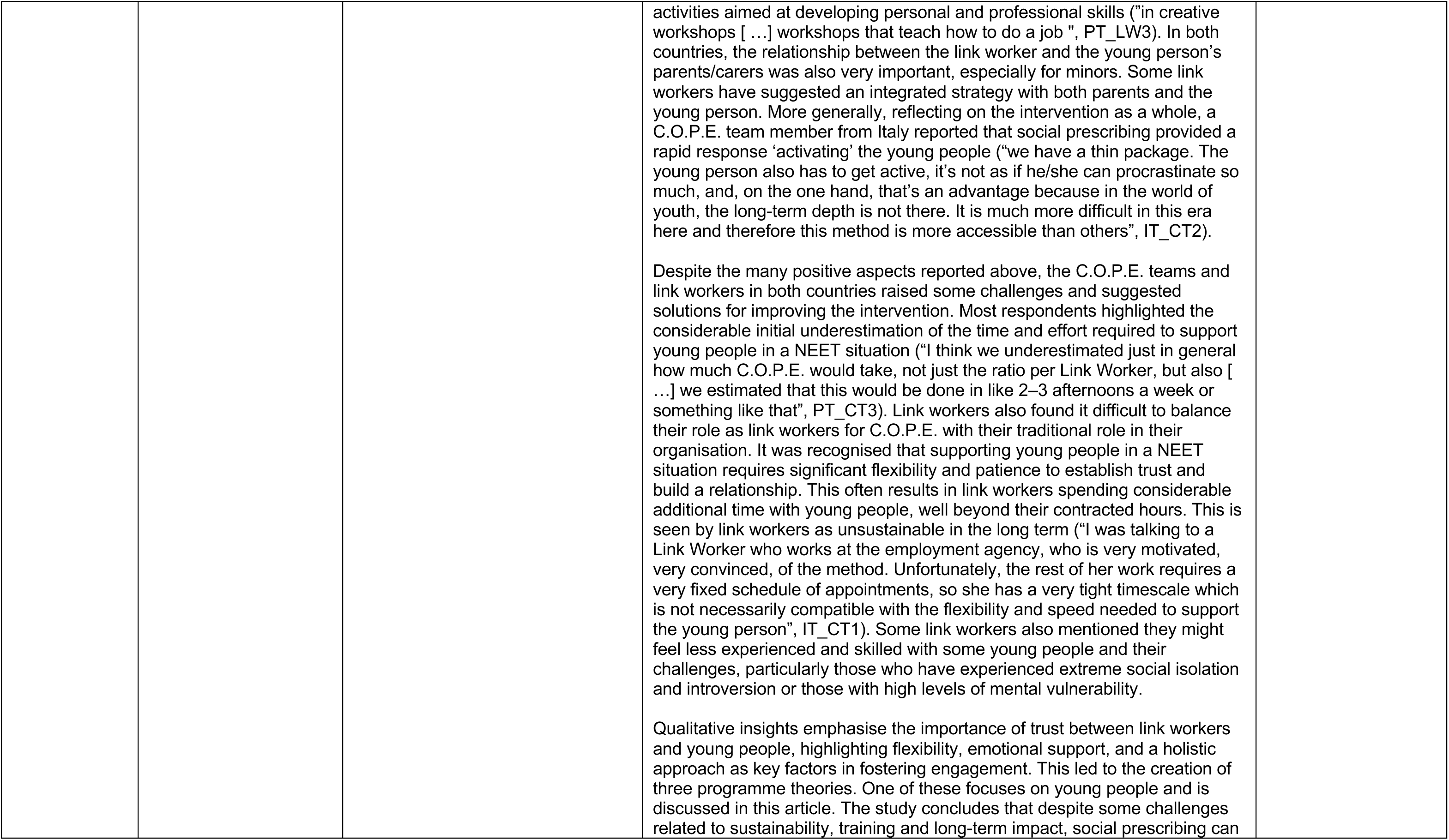
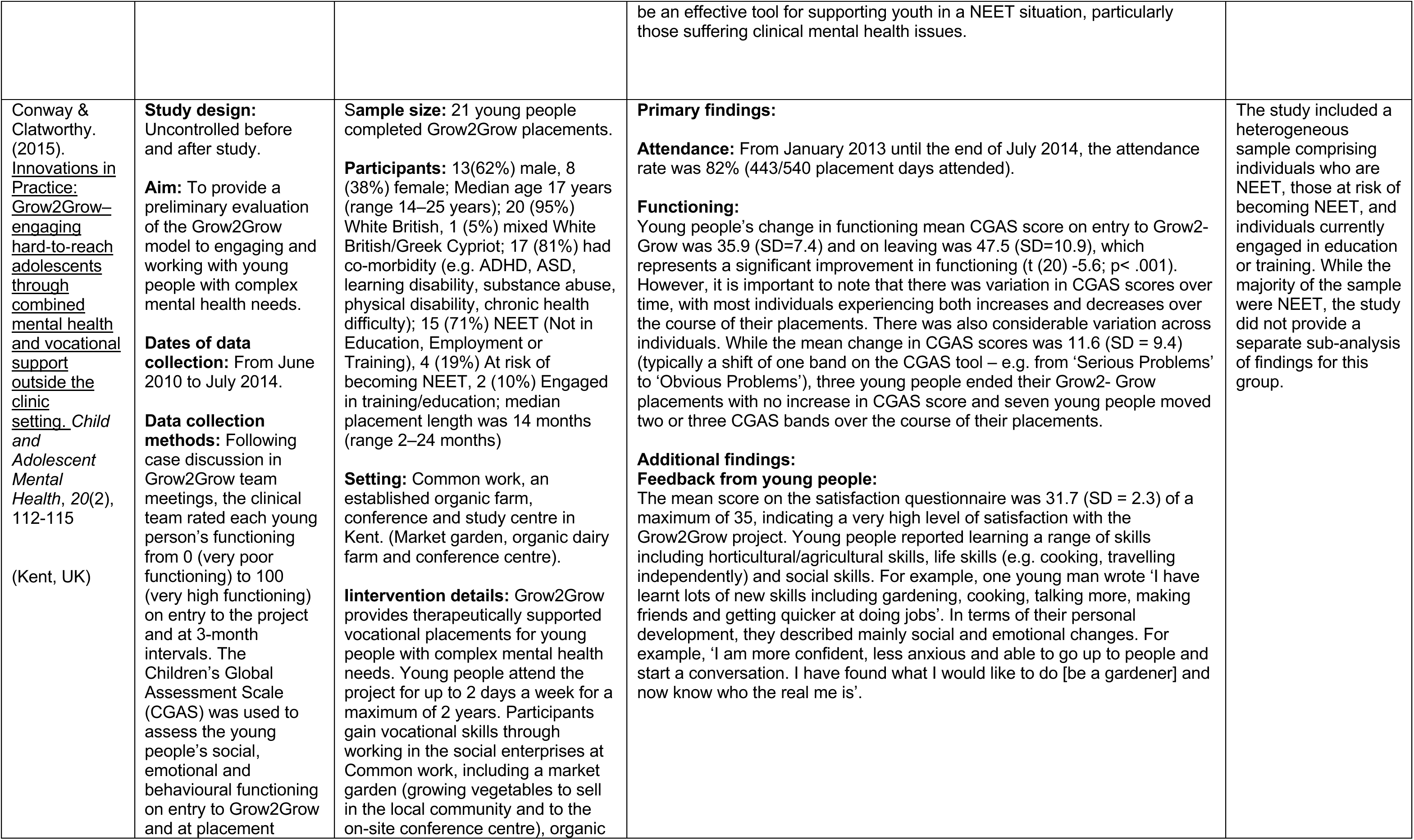
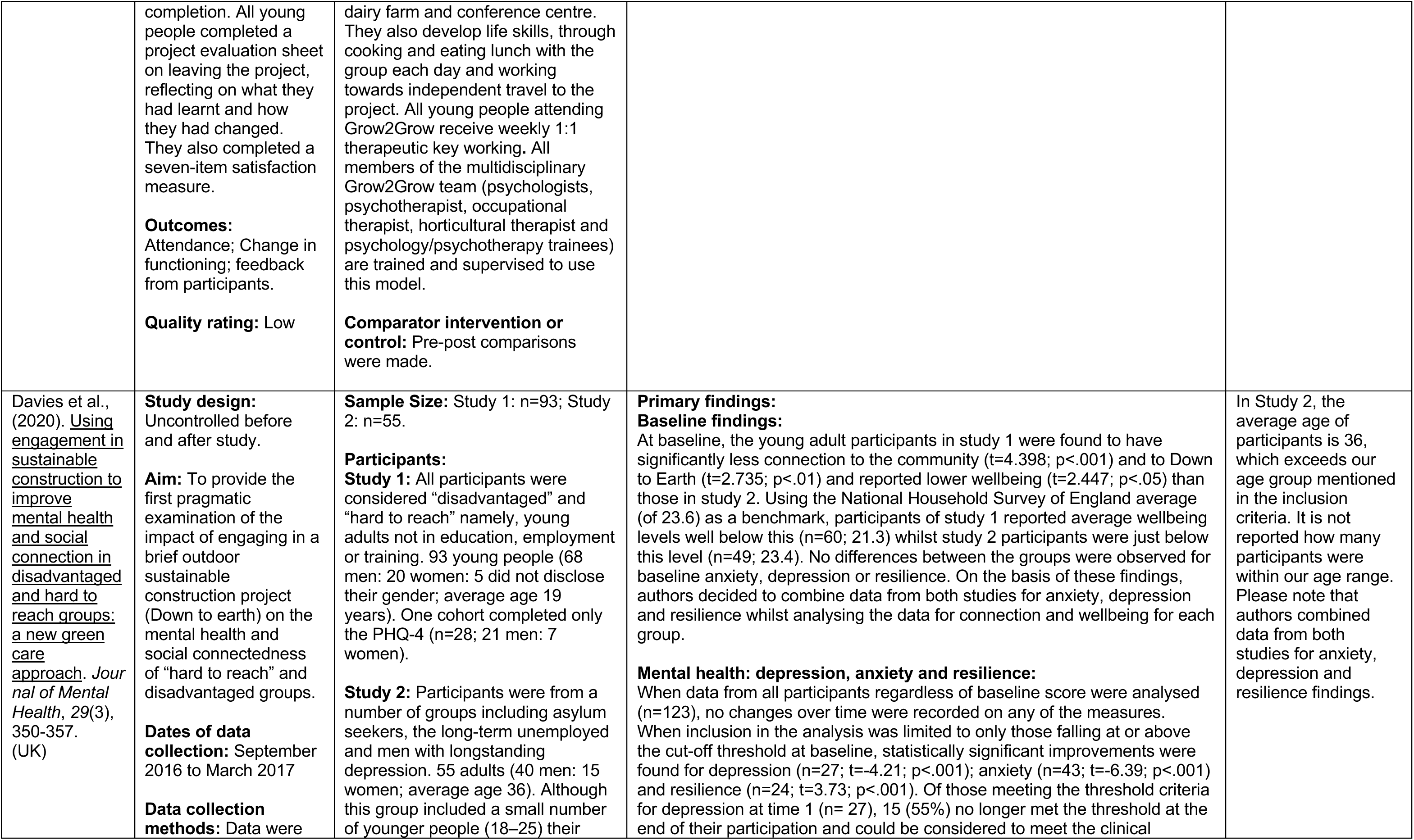
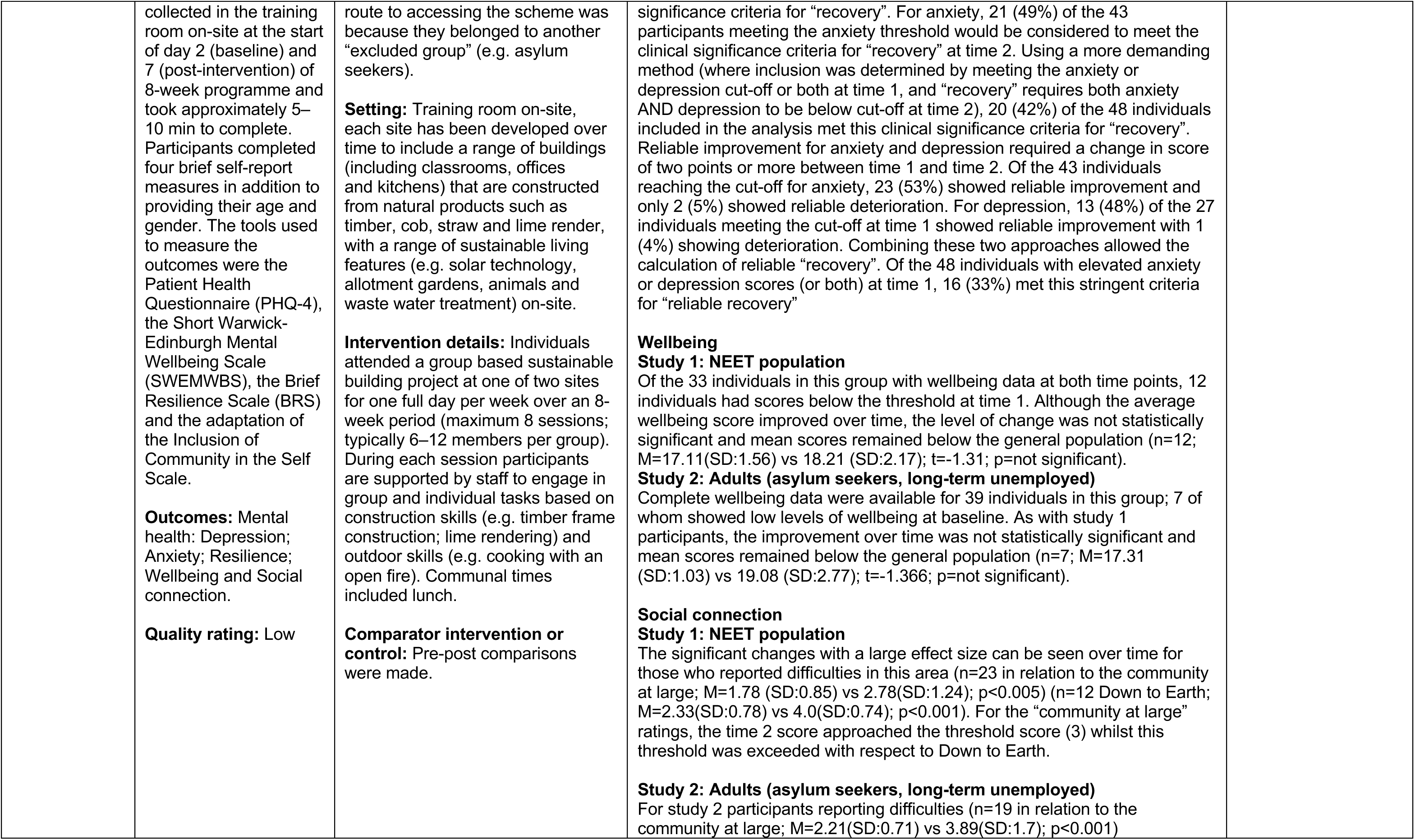
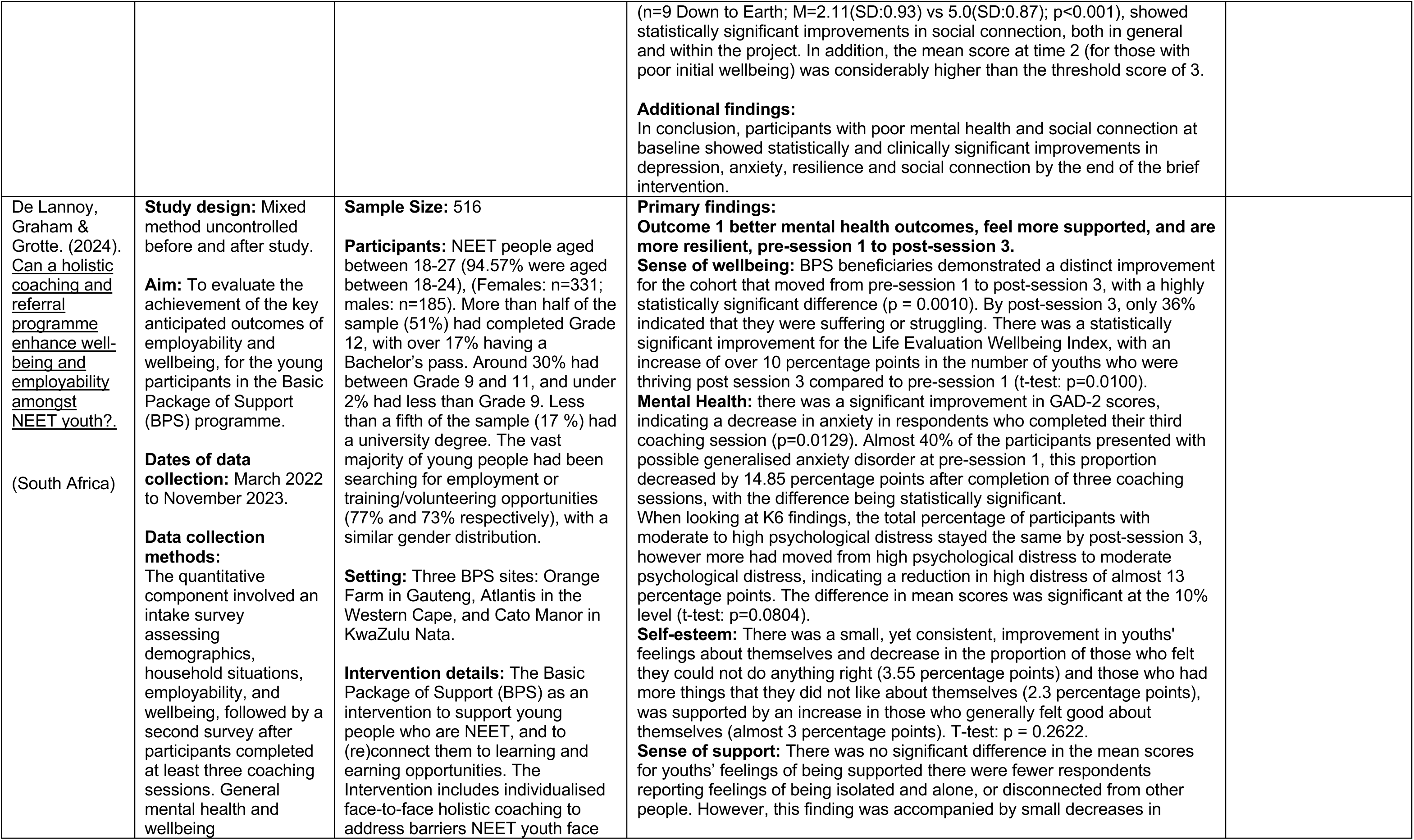
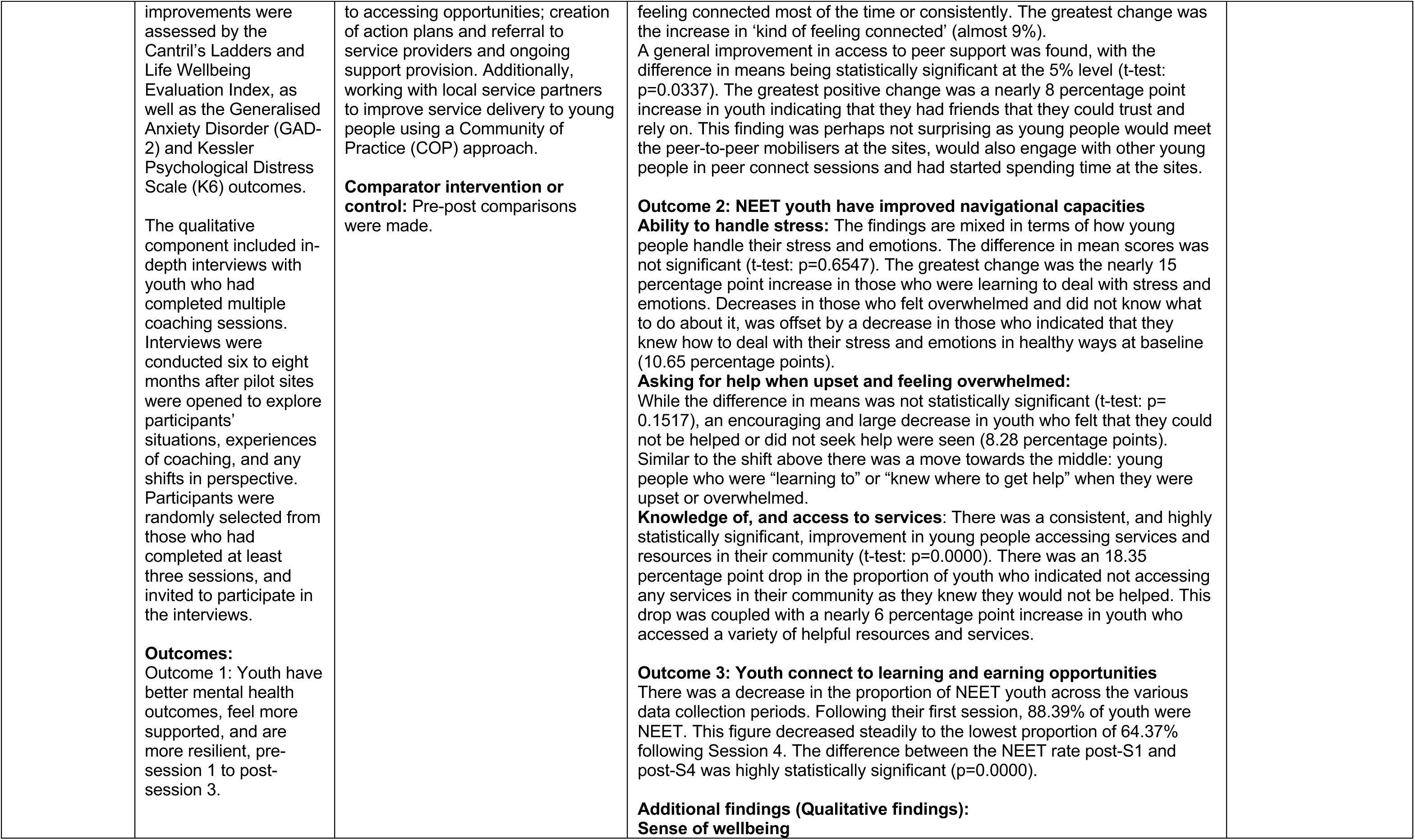
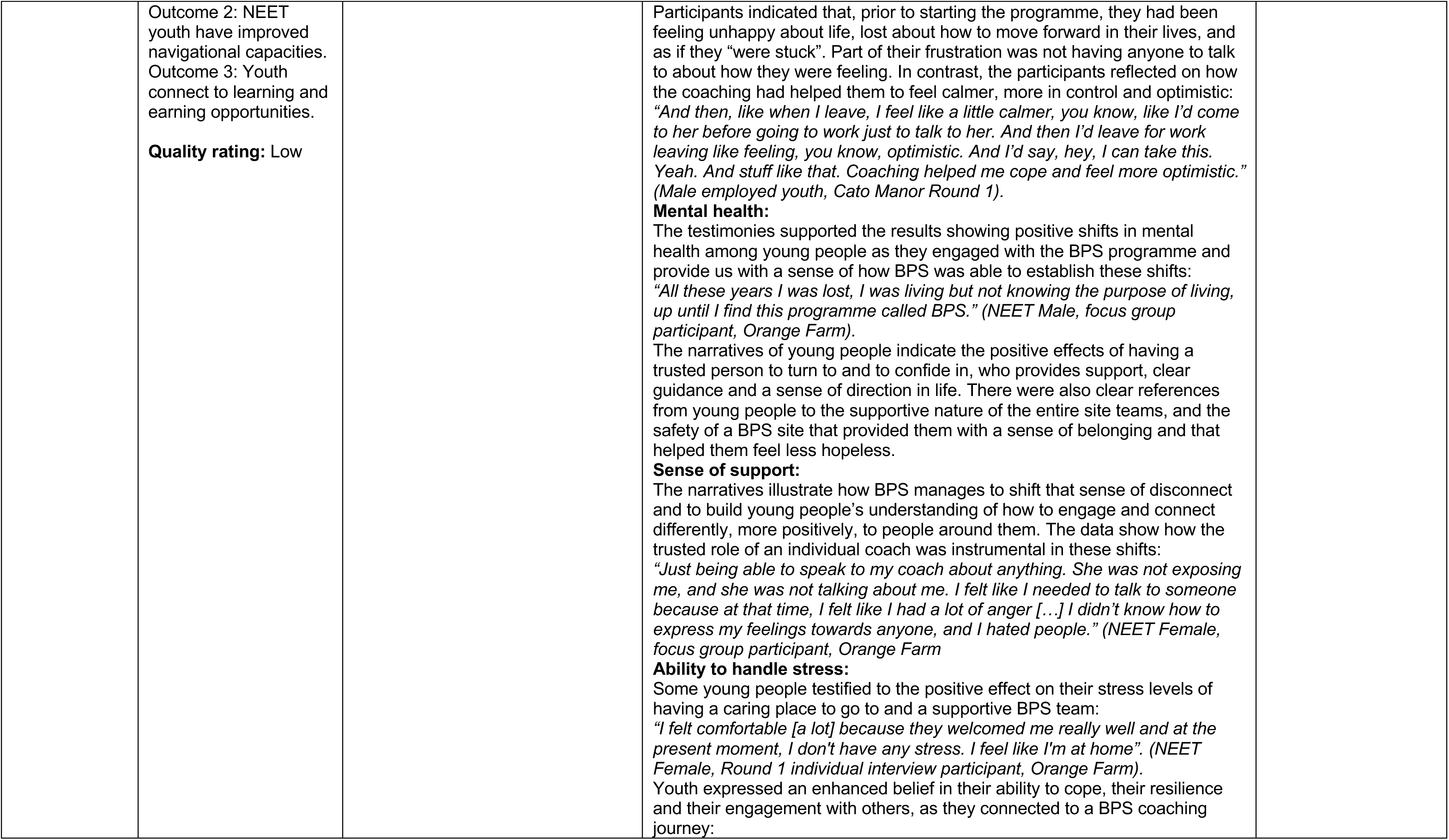
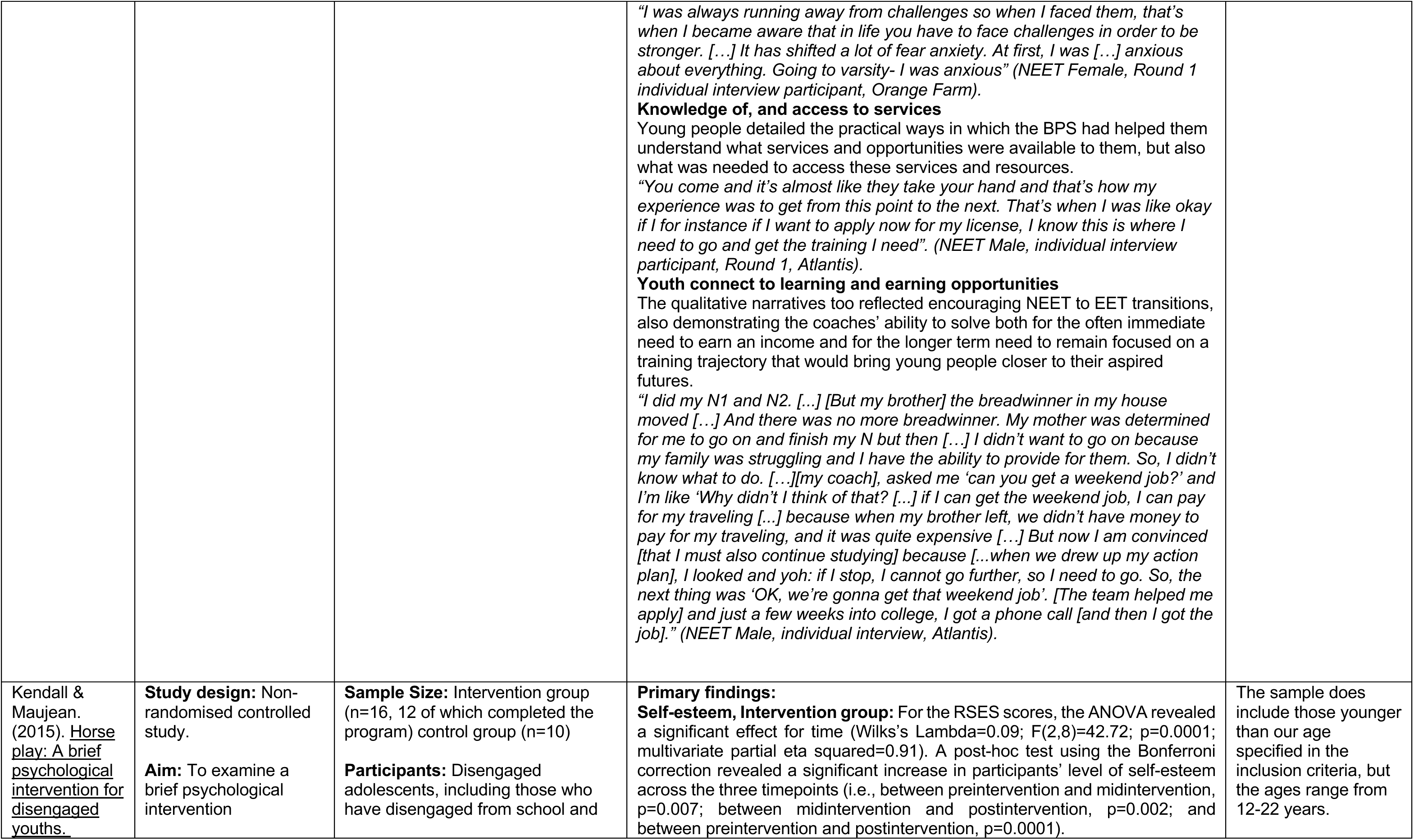
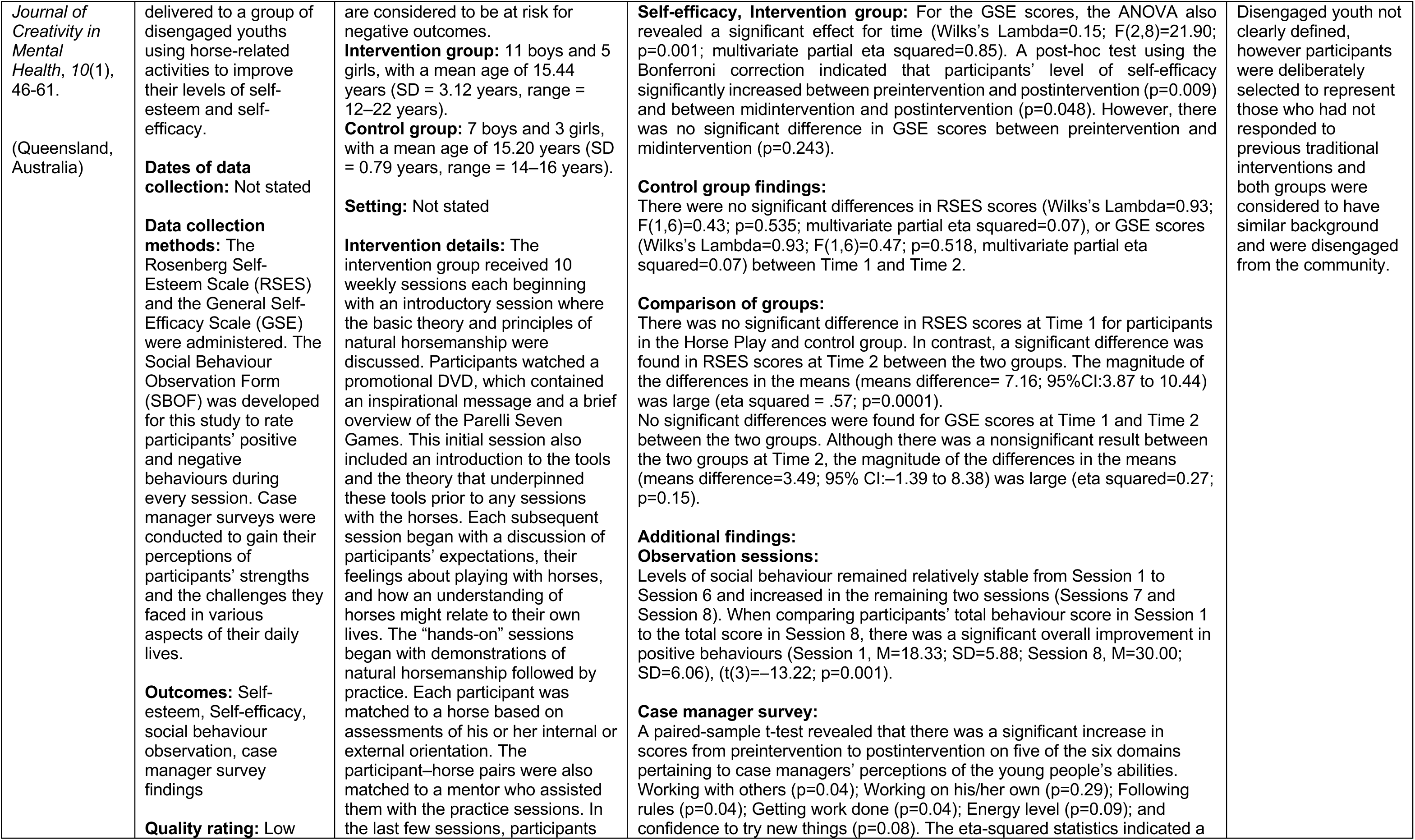
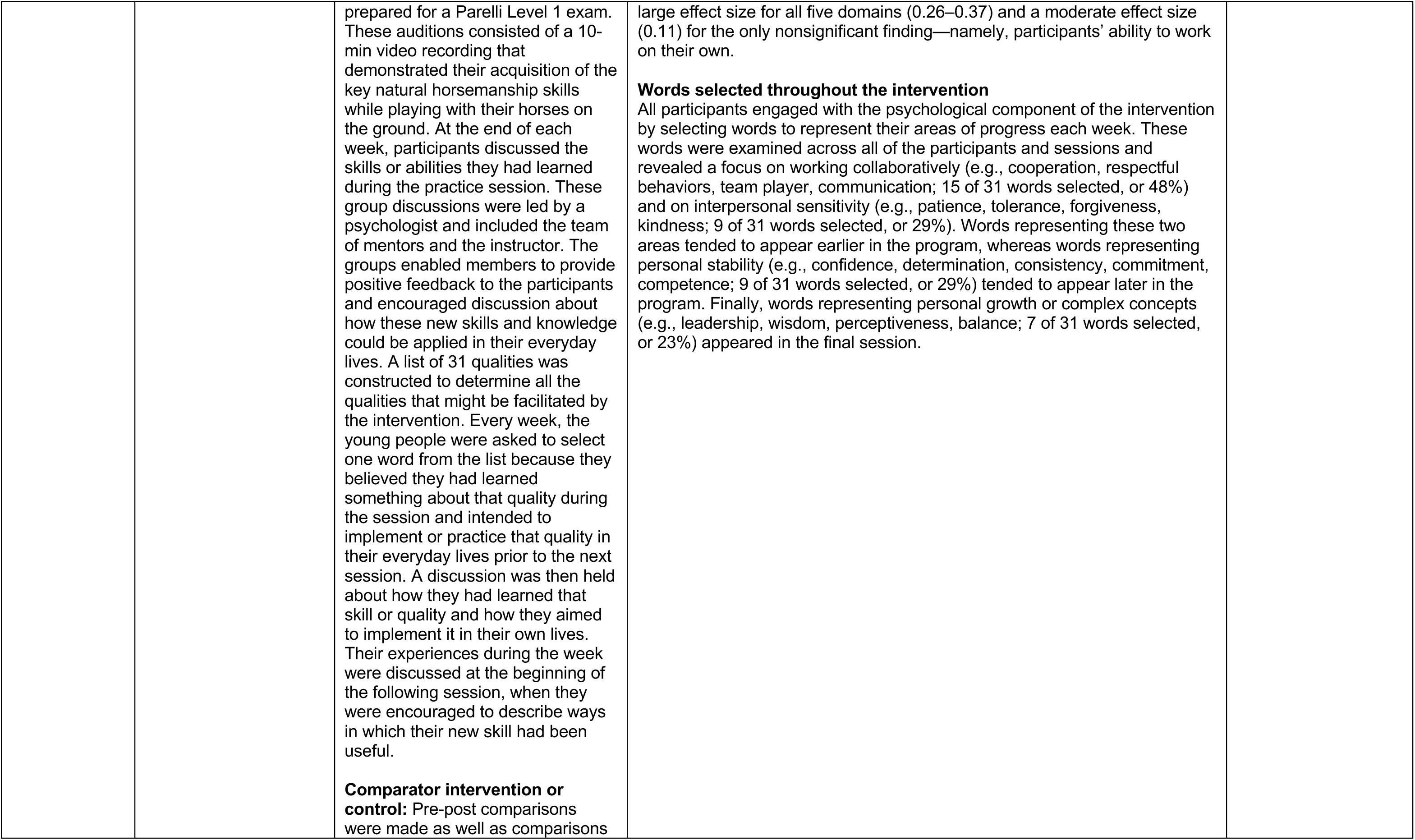
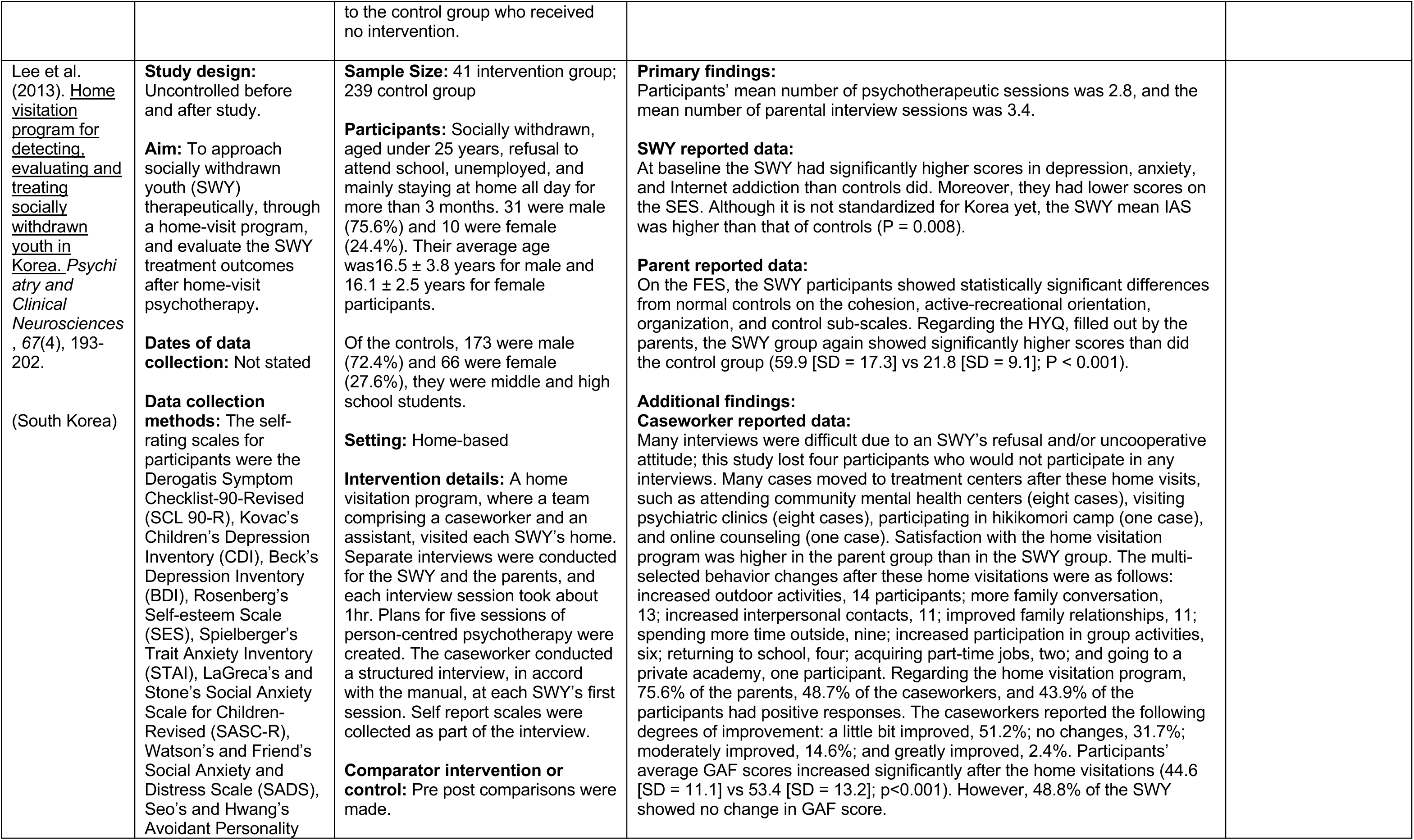
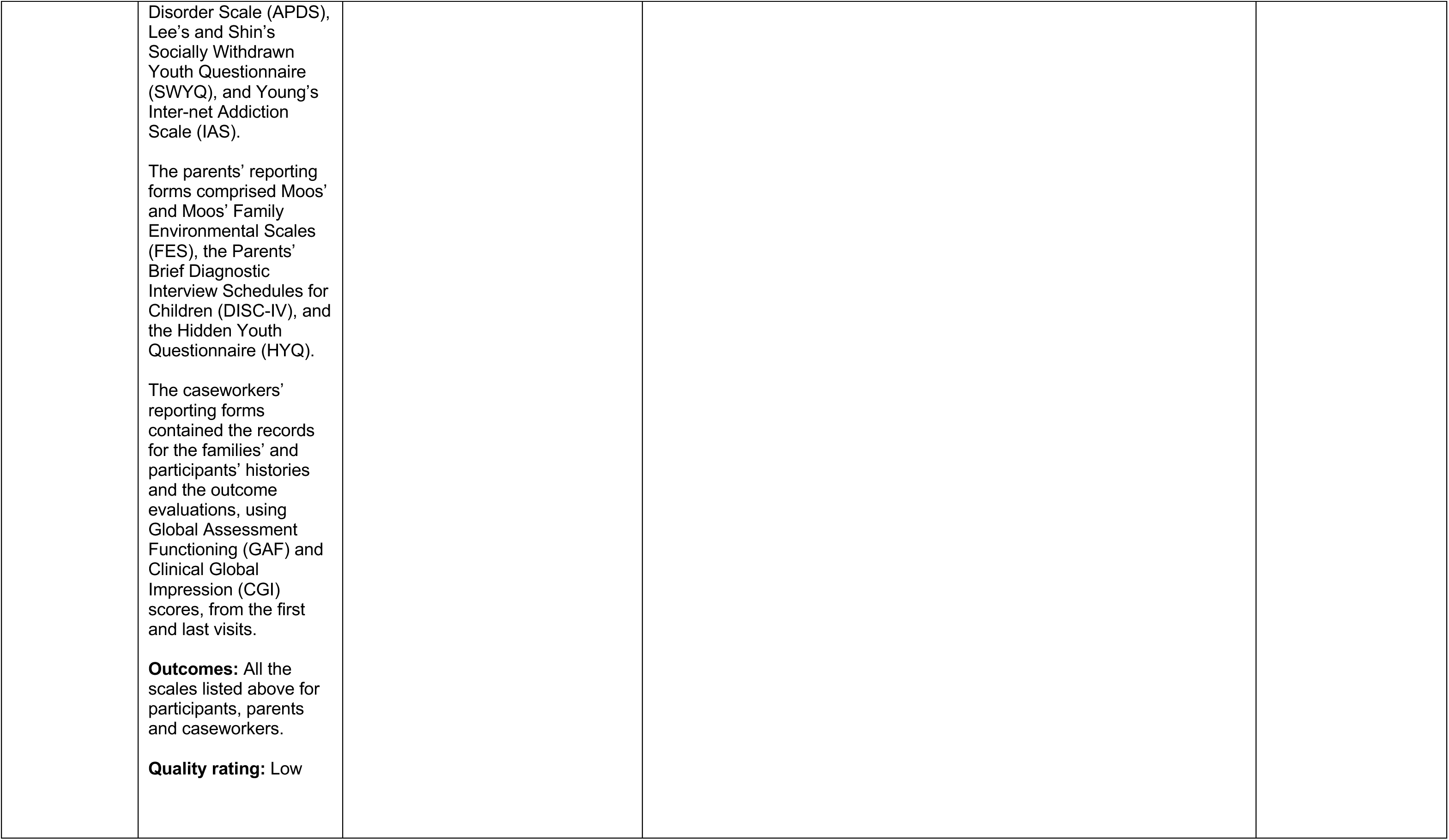
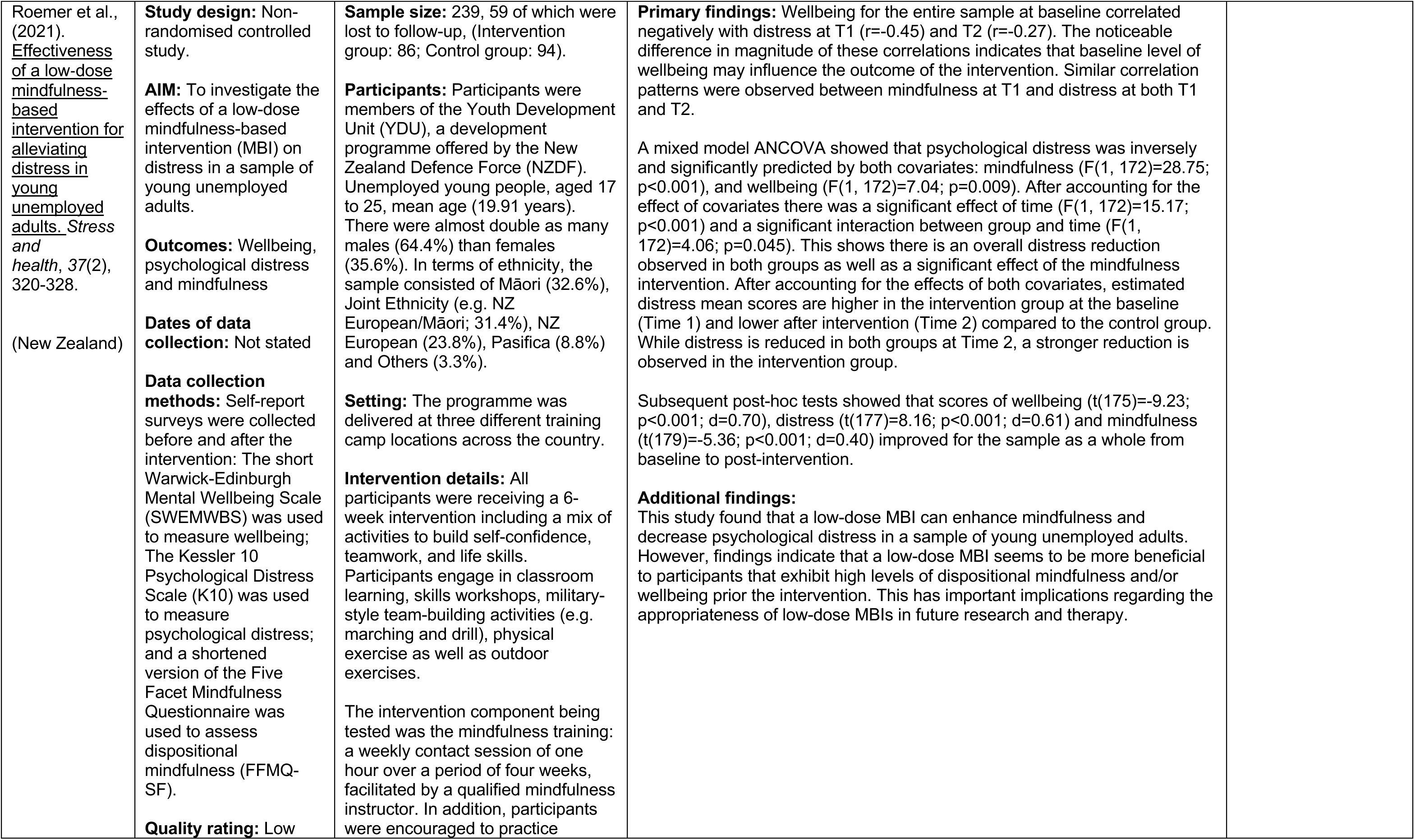
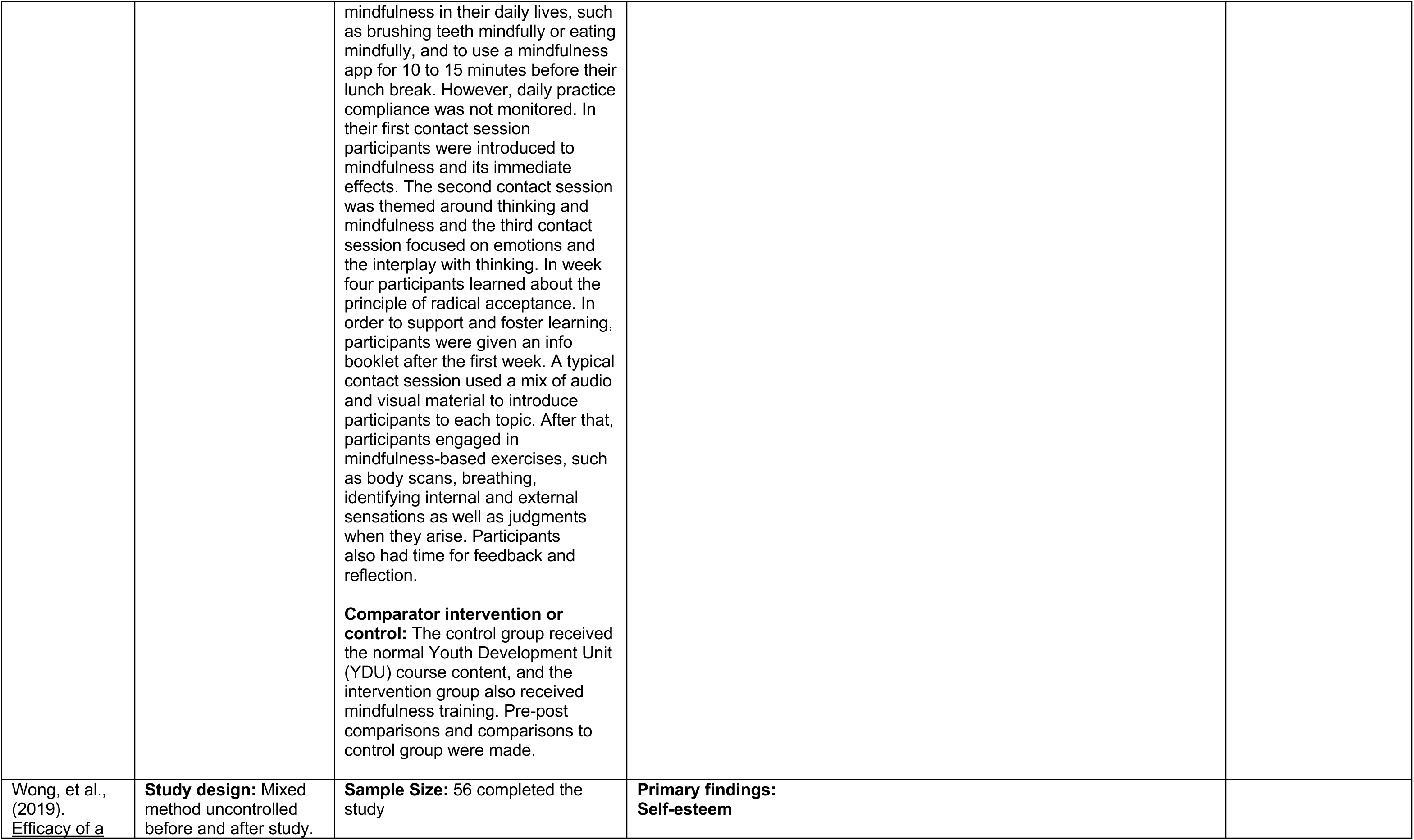
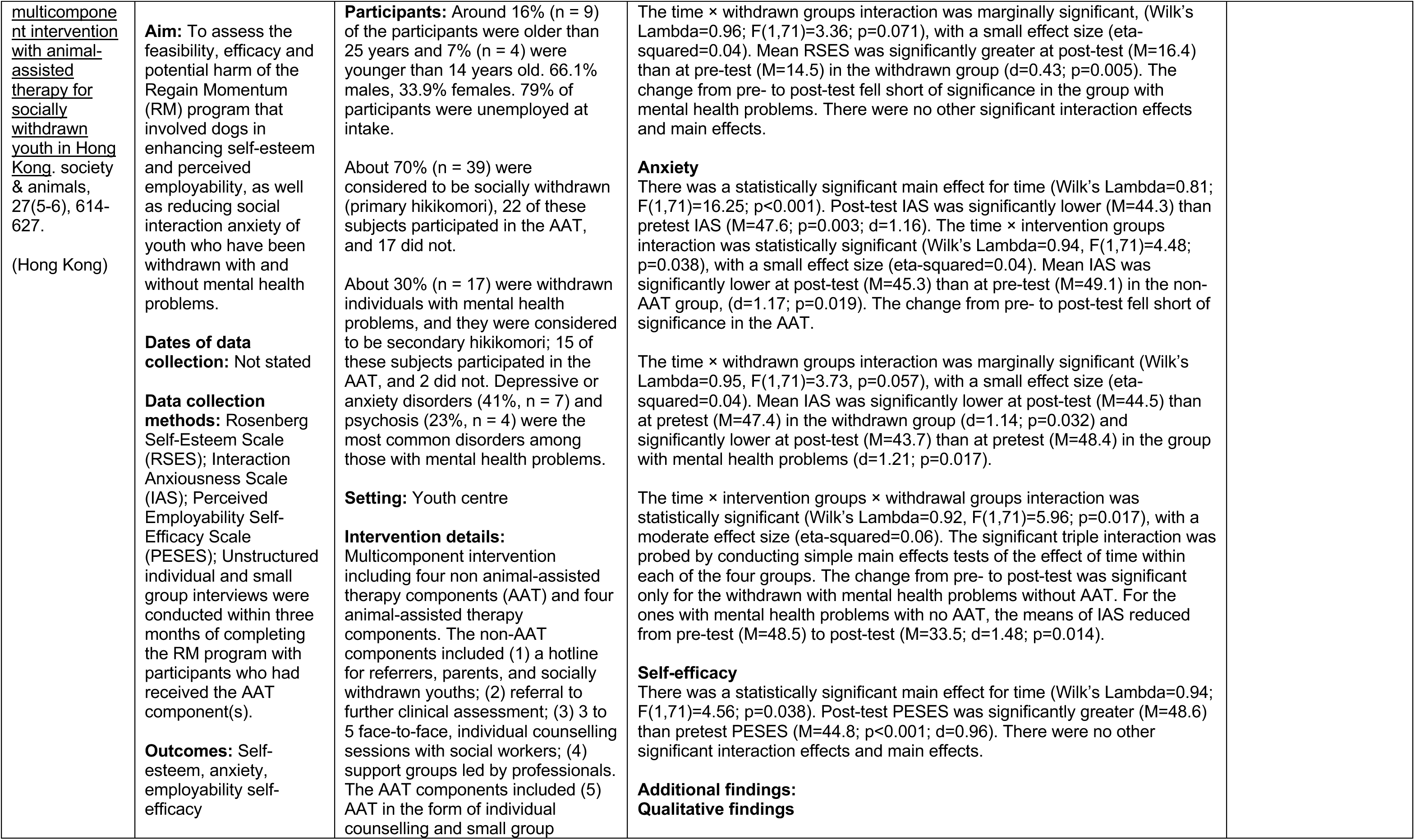
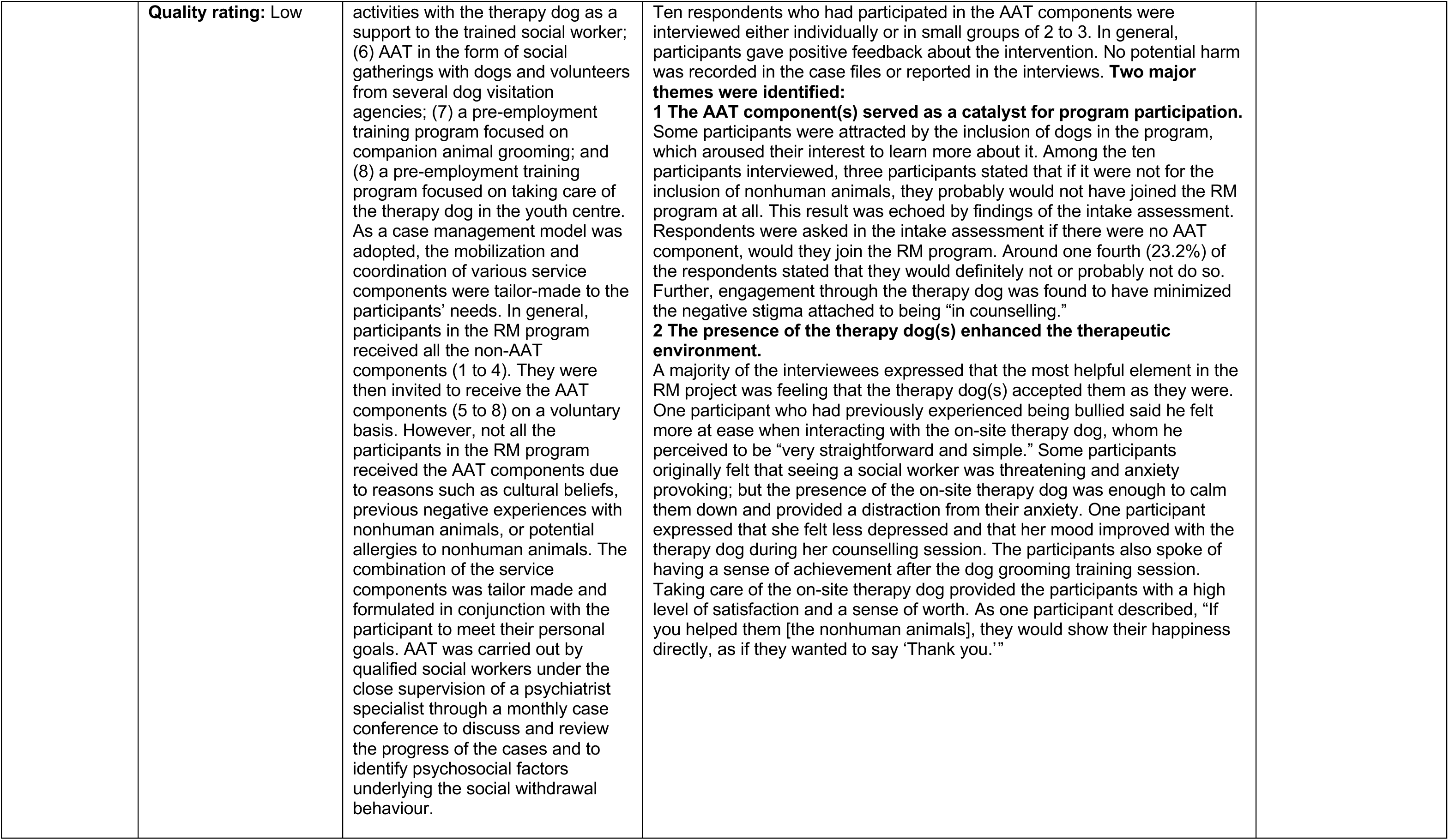
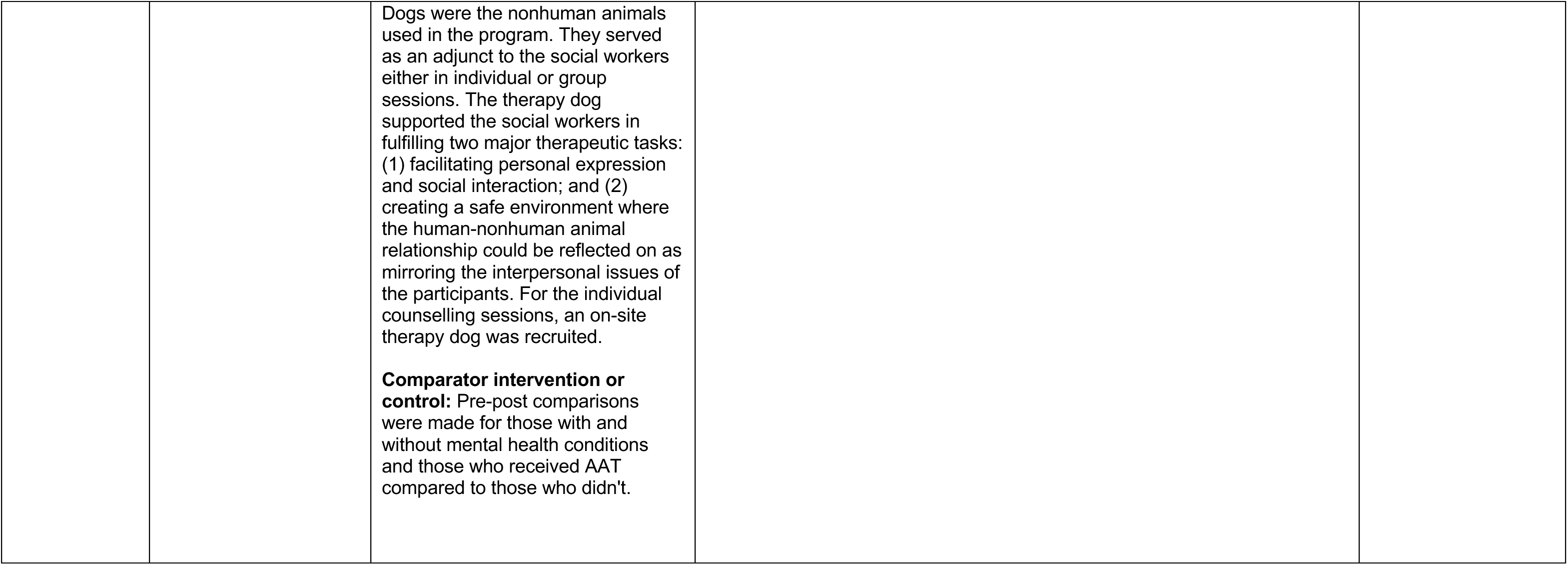
Summary of included studies.

| Citation<br>Url/doi<br>Country | Study Details | Participants & setting and<br>intervention | Key findings | Observations/notes |
| --- | --- | --- | --- | --- |
| Anestis et al., (2020).<br>Implementing dialectical behavior therapy skills training with at-risk male youth in a military-style residential program. *Cognitive and Behavioral Practice*, 27(2), 169-183.<br>(USA) | **Study design:** Randomised controlled trial<br>**Aim:** To determine if pairing Dialectical Behaviour Therapy for Adolescents (DBT-A) skills training for emotional and behavioural dysregulation with an established the National Guard Youth ChalleNGe Program (NGYCP) focused on functional outcomes would (a) be feasible and (b) engender positive change in at-risk male youth.<br>**Dates of data collection:** Data collection for the first cohort were collected in Spring 2016.<br>**Data collection methods:** Participants generally completed pretreatment measures approximately 1 month before group sessions | **Sample size:** 73 (52 provided posttreatment data) Intervention group n=31; Control group n=21.<br>**Participants:** non-treatment seeking male youth (aged 16–18; M = 16.71, SD = 0.62) with borderline personality features, self-harm, or suicidal ideation who had dropped out of high school and were enrolled in a military-style residential program. 70% identified as White, 23% identified as African American, and 7.1% identified as members of another racial group or as multiracial.<br>Symptoms reported by participants included borderline personality features (i.e., 50.7% of participants scored >60T on BOR, 50.0% on BOR-Affective Instability, 47.2% on BOR-Identity Problems, 54.8% on BOR-Negative Relationships, and 24.7% on BOR-Self-Harm) and/or suicidal ideation (e.g., 34.2% scored >60T on SUI). Substance use [Drug Problems (DRG), 58.9% >60T], symptoms of depression [Depression (DEP), 52.8% >60T] and anxiety [Anxiety (ANX), 45.7% >60T; Anxiety-Related Disorders (ARD), 43.7% >60T], and aggressive behaviour [Aggression (AGG), 31.9% >60T]. | **Primary findings:**<br>**Intervention group**<br>On average, youth in the DBT-A skills groups showed improvements in distress tolerance (Pretreatment M=39.16(SD:10.65), posttreatment M=46.58 (SD:12.67); 95%CI: -12.91 to -1.93; t=-2.76; p<0.01; Effect size -0.50) and all six dimensions of emotion regulation difficulties (DERS Pretreatment M=94.29(SD:23.15), posttreatment M=81.68(SD:23.64); 95%CI:1.73 to 23.50; t 2.37; p<0.05; Effect size 0.43).<br>**Control group**<br>Distress Tolerance (Pretreatment M=44.45 (SD:12.64), posttreatment M=50.71(SD:11.78).<br>Difficulties in Emotion Regulation (DERS Pretreatment M=93.40(SD:12.80), posttreatment M=99.76(SD:19.33).<br>**Comparison between groups**<br>Results of the repeated measures ANOVAs revealed no significant condition by time interaction for distress tolerance, (F(1,50)=1.30; p=0.26). However, results did reveal a significant interaction between condition and time for overall emotion regulation difficulties (i.e., total DERS score), F(1,50)=6.60; p=0.013). Specifically, whereas the DBT-A skills + NGYCP condition reported decreases in ER difficulties from pretreatment to posttreatment, the NGYCP only condition reported increases in ER difficulties during that time. In terms of specific dimensions of ER difficulties, the DBT-A + NGYCP condition reported improvements in all dimensions (i.e., all scales of the DERS), whereas the NGYCP only condition reported increases in several ER difficulties.<br>Results of the one-way (DBT-A skills + NGYCP vs. NGYCP only) ANCOVAs examining posttreatment DERS and DTS scores (controlling for pretreatment scores) revealed significant differences in overall ER difficulties (ANCOVA F(2,48)=9.55; $\eta^2$p=0.17; p<0.01), as well as the specific dimensions involving access to ER strategies perceived as effective, emotional clarity, and the | The study authors acknowledged the sample size was small; thus, any null findings may be due to a lack of power to detect effects rather than a true lack of effect.<br>The study authors also acknowledge that all participants were exposed to the boot camp environment, the outcomes observed may have been augmented by the additional boot camp intervention. Therefore, the results may not generalise to other boys or young men, or to at-risk youth in other types of settings. |
|  | began; posttreatment measures were completed 1 to 3 weeks after the final group session. Participants completed treatment feedback measures at the final skills training group session. The tools used for pre, and post treatment measures were Personality Assessment Inventory-Adolescent (PAI-A), Difficulties in Emotion Regulation Scale (DERS), Distress Tolerance Scale (DTS), WAI-Short Revised (WAI-SR), and Treatment Evaluation Questionnaire.<br>**Outcomes:** Distress Tolerance Scale (DTS), Difficulties in Emotion Regulation Scale (DERS).<br>**Quality rating:** Moderate | **Setting:** Each DBT-A group session was conducted in a classroom at the NGYCP sites.<br>**Intervention details:** Each skills training group was led by two clinical psychology Ph.D. students. The clinicians were blind to pre- and posttreatment data. Each DBT-A group session was conducted in a classroom at the NGYCP outside of normal operating hours and followed a similar structure. The Two-hour skills training sessions covered: mindfulness (two sessions), distress tolerance (three sessions), emotion regulation (four sessions), and interpersonal effectiveness (three sessions).<br>**Comparator intervention or control:** A control group that completed pre- and post-test measures but received no additional intervention beyond the boot camp program (NGYCP alone). The NGYCP focused on eight core components: academic excellence, health and hygiene, job skills, leadership/followership, life-coping skills, physical fitness, responsible citizenship, and service to community. Master's-level counsellors provide individual and group counselling on an ongoing basis and are available for crisis intervention as needed. | control of impulsive behaviours when distressed. There were no significant differences between conditions in distress tolerance.<br>**Additional findings:**<br>**Acceptability**<br>Scores on the modified Working Alliance Inventory-Short Revised (WAI-SR) indicated a strong working alliance between DBT-A skills group participants and group leaders. Average scores on the WAI Total and all WAI subscales were in the “very often” to “always” range (i.e., between 4 and 5 on a 5-point scale). Youth also responded to the open-ended treatment evaluation questionnaire administered at the final session of the intervention (n = 28, 3 turned in blank feedback forms). Youth responded to this measure with overwhelmingly positive feedback. One hundred percent of participants responded “yes” to the question “Did this group help?” When asked “What part of the group did not work for you?” 18 of 28 (64.3%) participants reported no complaints and had no suggestions for improvement. When asked “what worked best,” most cadets who provided specific comments indicated mindfulness (9/28, 32.1%) or having a forum to discuss emotions (8/28, 28.6%). Additionally, participants made comments on the evaluation questionnaire such as “I can handle my anger much better now,” “I learned to stop and listen and to think before I speak. I really learned a lot of stuff with these people, and I like it. I just wish it could be longer,” and “y’all are awesome!”<br>The clinicians provided informal feedback about their experience implementing the program, including that cadets responded particularly positively to the behavioral components of skills training and reported that such components were easier for them to implement into their lives at the boot camp. They specifically reported success with Distract and TIPP DT skills (e.g., intense exercise, temperature), as these were easier to utilize in the boot camp setting (“self-soothe skills didn’t work for me because we can’t do a lot in here to soothe ourselves”). Cadets also reportedly appeared especially engaged when the group used role-play to practice IE skills, and many used the opportunity to practice communicating with authority figures, such as the cadres or their parents. Cadets regularly reported to clinicians that difficult emotions and/or mood dysregulation were met with an invalidating “suck it up and pull it together” attitude from their supervising cadres and peers in the larger program, and they noted that it was refreshing to have a forum for discussing emotions and learning skills to cope with difficult emotions instead of trying to ignore them. |  |
| Bertotti et al., (2025). <u>Social Prescribing for Young People in NEET Situations: A Realist Evaluation of the COPE Project in Italy and Portugal</u>. SSM -Mental Health, 100440.<br>(Italy and Portugal) | **Study design:** Mixed method uncontrolled before and after study.<br>**Aim:** To evaluate the mental health outcomes of a social prescribing intervention—C.O.P.E. (Capabilities, Opportunities, Places, and Engagement) project—on young people in a NEET situation in Italy and Portugal.<br>**Dates of data collection:** Interviews were conducted between June and September 2023. Quantitative data collection dates not specified.<br>**Data collection methods:**<br>**Quantitative:** A survey including: mental wellbeing (Short Warwick Edinburgh Mental Wellbeing Scale); Clinical Outcomes in Routine Evaluation - Outcome Measurement (CORE-OM): a 34-item scale that measures different aspects of psychological distress and wellbeing. | **Sample Size:** quantitative cohort data from 416 young people with qualitative interviews with 30 young people, 13 link workers, and 9 team members.<br>**Participants:** Young people who are NEET aged 16-34 (majority of participants were aged between 16 and 25; mean age of 23.3), females (50.4 %) and males (47.2 %). Comparatively, a higher proportion of Italian respondents had been diagnosed with a mental health issue (36 % versus 21.5 %) and were more likely to be taking medication (72% versus 58.1%).<br>**Setting:** Not stated (Italy and Portugal)<br>**Intervention details:** The intervention uses a social prescribing approach to address young people's social and mental health needs, focusing on their strengths. Through a link worker, who meets with them over flexible sessions, young people receive active listening and support to co-create an Individual Action Plan (IAP) that identifies, and guides tailored actions. For link workers to provide the best possible support to young people, a 'relational proximity community network' approach was adopted, which focused on working closely with several organisations, from the public, private and third sectors.<br>**Comparator intervention or control:** | **Primary findings:**<br>**Mental wellbeing:**<br>Wilcoxon signed rank test p-value analysis was performed on the sample. This finding revealed a statistically significant positive change between baseline and follow-up (95 % CI; $p < 0.05$; $s = 5.8$). A statistically positive change was also found in the Portuguese sample, whilst the Italian sample did not show a statistically significant change, although an improvement in mental wellbeing was also found. Mental wellbeing at baseline was lower in Italy (21.2) than in Portugal (23.7). At the same time, the change in mental wellbeing was more marked in Portugal than in Italy. a much greater positive change was experienced by females (1.76) than males (0.59) and for the younger population aged 16–24. Based on studies comparing the SWEMWBS with the PHQ-9, a widely used depression scale, it is possible to estimate the prevalence of depression in the young population. About 29 (23 %), nearly 1 out of 4, had probable clinical depression at baseline. This percentage was almost halved by follow-up ($n = 15$, 12 %). This group showed an average change of 4.4 points, which is considered 'meaningful' for this mental wellbeing scale (a score of three or above is considered 'meaningful' for the SWEMWBS). It is important to note that this is a small sample ($n = 29$) and there is a potential issue of regression to the mean, but it indicates a meaningful change of 4.4 points. With due caution, this may mean that C.O.P.E. had a relatively stronger impact on young people with more complex mental health issues at baseline.<br>**Psychological distress:**<br>Data was obtained from 89 people at both baseline and follow-up. All dimensions of the CORE-OM improved, particularly problems/symptoms which are related to questions about anxiety, depression, physical health and trauma. The dimension 'problems/symptoms' (anxiety, depression, physical health and trauma) has moved from a clinical to a non-clinical state. A Wilcoxon signed rank test was performed to establish whether there was a significant change from baseline to follow-up, where a declining score indicates an improvement in psychological wellbeing (decline in psychological distress). The intervention had a statistically significant positive effect on psychological distress, meaning that psychological distress decreased at follow-up ($Z = -4.064$, $p < 0.001$). The R-value which measures correlation between the two baseline and follow-up samples was $-0.43078$, indicating a moderately strong negative correlation between the baseline and follow-up samples.<br>**Additional findings:**<br>**Qualitative findings:** Young people reported a range of outcomes, including increased self-esteem, enabling them to recognise their strengths (asset- | This data also includes a small proportion of people in employment (2.3 %) and in education (6.7 %) due to the C.O.P.E. team agreeing to include those young people in particularly vulnerable situations who were attending short-term online training courses (e.g. foreign language) or in a casual work, that would not be classifiable under the definition of NEET. |
|  | **Qualitative:** in-depth 1-2-1 interviews<br>**Outcomes:** Mental wellbeing, psychological distress, interview findings.<br>**Quality rating:** Low | Pre-post comparisons were made. | based approaches) and learn to work more independently, particularly when seeking new jobs and training opportunities. Some young people reported that C.O.P.E. provided them with 'protection' and the first proper adult support they had received. For some young people changes were 'transformational' for others less so, but still useful and emphasising different aspects of the support received. The interviews revealed an increase in self-esteem and the capabilities to see their strengths ("[link worker] always pointed out my strengths [ ...] always saw a strength in me, even when I didn't see it because I was a bit down", IT_YP1).<br>Young people mentioned that the support they received helped them feel calmer and more confident about their actions, such as sending emails to potential employers. For many young people, link workers are the first type of adult and friendly support they have encountered ("I have no friends because I have a lot of difficulty finding friends and talking to a wiser, more mature person helps me", IT_YP10). Some of the young people in Portugal particularly emphasised the improvement of socio-professional skills ("I have acquired tools that have helped me in job interviews", PT_YP6) and increased social capital/ improved networking ("I met new people", PT_YP2). Young people mentioned they had become more active and accountable in their decisions ("I just got better and yes there was a bit of a clash on this last job too, to tell the truth, because my CSM network had offered me an internship of just a few hours and a few days a week and therefore a reduced salary and I still made that choice, I listened to all the link worker's advice up to that point, but on this job I did everything myself, I decided it", IT_YP1).<br>Young people appreciated the emotional support they received alongside the job search guidance, keeping them motivated. They found that the link workers support provided them with a space to reflect on their feelings and concerns ("we talked about my life, about my head, how it was, if everything was okay, psychologically, everything, if I was holding up. To summarise her work more, I think she was almost my 'real' psychologist", PT_3). All young people interviewed appreciated the 'informal' attitude of the link worker, which made them feel 'heard' without judgement ("she was very attentive, she listened a lot, she was very open", IT_YP5). In general, there was a balance between emotional and practical support. Link workers mentioned the importance of creating trust and offering young people something practical to do, which would increase their motivation and sense of responsibility ("I have this very nice image of this girl who wanted an agenda, i.e. to have appointments. So we started writing down appointments and tasks. For her this helped a lot, it almost gave her a reason to get up in the morning", IT_LW3). The focus was on supporting broader aspects of the young person's life which would help to create a routine ("support in organising the routine, whatever is necessary", PT_LW5). A multifaceted approach was used to involve young people, including volunteering or<br>activities aimed at developing personal and professional skills ("in creative workshops [...] workshops that teach how to do a job ", PT_LW3). In both countries, the relationship between the link worker and the young person's parents/carers was also very important, especially for minors. Some link workers have suggested an integrated strategy with both parents and the young person. More generally, reflecting on the intervention as a whole, a C.O.P.E. team member from Italy reported that social prescribing provided a rapid response 'activating' the young people ("we have a thin package. The young person also has to get active, it's not as if he/she can procrastinate so much, and, on the one hand, that's an advantage because in the world of youth, the long-term depth is not there. It is much more difficult in this era here and therefore this method is more accessible than others", IT_CT2).<br>Despite the many positive aspects reported above, the C.O.P.E. teams and link workers in both countries raised some challenges and suggested solutions for improving the intervention. Most respondents highlighted the considerable initial underestimation of the time and effort required to support young people in a NEET situation ("I think we underestimated just in general how much C.O.P.E. would take, not just the ratio per Link Worker, but also [...] we estimated that this would be done in like 2–3 afternoons a week or something like that", PT_CT3). Link workers also found it difficult to balance their role as link workers for C.O.P.E. with their traditional role in their organisation. It was recognised that supporting young people in a NEET situation requires significant flexibility and patience to establish trust and build a relationship. This often results in link workers spending considerable additional time with young people, well beyond their contracted hours. This is seen by link workers as unsustainable in the long term ("I was talking to a Link Worker who works at the employment agency, who is very motivated, very convinced, of the method. Unfortunately, the rest of her work requires a very fixed schedule of appointments, so she has a very tight timescale which is not necessarily compatible with the flexibility and speed needed to support the young person", IT_CT1). Some link workers also mentioned they might feel less experienced and skilled with some young people and their challenges, particularly those who have experienced extreme social isolation and introversion or those with high levels of mental vulnerability.<br>Qualitative insights emphasise the importance of trust between link workers and young people, highlighting flexibility, emotional support, and a holistic approach as key factors in fostering engagement. This led to the creation of three programme theories. One of these focuses on young people and is discussed in this article. The study concludes that despite some challenges related to sustainability, training and long-term impact, social prescribing can<br>be an effective tool for supporting youth in a NEET situation, particularly those suffering clinical mental health issues. |  |
| Conway & Clatworthy. (2015). <u>Innovations in Practice: Grow2Grow—engaging hard-to-reach adolescents through combined mental health and vocational support outside the clinic setting. Child and Adolescent Mental Health, 20(2), 112-115</u><br>(Kent, UK) | **Study design:** Uncontrolled before and after study.<br>**Aim:** To provide a preliminary evaluation of the Grow2Grow model to engaging and working with young people with complex mental health needs.<br>**Dates of data collection:** From June 2010 to July 2014.<br>**Data collection methods:** Following case discussion in Grow2Grow team meetings, the clinical team rated each young person's functioning from 0 (very poor functioning) to 100 (very high functioning) on entry to the project and at 3-month intervals. The Children's Global Assessment Scale (CGAS) was used to assess the young people's social, emotional and behavioural functioning on entry to Grow2Grow and at placement | **Sample size:** 21 young people completed Grow2Grow placements.<br>**Participants:** 13(62%) male, 8 (38%) female; Median age 17 years (range 14–25 years); 20 (95%) White British, 1 (5%) mixed White British/Greek Cypriot; 17 (81%) had co-morbidity (e.g. ADHD, ASD, learning disability, substance abuse, physical disability, chronic health difficulty); 15 (71%) NEET (Not in Education, Employment or Training), 4 (19%) At risk of becoming NEET, 2 (10%) Engaged in training/education; median placement length was 14 months (range 2–24 months)<br>**Setting:** Common work, an established organic farm, conference and study centre in Kent. (Market garden, organic dairy farm and conference centre).<br>**Intervention details:** Grow2Grow provides therapeutically supported vocational placements for young people with complex mental health needs. Young people attend the project for up to 2 days a week for a maximum of 2 years. Participants gain vocational skills through working in the social enterprises at Common work, including a market garden (growing vegetables to sell in the local community and to the on-site conference centre), organic | **Primary findings:**<br>**Attendance:** From January 2013 until the end of July 2014, the attendance rate was 82% (443/540 placement days attended).<br>**Functioning:** Young people's change in functioning mean CGAS score on entry to Grow2-Grow was 35.9 (SD=7.4) and on leaving was 47.5 (SD=10.9), which represents a significant improvement in functioning (t (20) -5.6; p< .001). However, it is important to note that there was variation in CGAS scores over time, with most individuals experiencing both increases and decreases over the course of their placements. There was also considerable variation across individuals. While the mean change in CGAS scores was 11.6 (SD = 9.4) (typically a shift of one band on the CGAS tool – e.g. from 'Serious Problems' to 'Obvious Problems'), three young people ended their Grow2- Grow placements with no increase in CGAS score and seven young people moved two or three CGAS bands over the course of their placements.<br>**Additional findings:**<br>**Feedback from young people:** The mean score on the satisfaction questionnaire was 31.7 (SD = 2.3) of a maximum of 35, indicating a very high level of satisfaction with the Grow2Grow project. Young people reported learning a range of skills including horticultural/agricultural skills, life skills (e.g. cooking, travelling independently) and social skills. For example, one young man wrote 'I have learnt lots of new skills including gardening, cooking, talking more, making friends and getting quicker at doing jobs'. In terms of their personal development, they described mainly social and emotional changes. For example, 'I am more confident, less anxious and able to go up to people and start a conversation. I have found what I would like to do [be a gardener] and now know who the real me is'. | The study included a heterogeneous sample comprising individuals who are NEET, those at risk of becoming NEET, and individuals currently engaged in education or training. While the majority of the sample were NEET, the study did not provide a separate sub-analysis of findings for this group. |
|  | completion. All young people completed a project evaluation sheet on leaving the project, reflecting on what they had learnt and how they had changed. They also completed a seven-item satisfaction measure.<br>**Outcomes:**<br>Attendance; Change in functioning; feedback from participants.<br>**Quality rating:** Low | dairy farm and conference centre. They also develop life skills, through cooking and eating lunch with the group each day and working towards independent travel to the project. All young people attending Grow2Grow receive weekly 1:1 therapeutic key working. All members of the multidisciplinary Grow2Grow team (psychologists, psychotherapist, occupational therapist, horticultural therapist and psychology/psychotherapy trainees) are trained and supervised to use this model.<br>**Comparator intervention or control:** Pre-post comparisons were made. |  |  |
| Davies et al., (2020). <u>Using engagement in sustainable construction to improve mental health and social connection in disadvantaged and hard to reach groups: a new green care approach</u>. *Journal of Mental Health*, 29(3), 350-357. (UK) | **Study design:**<br>Uncontrolled before and after study.<br>**Aim:** To provide the first pragmatic examination of the impact of engaging in a brief outdoor sustainable construction project (Down to earth) on the mental health and social connectedness of "hard to reach" and disadvantaged groups.<br>**Dates of data collection:** September 2016 to March 2017<br>**Data collection methods:** Data were | **Sample Size:** Study 1: n=93; Study 2: n=55.<br>**Participants:**<br>**Study 1:** All participants were considered "disadvantaged" and "hard to reach" namely, young adults not in education, employment or training. 93 young people (68 men: 20 women: 5 did not disclose their gender; average age 19 years). One cohort completed only the PHQ-4 (n=28; 21 men: 7 women).<br>**Study 2:** Participants were from a number of groups including asylum seekers, the long-term unemployed and men with longstanding depression. 55 adults (40 men: 15 women; average age 36). Although this group included a small number of younger people (18–25) their | **Primary findings:**<br>**Baseline findings:**<br>At baseline, the young adult participants in study 1 were found to have significantly less connection to the community ($t=4.398$; $p<.001$) and to Down to Earth ($t=2.735$; $p<.01$) and reported lower wellbeing ($t=2.447$; $p<.05$) than those in study 2. Using the National Household Survey of England average (of 23.6) as a benchmark, participants of study 1 reported average wellbeing levels well below this ($n=60$; 21.3) whilst study 2 participants were just below this level ($n=49$; 23.4). No differences between the groups were observed for baseline anxiety, depression or resilience. On the basis of these findings, authors decided to combine data from both studies for anxiety, depression and resilience whilst analysing the data for connection and wellbeing for each group.<br>**Mental health: depression, anxiety and resilience:**<br>When data from all participants regardless of baseline score were analysed ($n=123$), no changes over time were recorded on any of the measures. When inclusion in the analysis was limited to only those falling at or above the cut-off threshold at baseline, statistically significant improvements were found for depression ($n=27$; $t=-4.21$; $p<.001$); anxiety ($n=43$; $t=-6.39$; $p<.001$) and resilience ($n=24$; $t=3.73$; $p<.001$). Of those meeting the threshold criteria for depression at time 1 ($n=27$), 15 (55%) no longer met the threshold at the end of their participation and could be considered to meet the clinical | In Study 2, the average age of participants is 36, which exceeds our age group mentioned in the inclusion criteria. It is not reported how many participants were within our age range. Please note that authors combined data from both studies for anxiety, depression and resilience findings. |
|  | collected in the training room on-site at the start of day 2 (baseline) and 7 (post-intervention) of 8-week programme and took approximately 5–10 min to complete. Participants completed four brief self-report measures in addition to providing their age and gender. The tools used to measure the outcomes were the Patient Health Questionnaire (PHQ-4), the Short Warwick-Edinburgh Mental Wellbeing Scale (SWEMWBS), the Brief Resilience Scale (BRS) and the adaptation of the Inclusion of Community in the Self Scale.<br>**Outcomes:** Mental health: Depression; Anxiety; Resilience; Wellbeing and Social connection.<br>**Quality rating:** Low | route to accessing the scheme was because they belonged to another “excluded group” (e.g. asylum seekers).<br>**Setting:** Training room on-site, each site has been developed over time to include a range of buildings (including classrooms, offices and kitchens) that are constructed from natural products such as timber, cob, straw and lime render, with a range of sustainable living features (e.g. solar technology, allotment gardens, animals and waste water treatment) on-site.<br>**Intervention details:** Individuals attended a group based sustainable building project at one of two sites for one full day per week over an 8-week period (maximum 8 sessions; typically 6–12 members per group). During each session participants are supported by staff to engage in group and individual tasks based on construction skills (e.g. timber frame construction; lime rendering) and outdoor skills (e.g. cooking with an open fire). Communal times included lunch.<br>**Comparator intervention or control:** Pre-post comparisons were made. | significance criteria for “recovery”. For anxiety, 21 (49%) of the 43 participants meeting the anxiety threshold would be considered to meet the clinical significance criteria for “recovery” at time 2. Using a more demanding method (where inclusion was determined by meeting the anxiety or depression cut-off or both at time 1, and “recovery” requires both anxiety AND depression to be below cut-off at time 2), 20 (42%) of the 48 individuals included in the analysis met this clinical significance criteria for “recovery”. Reliable improvement for anxiety and depression required a change in score of two points or more between time 1 and time 2. Of the 43 individuals reaching the cut-off for anxiety, 23 (53%) showed reliable improvement and only 2 (5%) showed reliable deterioration. For depression, 13 (48%) of the 27 individuals meeting the cut-off at time 1 showed reliable improvement with 1 (4%) showing deterioration. Combining these two approaches allowed the calculation of reliable “recovery”. Of the 48 individuals with elevated anxiety or depression scores (or both) at time 1, 16 (33%) met this stringent criteria for “reliable recovery”<br>**Wellbeing**<br>**Study 1: NEET population**<br>Of the 33 individuals in this group with wellbeing data at both time points, 12 individuals had scores below the threshold at time 1. Although the average wellbeing score improved over time, the level of change was not statistically significant and mean scores remained below the general population (n=12; M=17.11(SD:1.56) vs 18.21 (SD:2.17); t=-1.31; p=not significant).<br>**Study 2: Adults (asylum seekers, long-term unemployed)**<br>Complete wellbeing data were available for 39 individuals in this group; 7 of whom showed low levels of wellbeing at baseline. As with study 1 participants, the improvement over time was not statistically significant and mean scores remained below the general population (n=7; M=17.31 (SD:1.03) vs 19.08 (SD:2.77); t=-1.366; p=not significant).<br>**Social connection**<br>**Study 1: NEET population**<br>The significant changes with a large effect size can be seen over time for those who reported difficulties in this area (n=23 in relation to the community at large; M=1.78 (SD:0.85) vs 2.78(SD:1.24); p<0.005) (n=12 Down to Earth; M=2.33(SD:0.78) vs 4.0(SD:0.74); p<0.001). For the “community at large” ratings, the time 2 score approached the threshold score (3) whilst this threshold was exceeded with respect to Down to Earth.<br>**Study 2: Adults (asylum seekers, long-term unemployed)**<br>For study 2 participants reporting difficulties (n=19 in relation to the community at large; M=2.21(SD:0.71) vs 3.89(SD:1.7); p<0.001) |  |
|  |  |  | (n=9 Down to Earth; M=2.11(SD:0.93) vs 5.0(SD:0.87); $p<0.001$), showed statistically significant improvements in social connection, both in general and within the project. In addition, the mean score at time 2 (for those with poor initial wellbeing) was considerably higher than the threshold score of 3.<br>**Additional findings:**<br>In conclusion, participants with poor mental health and social connection at baseline showed statistically and clinically significant improvements in depression, anxiety, resilience and social connection by the end of the brief intervention. |  |
| De Lannoy, Graham & Grotte. (2024). <u>Can a holistic coaching and referral programme enhance well-being and employability amongst NEET youth?</u><br>(South Africa) | **Study design:** Mixed method uncontrolled before and after study.<br>**Aim:** To evaluate the achievement of the key anticipated outcomes of employability and wellbeing, for the young participants in the Basic Package of Support (BPS) programme.<br>**Dates of data collection:** March 2022 to November 2023.<br>**Data collection methods:**<br>The quantitative component involved an intake survey assessing demographics, household situations, employability, and wellbeing, followed by a second survey after participants completed at least three coaching sessions. General mental health and wellbeing | **Sample Size:** 516<br>**Participants:** NEET people aged between 18-27 (94.57% were aged between 18-24), (Females: n=331; males: n=185). More than half of the sample (51%) had completed Grade 12, with over 17% having a Bachelor's pass. Around 30% had between Grade 9 and 11, and under 2% had less than Grade 9. Less than a fifth of the sample (17 %) had a university degree. The vast majority of young people had been searching for employment or training/volunteering opportunities (77% and 73% respectively), with a similar gender distribution.<br>**Setting:** Three BPS sites: Orange Farm in Gauteng, Atlantis in the Western Cape, and Cato Manor in KwaZulu Nata.<br>**Intervention details:** The Basic Package of Support (BPS) as an intervention to support young people who are NEET, and to (re)connect them to learning and earning opportunities. The Intervention includes individualised face-to-face holistic coaching to address barriers NEET youth face | **Primary findings:**<br>**Outcome 1 better mental health outcomes, feel more supported, and are more resilient, pre-session 1 to post-session 3.**<br>**Sense of wellbeing:** BPS beneficiaries demonstrated a distinct improvement for the cohort that moved from pre-session 1 to post-session 3, with a highly statistically significant difference ($p = 0.0010$). By post-session 3, only 36% indicated that they were suffering or struggling. There was a statistically significant improvement for the Life Evaluation Wellbeing Index, with an increase of over 10 percentage points in the number of youths who were thriving post session 3 compared to pre-session 1 (t-test: $p=0.0100$).<br>**Mental Health:** there was a significant improvement in GAD-2 scores, indicating a decrease in anxiety in respondents who completed their third coaching session ($p=0.0129$). Almost 40% of the participants presented with possible generalised anxiety disorder at pre-session 1, this proportion decreased by 14.85 percentage points after completion of three coaching sessions, with the difference being statistically significant. When looking at K6 findings, the total percentage of participants with moderate to high psychological distress stayed the same by post-session 3, however more had moved from high psychological distress to moderate psychological distress, indicating a reduction in high distress of almost 13 percentage points. The difference in mean scores was significant at the 10% level (t-test: $p=0.0804$).<br>**Self-esteem:** There was a small, yet consistent, improvement in youths' feelings about themselves and decrease in the proportion of those who felt they could not do anything right (3.55 percentage points) and those who had more things that they did not like about themselves (2.3 percentage points), was supported by an increase in those who generally felt good about themselves (almost 3 percentage points). T-test: $p = 0.2622$.<br>**Sense of support:** There was no significant difference in the mean scores for youths' feelings of being supported there were fewer respondents reporting feelings of being isolated and alone, or disconnected from other people. However, this finding was accompanied by small decreases in |  |
|  | improvements were assessed by the Cantril's Ladders and Life Wellbeing Evaluation Index, as well as the Generalised Anxiety Disorder (GAD-2) and Kessler Psychological Distress Scale (K6) outcomes.<br>The qualitative component included in-depth interviews with youth who had completed multiple coaching sessions. Interviews were conducted six to eight months after pilot sites were opened to explore participants' situations, experiences of coaching, and any shifts in perspective. Participants were randomly selected from those who had completed at least three sessions, and invited to participate in the interviews.<br>**Outcomes:**<br>Outcome 1: Youth have better mental health outcomes, feel more supported, and are more resilient, pre-session 1 to post-session 3. | to accessing opportunities; creation of action plans and referral to service providers and ongoing support provision. Additionally, working with local service partners to improve service delivery to young people using a Community of Practice (COP) approach.<br>**Comparator intervention or control:** Pre-post comparisons were made. | feeling connected most of the time or consistently. The greatest change was the increase in 'kind of feeling connected' (almost 9%).<br>A general improvement in access to peer support was found, with the difference in means being statistically significant at the 5% level (t-test: $p=0.0337$). The greatest positive change was a nearly 8 percentage point increase in youth indicating that they had friends that they could trust and rely on. This finding was perhaps not surprising as young people would meet the peer-to-peer mobilisers at the sites, would also engage with other young people in peer connect sessions and had started spending time at the sites.<br>**Outcome 2: NEET youth have improved navigational capacities**<br>**Ability to handle stress:** The findings are mixed in terms of how young people handle their stress and emotions. The difference in mean scores was not significant (t-test: $p=0.6547$). The greatest change was the nearly 15 percentage point increase in those who were learning to deal with stress and emotions. Decreases in those who felt overwhelmed and did not know what to do about it, was offset by a decrease in those who indicated that they knew how to deal with their stress and emotions in healthy ways at baseline (10.65 percentage points).<br>**Asking for help when upset and feeling overwhelmed:**<br>While the difference in means was not statistically significant (t-test: $p=0.1517$), an encouraging and large decrease in youth who felt that they could not be helped or did not seek help were seen (8.28 percentage points). Similar to the shift above there was a move towards the middle: young people who were "learning to" or "knew where to get help" when they were upset or overwhelmed.<br>**Knowledge of, and access to services:** There was a consistent, and highly statistically significant, improvement in young people accessing services and resources in their community (t-test: $p=0.0000$). There was an 18.35 percentage point drop in the proportion of youth who indicated not accessing any services in their community as they knew they would not be helped. This drop was coupled with a nearly 6 percentage point increase in youth who accessed a variety of helpful resources and services.<br>**Outcome 3: Youth connect to learning and earning opportunities**<br>There was a decrease in the proportion of NEET youth across the various data collection periods. Following their first session, 88.39% of youth were NEET. This figure decreased steadily to the lowest proportion of 64.37% following Session 4. The difference between the NEET rate post-S1 and post-S4 was highly statistically significant ($p=0.0000$).<br>**Additional findings (Qualitative findings):**<br>**Sense of wellbeing** |  |
|  | Outcome 2: NEET youth have improved navigational capacities.<br>Outcome 3: Youth connect to learning and earning opportunities.<br>**Quality rating:** Low |  | Participants indicated that, prior to starting the programme, they had been feeling unhappy about life, lost about how to move forward in their lives, and as if they “were stuck”. Part of their frustration was not having anyone to talk to about how they were feeling. In contrast, the participants reflected on how the coaching had helped them to feel calmer, more in control and optimistic: *“And then, like when I leave, I feel like a little calmer, you know, like I’d come to her before going to work just to talk to her. And then I’d leave for work leaving like feeling, you know, optimistic. And I’d say, hey, I can take this. Yeah. And stuff like that. Coaching helped me cope and feel more optimistic.”* (Male employed youth, Cato Manor Round 1).<br>**Mental health:**<br>The testimonies supported the results showing positive shifts in mental health among young people as they engaged with the BPS programme and provide us with a sense of how BPS was able to establish these shifts: *“All these years I was lost, I was living but not knowing the purpose of living, up until I find this programme called BPS.”* (NEET Male, focus group participant, Orange Farm).<br>The narratives of young people indicate the positive effects of having a trusted person to turn to and to confide in, who provides support, clear guidance and a sense of direction in life. There were also clear references from young people to the supportive nature of the entire site teams, and the safety of a BPS site that provided them with a sense of belonging and that helped them feel less hopeless.<br>**Sense of support:**<br>The narratives illustrate how BPS manages to shift that sense of disconnect and to build young people’s understanding of how to engage and connect differently, more positively, to people around them. The data show how the trusted role of an individual coach was instrumental in these shifts: *“Just being able to speak to my coach about anything. She was not exposing me, and she was not talking about me. I felt like I needed to talk to someone because at that time, I felt like I had a lot of anger [...] I didn’t know how to express my feelings towards anyone, and I hated people.”* (NEET Female, focus group participant, Orange Farm)<br>**Ability to handle stress:**<br>Some young people testified to the positive effect on their stress levels of having a caring place to go to and a supportive BPS team: *“I felt comfortable [a lot] because they welcomed me really well and at the present moment, I don’t have any stress. I feel like I’m at home”.* (NEET Female, Round 1 individual interview participant, Orange Farm).<br>Youth expressed an enhanced belief in their ability to cope, their resilience and their engagement with others, as they connected to a BPS coaching journey: |  |
|  |  |  | *"I was always running away from challenges so when I faced them, that's when I became aware that in life you have to face challenges in order to be stronger. [...] It has shifted a lot of fear anxiety. At first, I was [...] anxious about everything. Going to varsity- I was anxious" (NEET Female, Round 1 individual interview participant, Orange Farm).*<br>**Knowledge of, and access to services**<br>Young people detailed the practical ways in which the BPS had helped them understand what services and opportunities were available to them, but also what was needed to access these services and resources.<br>*"You come and it's almost like they take your hand and that's how my experience was to get from this point to the next. That's when I was like okay if I for instance if I want to apply now for my license, I know this is where I need to go and get the training I need". (NEET Male, individual interview participant, Round 1, Atlantis).*<br>**Youth connect to learning and earning opportunities**<br>The qualitative narratives too reflected encouraging NEET to EET transitions, also demonstrating the coaches' ability to solve both for the often immediate need to earn an income and for the longer term need to remain focused on a training trajectory that would bring young people closer to their aspired futures.<br>*"I did my N1 and N2. [...] [But my brother] the breadwinner in my house moved [...] And there was no more breadwinner. My mother was determined for me to go on and finish my N but then [...] I didn't want to go on because my family was struggling and I have the ability to provide for them. So, I didn't know what to do. [...] [my coach], asked me 'can you get a weekend job?' and I'm like 'Why didn't I think of that? [...] if I can get the weekend job, I can pay for my traveling [...] because when my brother left, we didn't have money to pay for my traveling, and it was quite expensive [...] But now I am convinced [that I must also continue studying] because [...] when we drew up my action plan], I looked and yoh: if I stop, I cannot go further, so I need to go. So, the next thing was 'OK, we're gonna get that weekend job'. [The team helped me apply] and just a few weeks into college, I got a phone call [and then I got the job]." (NEET Male, individual interview, Atlantis).* |  |
| Kendall & Maujean. (2015). <u>Horse play: A brief psychological intervention for disengaged youths.</u> | **Study design:** Non-randomised controlled study.<br>**Aim:** To examine a brief psychological intervention | **Sample Size:** Intervention group (n=16, 12 of which completed the program) control group (n=10)<br>**Participants:** Disengaged adolescents, including those who have disengaged from school and | **Primary findings:**<br>**Self-esteem, Intervention group:** For the RSES scores, the ANOVA revealed a significant effect for time (Wilks's Lambda=0.09; $F(2,8)=42.72$; $p=0.0001$; multivariate partial eta squared=0.91). A post-hoc test using the Bonferroni correction revealed a significant increase in participants' level of self-esteem across the three timepoints (i.e., between preintervention and midintervention, $p=0.007$; between midintervention and postintervention, $p=0.002$; and between preintervention and postintervention, $p=0.0001$). | The sample does include those younger than our age specified in the inclusion criteria, but the ages range from 12-22 years. |
| *Journal of Creativity in Mental Health*, 10(1), 46-61.<br>(Queensland, Australia) | delivered to a group of disengaged youths using horse-related activities to improve their levels of self-esteem and self-efficacy.<br>**Dates of data collection:** Not stated<br>**Data collection methods:** The Rosenberg Self-Esteem Scale (RSES) and the General Self-Efficacy Scale (GSE) were administered. The Social Behaviour Observation Form (SBOF) was developed for this study to rate participants' positive and negative behaviours during every session. Case manager surveys were conducted to gain their perceptions of participants' strengths and the challenges they faced in various aspects of their daily lives.<br>**Outcomes:** Self-esteem, Self-efficacy, social behaviour observation, case manager survey findings<br>**Quality rating:** Low | are considered to be at risk for negative outcomes.<br>**Intervention group:** 11 boys and 5 girls, with a mean age of 15.44 years (SD = 3.12 years, range = 12–22 years).<br>**Control group:** 7 boys and 3 girls, with a mean age of 15.20 years (SD = 0.79 years, range = 14–16 years).<br>**Setting:** Not stated<br>**Intervention details:** The intervention group received 10 weekly sessions each beginning with an introductory session where the basic theory and principles of natural horsemanship were discussed. Participants watched a promotional DVD, which contained an inspirational message and a brief overview of the Parelli Seven Games. This initial session also included an introduction to the tools and the theory that underpinned these tools prior to any sessions with the horses. Each subsequent session began with a discussion of participants' expectations, their feelings about playing with horses, and how an understanding of horses might relate to their own lives. The "hands-on" sessions began with demonstrations of natural horsemanship followed by practice. Each participant was matched to a horse based on assessments of his or her internal or external orientation. The participant–horse pairs were also matched to a mentor who assisted them with the practice sessions. In the last few sessions, participants | **Self-efficacy, Intervention group:** For the GSE scores, the ANOVA also revealed a significant effect for time (Wilks's Lambda=0.15; $F(2,8)=21.90$; $p=0.001$; multivariate partial eta squared=0.85). A post-hoc test using the Bonferroni correction indicated that participants' level of self-efficacy significantly increased between preintervention and postintervention ($p=0.009$) and between midintervention and postintervention ($p=0.048$). However, there was no significant difference in GSE scores between preintervention and midintervention ($p=0.243$).<br>**Control group findings:** There were no significant differences in RSES scores (Wilks's Lambda=0.93; $F(1,6)=0.43$; $p=0.535$; multivariate partial eta squared=0.07), or GSE scores (Wilks's Lambda=0.93; $F(1,6)=0.47$; $p=0.518$, multivariate partial eta squared=0.07) between Time 1 and Time 2.<br>**Comparison of groups:** There was no significant difference in RSES scores at Time 1 for participants in the Horse Play and control group. In contrast, a significant difference was found in RSES scores at Time 2 between the two groups. The magnitude of the differences in the means (means difference= 7.16; 95%CI:3.87 to 10.44) was large (eta squared = .57; $p=0.0001$). No significant differences were found for GSE scores at Time 1 and Time 2 between the two groups. Although there was a nonsignificant result between the two groups at Time 2, the magnitude of the differences in the means (means difference=3.49; 95% CI:–1.39 to 8.38) was large (eta squared=0.27; $p=0.15$).<br>**Additional findings:**<br>**Observation sessions:** Levels of social behaviour remained relatively stable from Session 1 to Session 6 and increased in the remaining two sessions (Sessions 7 and Session 8). When comparing participants' total behaviour score in Session 1 to the total score in Session 8, there was a significant overall improvement in positive behaviours (Session 1, $M=18.33$; $SD=5.88$; Session 8, $M=30.00$; $SD=6.06$), ($t(3)=-13.22$; $p=0.001$).<br>**Case manager survey:** A paired-sample t-test revealed that there was a significant increase in scores from preintervention to postintervention on five of the six domains pertaining to case managers' perceptions of the young people's abilities. Working with others ($p=0.04$); Working on his/her own ($p=0.29$); Following rules ($p=0.04$); Getting work done ($p=0.04$); Energy level ($p=0.09$); and confidence to try new things ($p=0.08$). The eta-squared statistics indicated a | Disengaged youth not clearly defined, however participants were deliberately selected to represent those who had not responded to previous traditional interventions and both groups were considered to have similar background and were disengaged from the community. |
|  |  | prepared for a Parelli Level 1 exam. These auditions consisted of a 10-min video recording that demonstrated their acquisition of the key natural horsemanship skills while playing with their horses on the ground. At the end of each week, participants discussed the skills or abilities they had learned during the practice session. These group discussions were led by a psychologist and included the team of mentors and the instructor. The groups enabled members to provide positive feedback to the participants and encouraged discussion about how these new skills and knowledge could be applied in their everyday lives. A list of 31 qualities was constructed to determine all the qualities that might be facilitated by the intervention. Every week, the young people were asked to select one word from the list because they believed they had learned something about that quality during the session and intended to implement or practice that quality in their everyday lives prior to the next session. A discussion was then held about how they had learned that skill or quality and how they aimed to implement it in their own lives. Their experiences during the week were discussed at the beginning of the following session, when they were encouraged to describe ways in which their new skill had been useful.<br>**Comparator intervention or control:** Pre-post comparisons were made as well as comparisons<br>to the control group who received no intervention. | large effect size for all five domains (0.26–0.37) and a moderate effect size (0.11) for the only nonsignificant finding—namely, participants' ability to work on their own.<br>**Words selected throughout the intervention**<br>All participants engaged with the psychological component of the intervention by selecting words to represent their areas of progress each week. These words were examined across all of the participants and sessions and revealed a focus on working collaboratively (e.g., cooperation, respectful behaviors, team player, communication; 15 of 31 words selected, or 48%) and on interpersonal sensitivity (e.g., patience, tolerance, forgiveness, kindness; 9 of 31 words selected, or 29%). Words representing these two areas tended to appear earlier in the program, whereas words representing personal stability (e.g., confidence, determination, consistency, commitment, competence; 9 of 31 words selected, or 29%) tended to appear later in the program. Finally, words representing personal growth or complex concepts (e.g., leadership, wisdom, perceptiveness, balance; 7 of 31 words selected, or 23%) appeared in the final session. |  |
| Lee et al. (2013). <u>Home visitation program for detecting, evaluating and treating socially withdrawn youth in Korea</u>. *Psychiatry and Clinical Neurosciences*, 67(4), 193-202.<br>(South Korea) | **Study design:** Uncontrolled before and after study.<br>**Aim:** To approach socially withdrawn youth (SWY) therapeutically, through a home-visit program, and evaluate the SWY treatment outcomes after home-visit psychotherapy.<br>**Dates of data collection:** Not stated<br>**Data collection methods:** The self-rating scales for participants were the Derogatis Symptom Checklist-90-Revised (SCL 90-R), Kovac's Children's Depression Inventory (CDI), Beck's Depression Inventory (BDI), Rosenberg's Self-esteem Scale (SES), Spielberger's Trait Anxiety Inventory (STAI), LaGreca's and Stone's Social Anxiety Scale for Children-Revised (SASC-R), Watson's and Friend's Social Anxiety and Distress Scale (SADS), Seo's and Hwang's Avoidant Personality | **Sample Size:** 41 intervention group; 239 control group<br>**Participants:** Socially withdrawn, aged under 25 years, refusal to attend school, unemployed, and mainly staying at home all day for more than 3 months. 31 were male (75.6%) and 10 were female (24.4%). Their average age was $16.5 \pm 3.8$ years for male and $16.1 \pm 2.5$ years for female participants.<br>Of the controls, 173 were male (72.4%) and 66 were female (27.6%), they were middle and high school students.<br>**Setting:** Home-based<br>**Intervention details:** A home visitation program, where a team comprising a caseworker and an assistant, visited each SWY's home. Separate interviews were conducted for the SWY and the parents, and each interview session took about 1hr. Plans for five sessions of person-centred psychotherapy were created. The caseworker conducted a structured interview, in accord with the manual, at each SWY's first session. Self report scales were collected as part of the interview.<br>**Comparator intervention or control:** Pre post comparisons were made. | **Primary findings:** Participants' mean number of psychotherapeutic sessions was 2.8, and the mean number of parental interview sessions was 3.4.<br>**SWY reported data:** At baseline the SWY had significantly higher scores in depression, anxiety, and Internet addiction than controls did. Moreover, they had lower scores on the SES. Although it is not standardized for Korea yet, the SWY mean IAS was higher than that of controls ($P = 0.008$).<br>**Parent reported data:** On the FES, the SWY participants showed statistically significant differences from normal controls on the cohesion, active-recreational orientation, organization, and control sub-scales. Regarding the HYQ, filled out by the parents, the SWY group again showed significantly higher scores than did the control group ($59.9 [SD = 17.3]$ vs $21.8 [SD = 9.1]$; $P < 0.001$).<br>**Additional findings:**<br>**Caseworker reported data:** Many interviews were difficult due to an SWY's refusal and/or uncooperative attitude; this study lost four participants who would not participate in any interviews. Many cases moved to treatment centers after these home visits, such as attending community mental health centers (eight cases), visiting psychiatric clinics (eight cases), participating in hikikomori camp (one case), and online counseling (one case). Satisfaction with the home visitation program was higher in the parent group than in the SWY group. The multi-selected behavior changes after these home visitations were as follows: increased outdoor activities, 14 participants; more family conversation, 13; increased interpersonal contacts, 11; improved family relationships, 11; spending more time outside, nine; increased participation in group activities, six; returning to school, four; acquiring part-time jobs, two; and going to a private academy, one participant. Regarding the home visitation program, 75.6% of the parents, 48.7% of the caseworkers, and 43.9% of the participants had positive responses. The caseworkers reported the following degrees of improvement: a little bit improved, 51.2%; no changes, 31.7%; moderately improved, 14.6%; and greatly improved, 2.4%. Participants' average GAF scores increased significantly after the home visitations ($44.6 [SD = 11.1]$ vs $53.4 [SD = 13.2]$; $p < 0.001$). However, 48.8% of the SWY showed no change in GAF score. |  |
|  | Disorder Scale (APDS), Lee's and Shin's Socially Withdrawn Youth Questionnaire (SWYQ), and Young's Inter-net Addiction Scale (IAS).<br>The parents' reporting forms comprised Moos' and Moos' Family Environmental Scales (FES), the Parents' Brief Diagnostic Interview Schedules for Children (DISC-IV), and the Hidden Youth Questionnaire (HYQ).<br>The caseworkers' reporting forms contained the records for the families' and participants' histories and the outcome evaluations, using Global Assessment Functioning (GAF) and Clinical Global Impression (CGI) scores, from the first and last visits.<br>**Outcomes:** All the scales listed above for participants, parents and caseworkers.<br>**Quality rating:** Low |  |  |  |
| Roemer et al., (2021). <u>Effectiveness of a low-dose mindfulness-based intervention for alleviating distress in young unemployed adults</u>. *Stress and health*, 37(2), 320-328.<br>(New Zealand) | **Study design:** Non-randomised controlled study.<br>**AIM:** To investigate the effects of a low-dose mindfulness-based intervention (MBI) on distress in a sample of young unemployed adults.<br>**Outcomes:** Wellbeing, psychological distress and mindfulness<br>**Dates of data collection:** Not stated<br>**Data collection methods:** Self-report surveys were collected before and after the intervention: The short Warwick-Edinburgh Mental Wellbeing Scale (SWEMWBS) was used to measure wellbeing; The Kessler 10 Psychological Distress Scale (K10) was used to measure psychological distress; and a shortened version of the Five Facet Mindfulness Questionnaire was used to assess dispositional mindfulness (FFMQ-SF).<br>**Quality rating:** Low | **Sample size:** 239, 59 of which were lost to follow-up, (Intervention group: 86; Control group: 94).<br>**Participants:** Participants were members of the Youth Development Unit (YDU), a development programme offered by the New Zealand Defence Force (NZDF). Unemployed young people, aged 17 to 25, mean age (19.91 years). There were almost double as many males (64.4%) than females (35.6%). In terms of ethnicity, the sample consisted of Māori (32.6%), Joint Ethnicity (e.g. NZ European/Māori; 31.4%), NZ European (23.8%), Pasifica (8.8%) and Others (3.3%).<br>**Setting:** The programme was delivered at three different training camp locations across the country.<br>**Intervention details:** All participants were receiving a 6-week intervention including a mix of activities to build self-confidence, teamwork, and life skills. Participants engage in classroom learning, skills workshops, military-style team-building activities (e.g. marching and drill), physical exercise as well as outdoor exercises.<br>The intervention component being tested was the mindfulness training: a weekly contact session of one hour over a period of four weeks, facilitated by a qualified mindfulness instructor. In addition, participants were encouraged to practice<br>mindfulness in their daily lives, such as brushing teeth mindfully or eating mindfully, and to use a mindfulness app for 10 to 15 minutes before their lunch break. However, daily practice compliance was not monitored. In their first contact session participants were introduced to mindfulness and its immediate effects. The second contact session was themed around thinking and mindfulness and the third contact session focused on emotions and the interplay with thinking. In week four participants learned about the principle of radical acceptance. In order to support and foster learning, participants were given an info booklet after the first week. A typical contact session used a mix of audio and visual material to introduce participants to each topic. After that, participants engaged in mindfulness-based exercises, such as body scans, breathing, identifying internal and external sensations as well as judgments when they arise. Participants also had time for feedback and reflection.<br>**Comparator intervention or control:** The control group received the normal Youth Development Unit (YDU) course content, and the intervention group also received mindfulness training. Pre-post comparisons and comparisons to control group were made. | **Primary findings:** Wellbeing for the entire sample at baseline correlated negatively with distress at T1 ($r=-0.45$) and T2 ($r=-0.27$). The noticeable difference in magnitude of these correlations indicates that baseline level of wellbeing may influence the outcome of the intervention. Similar correlation patterns were observed between mindfulness at T1 and distress at both T1 and T2.<br>A mixed model ANCOVA showed that psychological distress was inversely and significantly predicted by both covariates: mindfulness ($F(1, 172)=28.75$; $p<0.001$), and wellbeing ($F(1, 172)=7.04$; $p=0.009$). After accounting for the effect of covariates there was a significant effect of time ($F(1, 172)=15.17$; $p<0.001$) and a significant interaction between group and time ($F(1, 172)=4.06$; $p=0.045$). This shows there is an overall distress reduction observed in both groups as well as a significant effect of the mindfulness intervention. After accounting for the effects of both covariates, estimated distress mean scores are higher in the intervention group at the baseline (Time 1) and lower after intervention (Time 2) compared to the control group. While distress is reduced in both groups at Time 2, a stronger reduction is observed in the intervention group.<br>Subsequent post-hoc tests showed that scores of wellbeing ($t(175)=-9.23$; $p<0.001$; $d=0.70$), distress ($t(177)=8.16$; $p<0.001$; $d=0.61$) and mindfulness ($t(179)=-5.36$; $p<0.001$; $d=0.40$) improved for the sample as a whole from baseline to post-intervention.<br>**Additional findings:** This study found that a low-dose MBI can enhance mindfulness and decrease psychological distress in a sample of young unemployed adults. However, findings indicate that a low-dose MBI seems to be more beneficial to participants that exhibit high levels of dispositional mindfulness and/or wellbeing prior the intervention. This has important implications regarding the appropriateness of low-dose MBIs in future research and therapy. |  |
| Wong, et al., (2019).<br>Efficacy of a | **Study design:** Mixed method uncontrolled before and after study. | **Sample Size:** 56 completed the study | **Primary findings:**<br>**Self-esteem** |  |
| <u>multicomponent intervention with animal-assisted therapy for socially withdrawn youth in Hong Kong.</u> society & animals, 27(5-6), 614-627.<br>(Hong Kong) | **Aim:** To assess the feasibility, efficacy and potential harm of the Regain Momentum (RM) program that involved dogs in enhancing self-esteem and perceived employability, as well as reducing social interaction anxiety of youth who have been withdrawn with and without mental health problems.<br>**Dates of data collection:** Not stated<br>**Data collection methods:** Rosenberg Self-Esteem Scale (RSES); Interaction Anxiousness Scale (IAS); Perceived Employability Self-Efficacy Scale (PESES); Unstructured individual and small group interviews were conducted within three months of completing the RM program with participants who had received the AAT component(s).<br>**Outcomes:** Self-esteem, anxiety, employability self-efficacy | **Participants:** Around 16% (n = 9) of the participants were older than 25 years and 7% (n = 4) were younger than 14 years old. 66.1% males, 33.9% females. 79% of participants were unemployed at intake.<br>About 70% (n = 39) were considered to be socially withdrawn (primary hikikomori), 22 of these subjects participated in the AAT, and 17 did not.<br>About 30% (n = 17) were withdrawn individuals with mental health problems, and they were considered to be secondary hikikomori; 15 of these subjects participated in the AAT, and 2 did not. Depressive or anxiety disorders (41%, n = 7) and psychosis (23%, n = 4) were the most common disorders among those with mental health problems.<br>**Setting:** Youth centre<br>**Intervention details:** Multicomponent intervention including four non animal-assisted therapy components (AAT) and four animal-assisted therapy components. The non-AAT components included (1) a hotline for referrers, parents, and socially withdrawn youths; (2) referral to further clinical assessment; (3) 3 to 5 face-to-face, individual counselling sessions with social workers; (4) support groups led by professionals. The AAT components included (5) AAT in the form of individual counselling and small group | The time × withdrawn groups interaction was marginally significant, (Wilk's Lambda=0.96; $F(1,71)=3.36$; $p=0.071$), with a small effect size (eta-squared=0.04). Mean RSES was significantly greater at post-test ($M=16.4$) than at pre-test ($M=14.5$) in the withdrawn group ($d=0.43$; $p=0.005$). The change from pre- to post-test fell short of significance in the group with mental health problems. There were no other significant interaction effects and main effects.<br>**Anxiety**<br>There was a statistically significant main effect for time (Wilk's Lambda=0.81; $F(1,71)=16.25$; $p<0.001$). Post-test IAS was significantly lower ($M=44.3$) than pretest IAS ($M=47.6$; $p=0.003$; $d=1.16$). The time × intervention groups interaction was statistically significant (Wilk's Lambda=0.94, $F(1,71)=4.48$; $p=0.038$), with a small effect size (eta-squared=0.04). Mean IAS was significantly lower at post-test ($M=45.3$) than at pre-test ($M=49.1$) in the non-AAT group, ($d=1.17$; $p=0.019$). The change from pre- to post-test fell short of significance in the AAT.<br>The time × withdrawn groups interaction was marginally significant (Wilk's Lambda=0.95, $F(1,71)=3.73$, $p=0.057$), with a small effect size (eta-squared=0.04). Mean IAS was significantly lower at post-test ($M=44.5$) than at pretest ($M=47.4$) in the withdrawn group ($d=1.14$; $p=0.032$) and significantly lower at post-test ($M=43.7$) than at pretest ($M=48.4$) in the group with mental health problems ($d=1.21$; $p=0.017$).<br>The time × intervention groups × withdrawal groups interaction was statistically significant (Wilk's Lambda=0.92, $F(1,71)=5.96$; $p=0.017$), with a moderate effect size (eta-squared=0.06). The significant triple interaction was probed by conducting simple main effects tests of the effect of time within each of the four groups. The change from pre- to post-test was significant only for the withdrawn with mental health problems without AAT. For the ones with mental health problems with no AAT, the means of IAS reduced from pre-test ($M=48.5$) to post-test ($M=33.5$; $d=1.48$; $p=0.014$).<br>**Self-efficacy**<br>There was a statistically significant main effect for time (Wilk's Lambda=0.94; $F(1,71)=4.56$; $p=0.038$). Post-test PESES was significantly greater ($M=48.6$) than pretest PESES ($M=44.8$; $p<0.001$; $d=0.96$). There were no other significant interaction effects and main effects.<br>**Additional findings:**<br>**Qualitative findings** |  |
|  | **Quality rating:** Low | activities with the therapy dog as a support to the trained social worker; (6) AAT in the form of social gatherings with dogs and volunteers from several dog visitation agencies; (7) a pre-employment training program focused on companion animal grooming; and (8) a pre-employment training program focused on taking care of the therapy dog in the youth centre. As a case management model was adopted, the mobilization and coordination of various service components were tailor-made to the participants' needs. In general, participants in the RM program received all the non-AAT components (1 to 4). They were then invited to receive the AAT components (5 to 8) on a voluntary basis. However, not all the participants in the RM program received the AAT components due to reasons such as cultural beliefs, previous negative experiences with nonhuman animals, or potential allergies to nonhuman animals. The combination of the service components was tailor made and formulated in conjunction with the participant to meet their personal goals. AAT was carried out by qualified social workers under the close supervision of a psychiatrist specialist through a monthly case conference to discuss and review the progress of the cases and to identify psychosocial factors underlying the social withdrawal behaviour. | Ten respondents who had participated in the AAT components were interviewed either individually or in small groups of 2 to 3. In general, participants gave positive feedback about the intervention. No potential harm was recorded in the case files or reported in the interviews. **Two major themes were identified:**<br>**1 The AAT component(s) served as a catalyst for program participation.** Some participants were attracted by the inclusion of dogs in the program, which aroused their interest to learn more about it. Among the ten participants interviewed, three participants stated that if it were not for the inclusion of nonhuman animals, they probably would not have joined the RM program at all. This result was echoed by findings of the intake assessment. Respondents were asked in the intake assessment if there were no AAT component, would they join the RM program. Around one fourth (23.2%) of the respondents stated that they would definitely not or probably not do so. Further, engagement through the therapy dog was found to have minimized the negative stigma attached to being "in counselling."<br>**2 The presence of the therapy dog(s) enhanced the therapeutic environment.** A majority of the interviewees expressed that the most helpful element in the RM project was feeling that the therapy dog(s) accepted them as they were. One participant who had previously experienced being bullied said he felt more at ease when interacting with the on-site therapy dog, whom he perceived to be "very straightforward and simple." Some participants originally felt that seeing a social worker was threatening and anxiety provoking; but the presence of the on-site therapy dog was enough to calm them down and provided a distraction from their anxiety. One participant expressed that she felt less depressed and that her mood improved with the therapy dog during her counselling session. The participants also spoke of having a sense of achievement after the dog grooming training session. Taking care of the on-site therapy dog provided the participants with a high level of satisfaction and a sense of worth. As one participant described, "If you helped them [the nonhuman animals], they would show their happiness directly, as if they wanted to say 'Thank you.'" |  |
|  |  | Dogs were the nonhuman animals used in the program. They served as an adjunct to the social workers either in individual or group sessions. The therapy dog supported the social workers in fulfilling two major therapeutic tasks: (1) facilitating personal expression and social interaction; and (2) creating a safe environment where the human-nonhuman animal relationship could be reflected on as mirroring the interpersonal issues of the participants. For the individual counselling sessions, an on-site therapy dog was recruited.<br>**Comparator intervention or control:** Pre-post comparisons were made for those with and without mental health conditions and those who received AAT compared to those who didn't. |  |  |

### 7.3 Quality appraisal

**Table 5.**
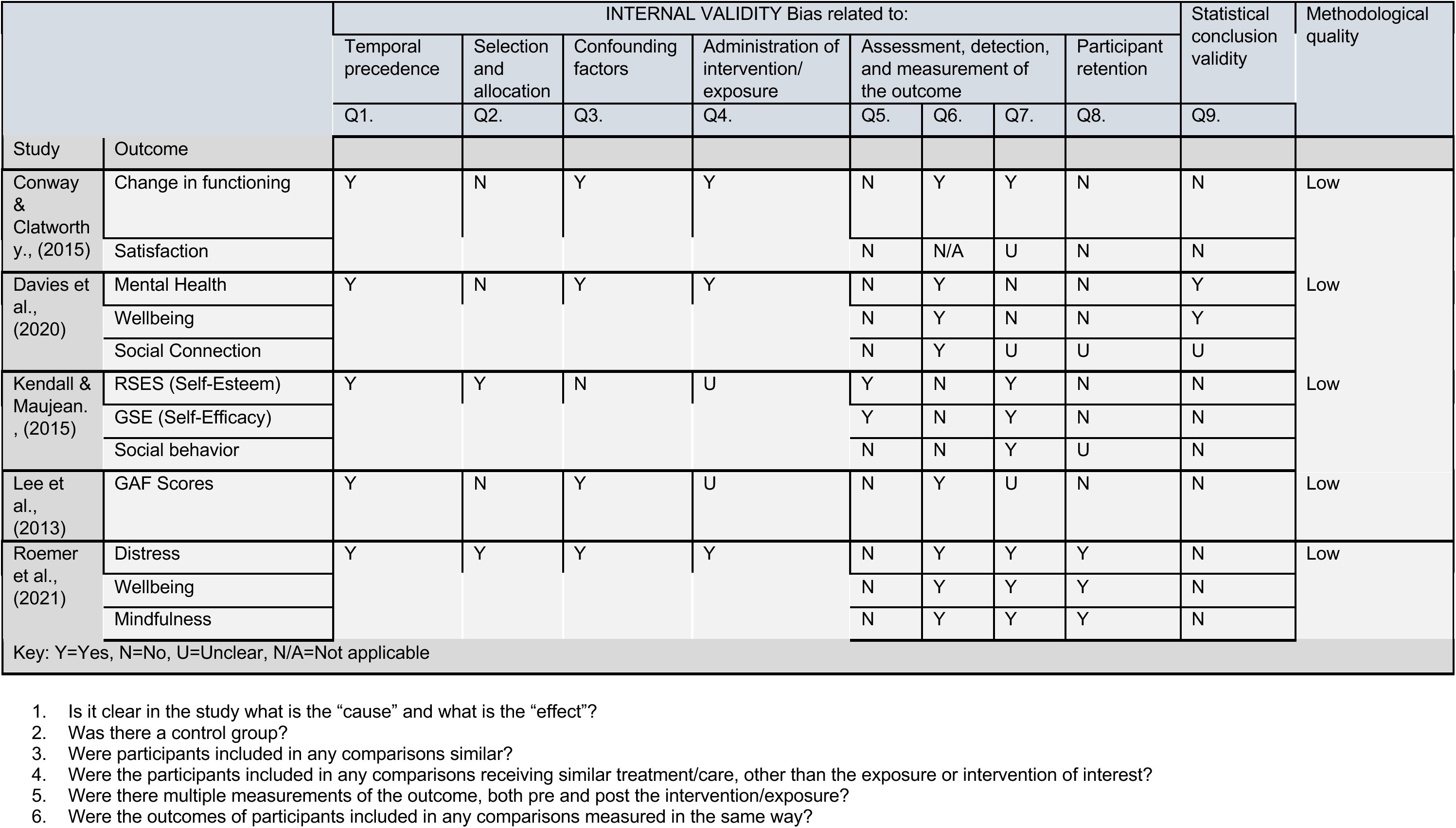
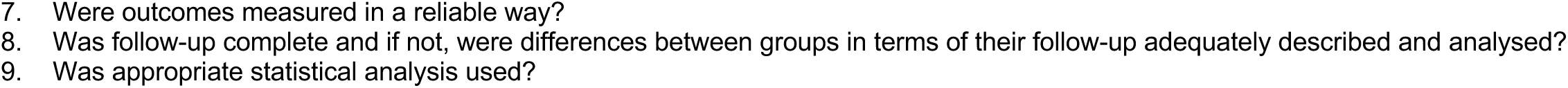
Quality appraisal results for quasi-experimental studies.

|  |  | INTERNAL VALIDITY Bias related to: |  |  |  |  |  |  |  | Statistical conclusion validity | Methodological quality |
| --- | --- | --- | --- | --- | --- | --- | --- | --- | --- | --- | --- |
|  |  | Temporal precedence | Selection and allocation | Confounding factors | Administration of intervention/ exposure | Assessment, detection, and measurement of the outcome |  |  | Participant retention |  |  |
|  |  | Q1. | Q2. | Q3. | Q4. | Q5. | Q6. | Q7. | Q8. | Q9. |  |
| Study | Outcome |  |  |  |  |  |  |  |  |  |  |
| Conway & Clatworth y., (2015) | Change in functioning | Y | N | Y | Y | N | Y | Y | N | N | Low |
|  | Satisfaction |  |  |  |  | N | N/A | U | N | N |  |
| Davies et al., (2020) | Mental Health | Y | N | Y | Y | N | Y | N | N | Y | Low |
|  | Wellbeing |  |  |  |  | N | Y | N | N | Y |  |
|  | Social Connection |  |  |  |  | N | Y | U | U | U |  |
| Kendall & Maujean. , (2015) | RSES (Self-Esteem) | Y | Y | N | U | Y | N | Y | N | N | Low |
|  | GSE (Self-Efficacy) |  |  |  |  | Y | N | Y | N | N |  |
|  | Social behavior |  |  |  |  | N | N | Y | U | N |  |
| Lee et al., (2013) | GAF Scores | Y | N | Y | U | N | Y | U | N | N | Low |
| Roemer et al., (2021) | Distress | Y | Y | Y | Y | N | Y | Y | Y | N | Low |
|  | Wellbeing |  |  |  |  | N | Y | Y | Y | N |  |
|  | Mindfulness |  |  |  |  | N | Y | Y | Y | N |  |
| Key: Y=Yes, N=No, U=Unclear, N/A=Not applicable |  |  |  |  |  |  |  |  |  |  |  |

1. Is it clear in the study what is the “cause” and what is the “effect”? 2. Was there a control group? 3. Were participants included in any comparisons similar? 4. Were the participants included in any comparisons receiving similar treatment/care, other than the exposure or intervention of interest? 5. Were there multiple measurements of the outcome, both pre and post the intervention/exposure? 6. Were the outcomes of participants included in any comparisons measured in the same way?
7. Were outcomes measured in a reliable way? 8. Was follow-up complete and if not, were differences between groups in terms of their follow-up adequately described and analysed? 9. Was appropriate statistical analysis used?

**Table 6.** Quality appraisal results for mixed methods studies.

| Study | Screening questions |  | Qualitative |  |  |  |  | Quantitative non-randomised |  |  |  |  | Mixed methods |  |  |  |  | Methodological quality |
| --- | --- | --- | --- | --- | --- | --- | --- | --- | --- | --- | --- | --- | --- | --- | --- | --- | --- | --- |
|  | S1. | S2. | Q1.1. | Q1.2. | Q1.3. | Q1.4. | Q1.5. | Q3.1. | Q3.2. | Q3.3. | Q3.4. | Q3.5. | Q5.1 | Q5.2. | Q5.3. | Q5.4. | Q5.5 |  |
| Bertotti et al., (2025) | Y | Y | Y | Y | Y | Y | Y | Y | Y | N | CT | Y | Y | Y | Y | Y | N | Low |
| De Lannoy, Graham & Grotte. (2024) | Y | Y | Y | Y | Y | Y | Y | CT | Y | CT | N | CT | Y | Y | Y | CT | N | Low |
| Wong, et al., (2019) | Y | Y | Y | Y | CT | CT | CT | CT | Y | CT | N | CT | CT | N | N | N | Y | Low |
| Key: Y=Yes, N=No, CT=Can't Tell |  |  |  |  |  |  |  |  |  |  |  |  |  |  |  |  |  |  |

Screening questions
S1. Are there clear research questions?
S2. Do the collected data allow to address the research questions?
Qualitative
1.1. Is the qualitative approach appropriate to answer the research question?
1.2. Are the qualitative data collection methods adequate to address the research question?
1.3. Are the findings adequately derived from the data?
1.4. Is the interpretation of results sufficiently substantiated by data?
1.5. Is there coherence between qualitative data sources, collection, analysis and interpretation?

**Table 7.**
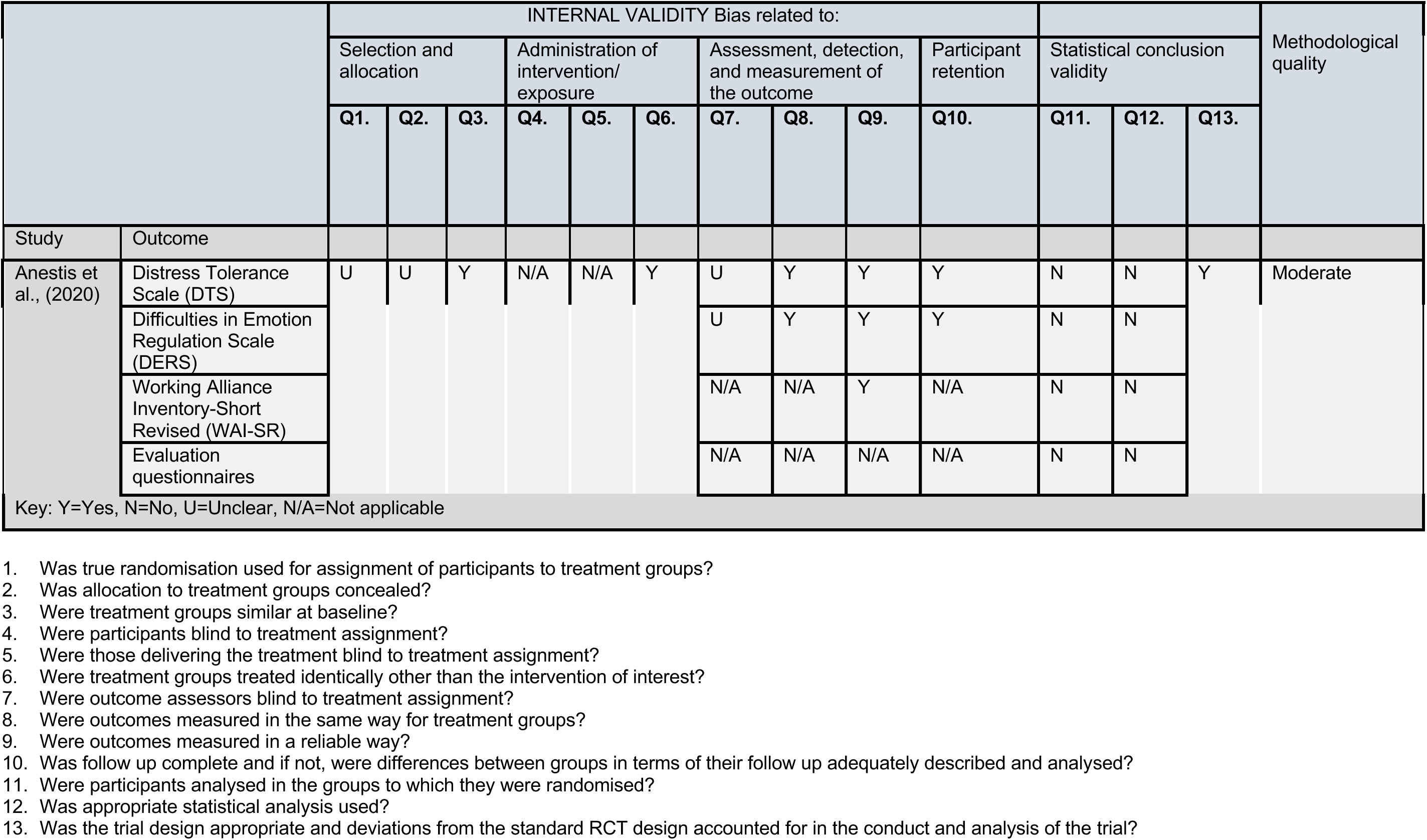
Quality appraisal results for randomised controlled trials.

### 7.4 Information available on request

The following are available on request: protocol; search strategies.

## 8. ADDITIONAL INFORMATION

### 8.1 Conflicts of interest

The authors declare they have no conflicts of interest to report.

#### Acknowledgements

The Public Health Wales team would like to thank Ben Williams, Kelly Davies and Olivia Gallen for their time, expertise, and contributions during stakeholder meetings in guiding the focus of the review and interpretation of findings.

## APPENDIX

### APPENDIX 1

#### MEDLINE search strategy

Ovid MEDLINE(R) ALL <1946 to June 06, 2025>

1. (neet* or "not in education employment or training" or NLFET* or "neither in the labour force nor in education or training" or "not in education employment or training" or "not in employment education or training" or "not participating in education employment or training" or "not participating in employment education or training" or "not being integrated into education employment or training" or "not being integrated into employment education or training" or "not engaged in education employment or training" or "not engaged in employment education or training" or "neither in employment nor in education or training" or "neither in work nor study" or "neither working nor studying" or "neither work nor study" or "neither in active employment nor in education or training" or "neither in employment nor education or training" or "neither in employment nor education and training" or "neither in work nor school" or "neither in employment nor in education" or ("neither work* nor enrol* in" adj2 education) or "neither employment nor training" or "neither in school nor in employment" or (“neither work* nor continu*" adj2 education) or "neither education nor employment" or "neither in school working nor seeking employment" or "neither in school nor in the workforce" or "neither education employment nor training" or "neither employment nor education" or "neither in school nor work*" or "not studying not working" or "neither working nor attending school" or "neither in employment nor education*" or "neither work nor education" or "neither employment, education nor training" or "neither working nor going to school" or "neither worked nor studied" or "neither work nor education" or "neither education nor work*" or "not in employment education or daily activities" or "not being in education employment or training" or "not in education employment and training" or "not in school not employed" or "not in education or employment" or "not in education or work*" or "not study and do not work" or "neither studies nor works" or "neither in employment education nor training" or "neither in education employment nor training" or "not in employment or education and training" or "neither in education nor in employment or training" or "neither in employment nor in education or training" or "not in employment nor in education or training" or "neither in employment education or training" or "neither in education employment or training" or "neither working nor enrolled in school" or "neither working nor in school" or "neither been working nor studying").ti,ab. 470
2. (("drop* out of" or "drop* out from") adj2 (education* or school* or college* or universit* or employment or work or training or vocation* or job* or labor or labour)).ti,ab. 766
3. ("disengaged young" or "disengaged youth*" or "disengaged teen*" or "disengaged adolescen*" or "economically inactive youth*" or "economically inactive young" or "economically inactive teen*" or "economically inactive adolescen*").ti,ab. 14
4. ((jobless or unemployed or unemployment or "out of work") adj2 (young or youth* or teen* or adolescen* or juvenile*)).ti,ab. 508
5. or/1-4 1747
6. Unemployment/ or Student Dropouts/ 10000
7. Adolescent/ or Young Adult/ 2865353
8. 6 and 7 3207
9. 5 or 8 4601
10. (interven* or program* or scheme* or strateg* or initiative* or workshop* or project* or training or support or service* or outreach or coach* or mentor* or counsel* or communit* or trial*).ti,ab. 7854994
11. 9 and 10 2466

